# Recommending Drug Combinations using Reinforcement Learning to target Genes/proteins that cause Stroke: A comprehensive Systematic Review and Network Meta-analysis

**DOI:** 10.1101/2023.04.20.23288906

**Authors:** Mahnaz Boush, Ali A. Kiaei, Danial Safaei, Sadegh Abadijou, Nader Salari, Masoud Mohammadi

## Abstract

**Objectives:** *<u>(Importance)</u>* Cerebrovascular accident (Stroke) is a term used in medicine to describe cutting off blood supply to a portion of the brain, which causes tissue damage in the brain. Clots of blood that form in the brain’s blood vessels and ruptures in the brain’s blood vessels are the root causes of cerebrovascular accidents. Dizziness, numbness, weakness on one side of the body, and difficulties communicating verbally, writing, or comprehending language are the symptoms of this condition. Smoking, being older and having high blood pressure, diabetes, high cholesterol, heart disease, a history of cerebrovascular accident in the family, atherosclerosis (which is the buildup of fatty material and plaque inside the coronary arteries), or high cholesterol all contribute to an increased risk of having a cerebrovascular accident. *<u>(Objective)</u>* This paper analyzes available studies on Cerebrovascular accident medication combinations.

**Evidence acquisition:** *<u>(Data sources)</u>* This systematic review and network meta-analysis analyzed the Science Direct, Embase, Scopus, PubMed, Web of Science (ISI), and Google Scholar databases without a lower time limit and up to July 2022. A network meta-analysis examines the efficacy of this drug combination on genes/proteins that serve as progression targets for cerebrovascular accidents.

**Results and Conclusion:** In scenarios 1 through 3, the p-values for the suggested medication combination and Cerebrovascular accident were 0.036633, 0.007763, and 0.003638, respectively. Scenario I is the combination of medications initially indicated for treating a cerebrovascular accident. The recommended combination of medications for cerebrovascular accidents is ten times more effective. This systematic review and network meta-analysis demonstrate that the recommended medication combination decreases the p-value between cerebrovascular accidents and the genes as potential progression targets, thereby enhancing the treatment for cerebrovascular accidents. The optimal combination of medications improves community health and decreases per-person management costs.

**Highlights:**

- Combined drugs that make the p-value between Stroke and target genes close to 1
- Using Reinforcement Learning to recommend drug combination
- A comprehensive systematic review of recent works
- A Network meta-analysis to measure the comparative efficacy
- Considered drug interactions

## INTRODUCTION

A stroke, also known as a cerebrovascular accident (CVA), is an abrupt disruption of the blood flow to the brain or the vasculature. Ischemic strokes account for about 85 percent of all cases, while hemorrhagic strokes make up the remaining 15 percent. The number of people who suffer from strokes and die because of having one has been steadily declining over the past few decades. A stroke is the most common cause of adult disability in any region of the world. It is, therefore, extremely important to diagnose the symptoms of a stroke early on and begin treatment as soon as possible to prevent or reduce the risk of morbidity and fatality. Any number of factors could have brought on a stroke. For instance, the most common cause of an ischemic stroke is high blood pressure, sometimes known as hypertension. Several other factors can lead to a younger person having a stroke, such as clotting abnormalities, carotid dissection, and the use of illegal drugs.(Khaku and Tadi, 2017)

### Rationale

Various drugs have been proposed to block the receptors for these human genes to manage this disease. Among these drugs, as shown in Table 1, the following can be mentioned: Tissue Plasminogen Activator, Prasugrel Hydrochloride, Tenecteplase, Warfarin, edoxaban, danaparoid, Salicylic Acid, Clopidogrel, Ticlopidine, Ticagrelor, Rivaroxaban, and Apixaban. Among these drugs, some of them have been mentioned in various articles as effective drugs in the management of Cerebrovascular accidents, such as: *<u>Salicylic Acid</u>* is a chemical that may be extracted from the bark of white willow trees and the leaves of wintergreen plants. It can also be manufactured in a laboratory. It inhibits the growth of bacteria and fungi, and it also removes dead skin cells (keratolytic activity).

**Table 1:**
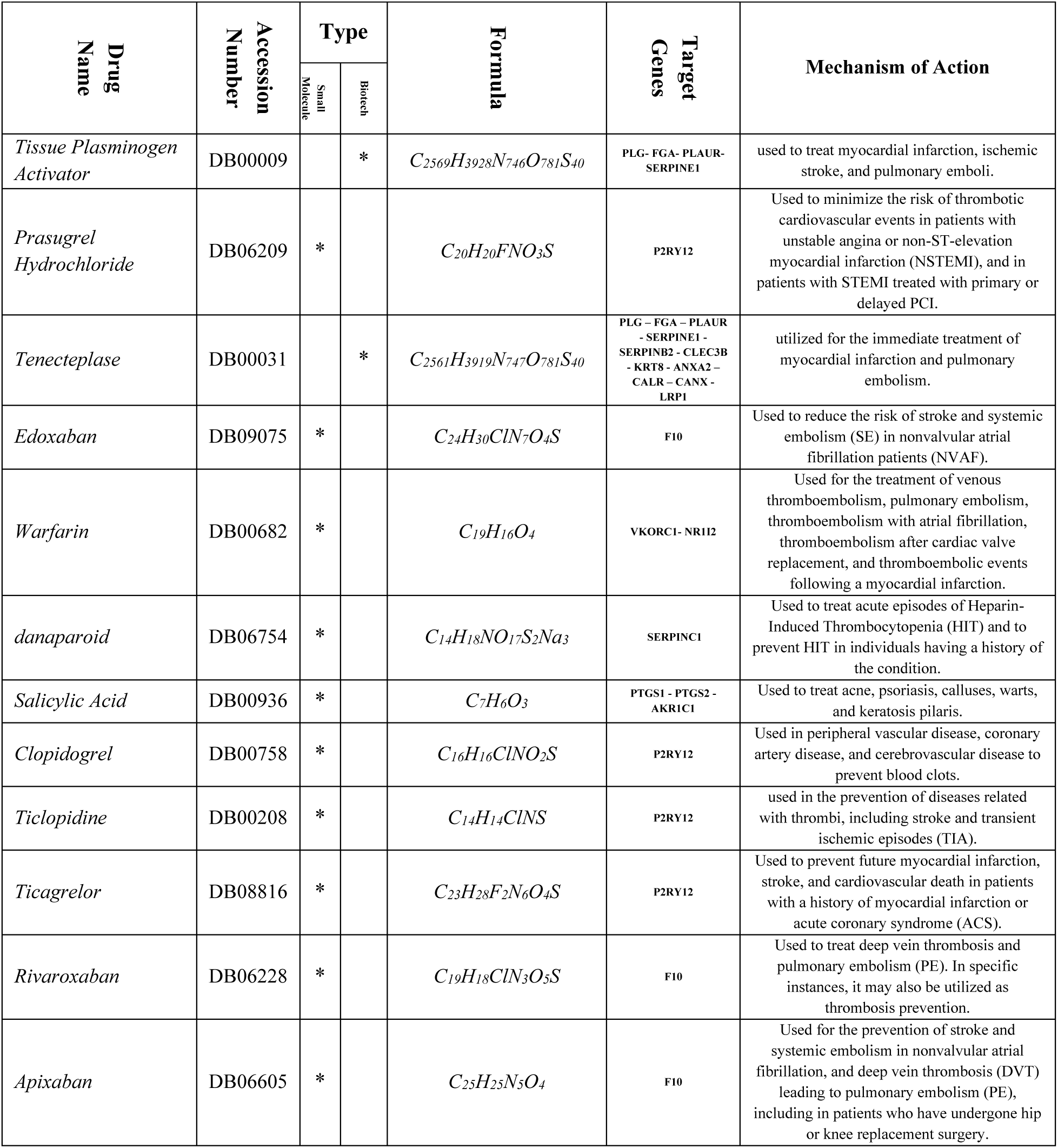
properties of some drugs that are mentioned in different studies as effective drugs to Cerebrovascular accident management.

| Drug Name | Accession Number | Type |  | Formula | Target Genes | Mechanism of Action |
| --- | --- | --- | --- | --- | --- | --- |
|  |  | Small Molecule | Biotech |  |  |  |
| *Tissue Plasminogen Activator* | DB00009 | | * | $C_{2569}H_{3928}N_{746}O_{781}S_{40}$ | PLG - FGA - PLAUR - SERPINE1 | used to treat myocardial infarction, ischemic stroke, and pulmonary emboli. |
| *Prasugrel Hydrochloride* | DB06209 | * | | $C_{20}H_{20}FNO_3S$ | P2RY12 | Used to minimize the risk of thrombotic cardiovascular events in patients with unstable angina or non-ST-elevation myocardial infarction (NSTEMI), and in patients with STEMI treated with primary or delayed PCI. |
| *Tenecteplase* | DB00031 | | * | $C_{2561}H_{3919}N_{747}O_{781}S_{40}$ | PLG - FGA - PLAUR - SERPINE1 - SERPINB2 - CLEC3B - KRT8 - ANXA2 - CALR - CANX - LRP1 | utilized for the immediate treatment of myocardial infarction and pulmonary embolism. |
| *Edoxaban* | DB09075 | * | | $C_{24}H_{30}ClN_7O_4S$ | F10 | Used to reduce the risk of stroke and systemic embolism (SE) in nonvalvular atrial fibrillation patients (NVAf). |
| *Warfarin* | DB00682 | * | | $C_{19}H_{16}O_4$ | VKORC1 - NR1I2 | Used for the treatment of venous thromboembolism, pulmonary embolism, thromboembolism with atrial fibrillation, thromboembolism after cardiac valve replacement, and thromboembolic events following a myocardial infarction. |
| *danaparoid* | DB06754 | * | | $C_{14}H_{18}NO_{17}S_2Na_3$ | SERPINC1 | Used to treat acute episodes of Heparin-Induced Thrombocytopenia (HIT) and to prevent HIT in individuals having a history of the condition. |
| *Salicylic Acid* | DB00936 | * | | $C_7H_6O_3$ | PTGS1 - PTGS2 - AKR1C1 | Used to treat acne, psoriasis, calluses, warts, and keratosis pilaris. |
| *Clopidogrel* | DB00758 | * | | $C_{16}H_{16}ClNO_2S$ | P2RY12 | Used in peripheral vascular disease, coronary artery disease, and cerebrovascular disease to prevent blood clots. |
| *Ticlopidine* | DB00208 | * | | $C_{14}H_{14}ClNS$ | P2RY12 | used in the prevention of diseases related with thrombi, including stroke and transient ischemic episodes (TIA). |
| *Ticagrelor* | DB08816 | * | | $C_{23}H_{28}F_2N_6O_4S$ | P2RY12 | Used to prevent future myocardial infarction, stroke, and cardiovascular death in patients with a history of myocardial infarction or acute coronary syndrome (ACS). |
| *Rivaroxaban* | DB06228 | * | | $C_{19}H_{18}ClN_3O_5S$ | F10 | Used to treat deep vein thrombosis and pulmonary embolism (PE). In specific instances, it may also be utilized as thrombosis prevention. |
| *Apixaban* | DB06605 | * | | $C_{25}H_{25}N_5O_4$ | F10 | Used for the prevention of stroke and systemic embolism in nonvalvular atrial fibrillation, and deep vein thrombosis (DVT) leading to pulmonary embolism (PE), including in patients who have undergone hip or knee replacement surgery. |

*<u>Tissue Plasminogen Activator</u>* is an enzyme produced naturally in the body that assists in the dissolution of blood clots. A synthetic version of this enzyme is produced in the laboratory to effectively treat heart attacks, strokes, and lung clots. In addition, it is being investigated as a potential treatment for cancer. A form of a systemic thrombolytic agent known as tissue plasminogen activator is described here. Additionally referred to as tPA.

*<u>Warfarin</u>* is a medication known as an anticoagulant typically taken to stop the formation of blood clots and their movement. Warfarin is now the oral anticoagulant prescribed the most frequently in North America, even though it was first sold on the market as a pesticide under various brand names, including d-Con and Rodex. When used for medical purposes, warfarin possesses several characteristics that should be considered. One of these characteristics is its capacity to cross the placental barrier, which, when present in a pregnant woman, can lead to fetal bleeding, spontaneous abortion, premature birth, stillbirth, and death in the neonate. The use of warfarin has been linked to several additional negative effects, including necrosis, purple toe syndrome, osteoporosis, calcification of the valves and arteries, and interactions with other medications. Warfarin does not influence the viscosity of the blood; rather, it inhibits the vitamin K-dependent synthesis of biologically active versions of several different clotting factors in addition to many regulatory factors.

### Objectives

We decided to conduct a systematic review of the existing studies in this field to provide a holistic yet comprehensive meta-analysis of the impact of drug combinations on the management of cerebrovascular accidents. Numerous adverse effects of drugs on managing cerebrovascular accidents have been reported, and there are no global statistics on the subject (Figure 1). This study aims to investigate, conduct a comprehensive literature review, and conduct an in-depth analysis of published findings regarding the effects of recommended medication combinations for treating cerebrovascular accidents.

**Figure 1:**
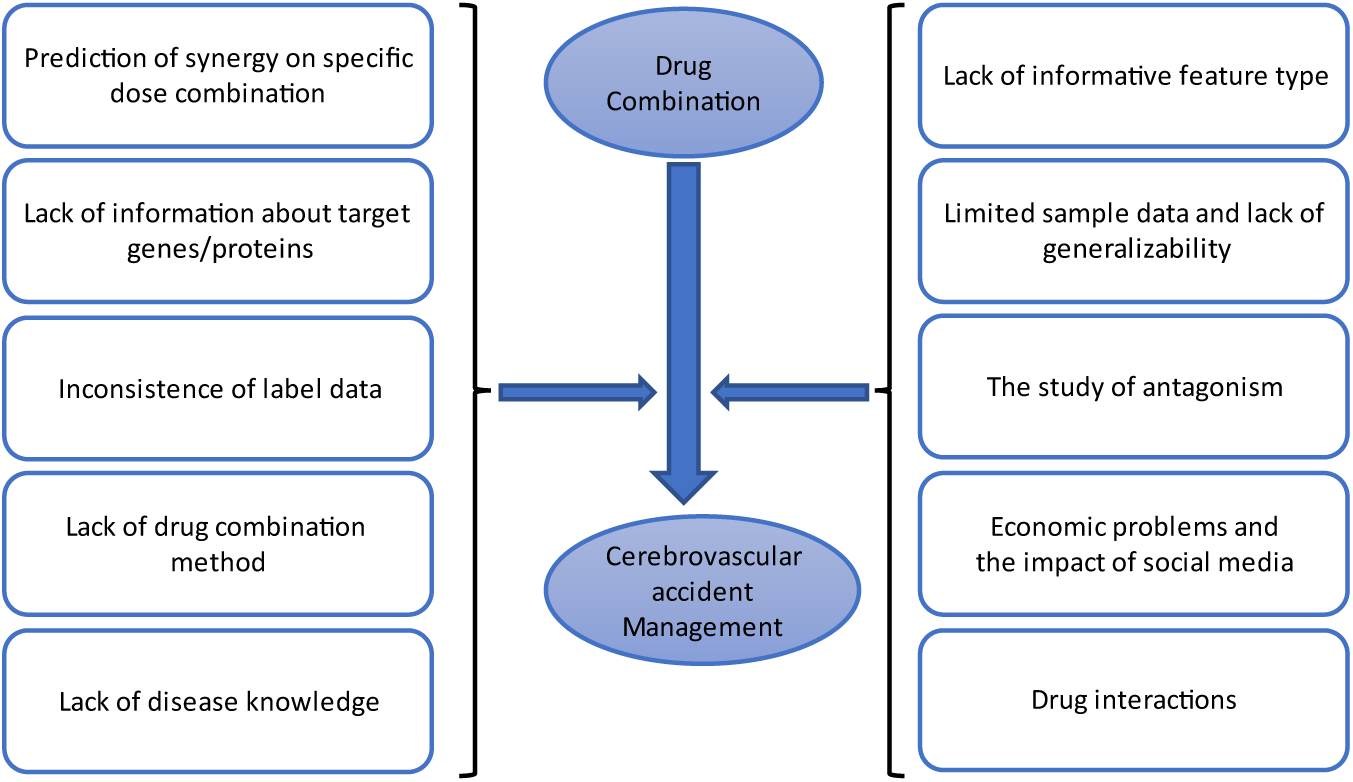
The effects of proposed drug combinations on the management of cerebrovascular incidents.

## METHOD

Our method consists of three stages. The first is implementing a reinforcement learning to recommend drug combinations for the disease. To evaluate the results of RL in the second stage, we have used a comprehensive systematic review and searched the recent articles that report the effect of such a combination on different populations (with a variety of ages, sex, etc.). We have employed Natural Language Processing (NLP) to search these articles since it uses context. We compared different manual and NLP-based searches and found that NLP could find articles based on the MeSH, not just the words we gave to the search engines. After evaluating the results of RL using reports in Systematic Review, in stage three, we evaluate the results of RL using the Network meta-analysis.

### Stage I: Reinforcement Learning

During the first stage of this investigation, a reinforcement learning (RL) model suggested some alternative medication combinations for the treatment of cerebrovascular accidents. The RL states consist of information regarding p-values between diseases and linked biological data, human genes, and information regarding p-values between the same human genes and effective medications. In addition, the reward is the p-value between the disease and the identical human genes after the medication combination.

#### State-value function

Using the Bellman equation in finding the state value, we have:

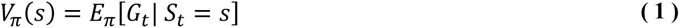

where *s* is state, *π* is policy, *V* is state value, *t* is time step, and *G*_*t*_ is the return (Gain) that is computed by:

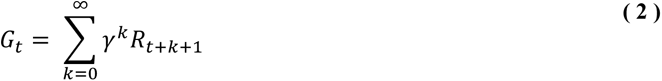

where *γ* is discount factor and R is the reward. For our purpose, if the final step of each episode is demonstrated by *T*, we have:

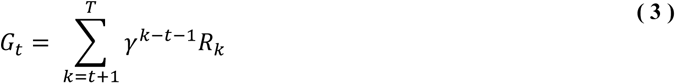

Back to our state value function, we can rewrite the formula as follows:

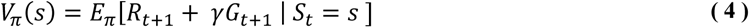

We can separate the parts of the state-value function as follow:

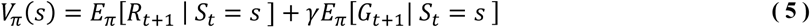

The first part can be computed by:

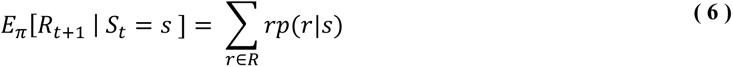

Where:

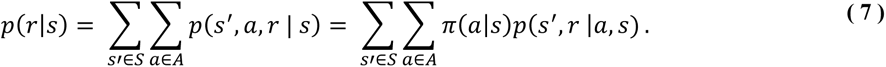

Therefore, the first part of the state-value function is as follows:

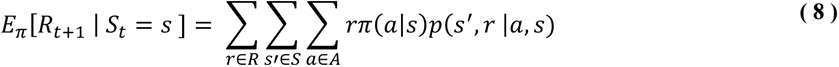

For the second part, we have the following:

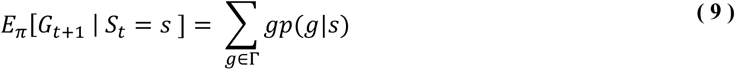

where

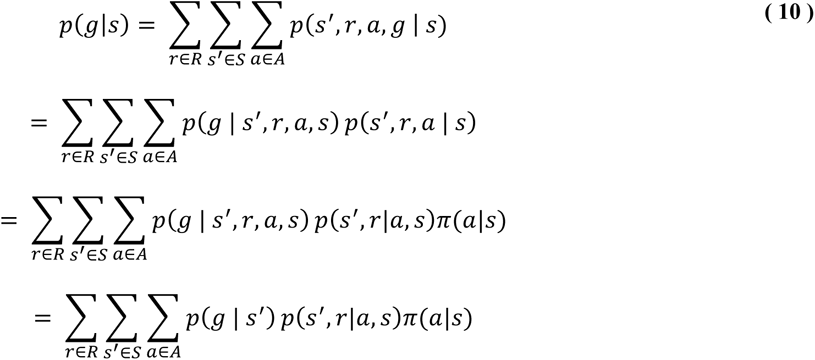

So, we have:

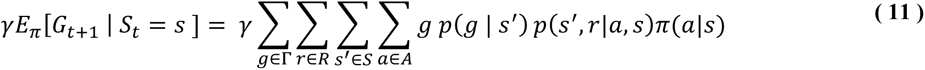

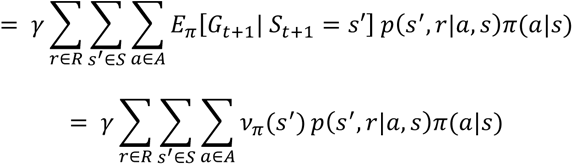

#### Action-value function

Since above, the action value function is computed by:

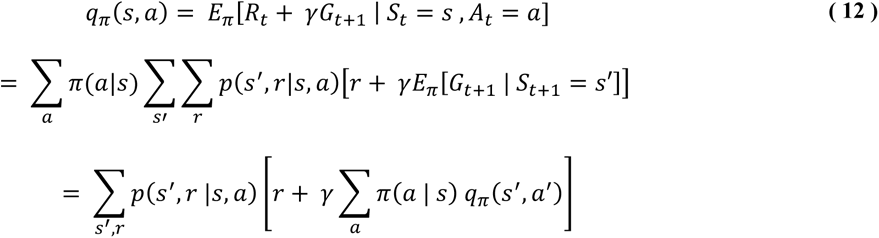

So, the current timestep’s state values and action values are computed recursively in terms of future values.

#### The Deep Neural Network used in the RL

The Deep Neural Network (DNN) model receives an input weight between Target (Cerebrovascular accident) and Associations (Drugs) in the form of an Interface Feature, which is composed of biological data (human gene). The p-value is used to determine this weight. Figure 2 shows the structure of our DNN model. The p-value between the Target and human genes is first considered in this figure. For example, *p*_1_^*FT*^ represents the p-value between the human gene F1 (as interface feature) and the Cerebrovascular accident (as Target). On the other hand, this model receives the p-value between these genes and different drugs. (For example, *p*_13_^*AF*^ represents the p-value between the first Association and the third Interface human gene.) Using the above biological data as inputs, the DNN model estimates the combined p-value between Associations and the Target. (For example, *cp*_2_^*AT*^ represents the combined p-value between the second association and Target.)

**Figure 2:**
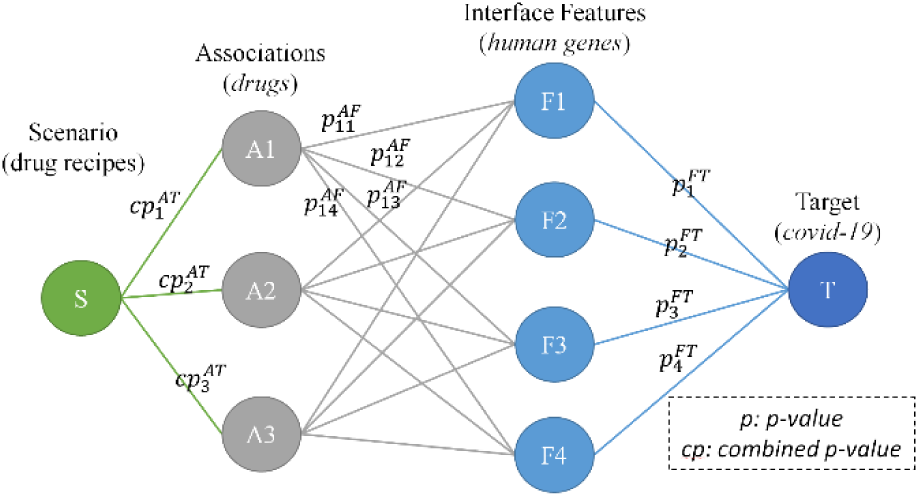
The general structure of the DNN model to suggest an effective drug combination in the management of disease using human genes as interface features

After selecting the association with the lowest combined p-value, the DNN model will update each weight based on the extent to which the chosen association influences the Interface attributes. This update is performed so that associations with the same effect on human genes as the selected association will have a larger p-value following its application. This process is repeated until the DNN model’s stop condition is met.

The DNN model selected medications based on their low p-values when correlated with cerebrovascular accidents. The drug selection algorithm computes the p-values of these pharmaceuticals in relation to cerebrovascular accidents. The following step is to choose the medication with the combined p-value that is the lowest. The algorithm for selecting drugs then modifies the weights assigned to cerebrovascular accidents and affected human genes based on the p-values associated with those genes and the initially selected drug. Therefore, the combined p-values between these medications and the disease have been updated. In each iteration of the drug selection procedure, the drug with the smallest combined p-value is sought, and the weights are adjusted accordingly. The algorithm for drug selection generated a series of scenario proposals, each of which included a drug recipe. This was done to encourage associations to collaborate, which was the ultimate goal of the algorithm.

### Stage II: A comprehensive Systematic Review

The findings of our RL model must be validated through a systematic evaluation of the first stage’s recommended medications in the second stage. In other words, we decided to utilize the systematic review to validate the work that we had already accomplished. This is done so that the outcomes of the initial stage can be utilized. Utilizing databases such as Science Direct, Embase, Scopus, PubMed, and Web of Science, we accessed previously published articles for review (ISI). Google Scholar was also used. Rather than conducting a manual search within these databases at this stage, a semantic search technique based on Natural Language Processing (NLP) is employed. This has the advantage of searching MeSH for every single term. For instance, a semantic search for the term “artificial intelligence” is equivalent to searching for the following keywords: “RL,” “Deep Learning,” “Transfer Learning,” “Transformers methods,” “SVM,” and “Reinforcement learning,” as well as other related keywords, some of which we may not be familiar with. It is believed that a broader and more accurate range of articles will be investigated in a relatively short time. In the third step, the effects of proposed synthetic drug combinations on human genes are examined using a network meta-analysis. In this study, the efficacy of each drug is evaluated based on the input biological data, which consists of human genes altered by a cerebrovascular accident. Then, in **Table 3**, Before and after the treatment combination, the p-value between human genes and Cerebrovascular accident is investigated. After the proposed drug combinations have been validated in steps two and three, step four involves the research of prescription drug information.

#### Information sources

At this point, an NLP-based systematic review is conducted to identify relevant studies within the Science Direct, Embase, Scopus, PubMed, and Web of Science databases (ISI). In addition, Google Scholar is searched. Even though our in-house RL model provides the suggested drug combination, as previously stated, the distinguishing feature of this article is not the use of this model but rather the validation of the suggested drug combination. By analyzing data from a large-scale clinical trial, this was accomplished. Thus, we relied on the systematic review to validate the findings of our investigation. The initial phase of the process involves extracting keywords from the outputs of the RL model and the Cerebrovascular accident subscription. Cerebrovascular accident, Tissue Plasminogen Activator, Prasugrel Hydrochloride, Tenecteplase, Warfarin, edoxaban, danaparoid, Salicylic Acid, Clopidogrel, Ticlopidine, Ticagrelor, Rivaroxaban, and Apixaban are the terms listed.

#### Search strategy

A natural language processing technique has been used by linking to these databases to conduct a semantic search of publication titles and abstracts. Due to the immense value of semantic search, MeSH phrases can be considered potential search keywords. For example, the word Cerebrovascular accident in semantic search is equivalent to Strokes, Cerebrovascular Accident, Cerebrovascular Accidents, CVA (Cerebrovascular Accident), CVAs (Cerebrovascular Accident), Cerebrovascular Apoplexy, Brain Vascular Accident, Brain Vascular Accidents, Cerebrovascular Stroke, Cerebrovascular Strokes, Apoplexy, Cerebral Stroke, Cerebral Stroke, Acute Stroke, Acute Strokes, Acute Cerebrovascular Accident, Acute Cerebrovascular Accidents, e Through November 2020, there is a time limit on searching for related articles in each database.

#### Eligibility criteria and Selection process

The following phases are included in studies selected for systematic review: 1) Studies incorporating at least one of the proposed cerebrovascular accident drugs. 2) Study-based research 3) Text-based study Included in the systematic review’s entry requirements were: 1- the phrase cerebrovascular accident and papers containing at least one of the potential medications. 2- Observational-style studies (i.e., non-interventional studies) (i.e., non-interventional studies) Three studies whose entire texts were available. The following were the criteria for exclusion: 1- Disparate research, 2-studies lacking sufficient data, 3- duplicate sources, and 4-research with unclear methodology. There are five intervention studies.

#### Study selection

Initially, duplicate research is eradicated. During the evaluation phase, a list of all remaining research titles is compiled to filter the research in a structured manner. As part of the first phase of the systematic review and screening, the titles and abstracts of the remaining research are reviewed thoroughly, and many studies are omitted based on selection criteria. In the second step, the competency evaluation, the full text of the remaining research from the screening phase is thoroughly reviewed based on the criteria, thereby eliminating several unrelated studies. To avoid the influence of personal preference on resource selection, an expert and an NLP Question-Answering (QA) agent conduct independent research and data extraction. The expert must provide a complete and accurate explanation for why the research was not chosen.

In contrast, the QA agent assigns a score to each article based on the questions posed. The articles with the lowest ratings will be removed. Questions posed by the quality assurance agent include, “Is this medication effective for the treatment of cerebrovascular accident?” For each output of the intelligent system, “this drug” is replaced with a different drug, generating an independent question. If there is a disagreement between the expert and the QA agent’s output, the latter expert will review the contentious research. In the present study, 33 cases were chosen for the third stage after completing the abovementioned steps.

#### Quality evaluation

To evaluate the quality of the remaining publications, it was decided to use a customized checklist based on the specific type of research being conducted (in terms of both the validity of their methods and findings). Frequently, checklists based on the STROBE method are used to evaluate and critique the quality of observational studies. The checklist is divided into six levels or broad sections, including the following: title, abstract, introduction, methodology, results, and discussion. Since several of these scales include subscales, there are 32 fields in total (subscales). Each of these 32 domains represents a distinct aspect of a study’s methodology. Subscales include the title, problem statement, study objectives, study type, statistical population, sampling method, sample size, variables and procedures definitions, data collection methods, statistical analysis techniques, and results.

Consequently, a score of 32 is regarded as the maximum that can be achieved during the quality assessment phase using the STROBE checklist. When a score of 16 is used as the benchmark, any article with a score of 16 or higher is considered moderate or high, depending on the perspective. The sixteen articles with a score lower than 16 were disqualified from the study due to their low level of methodological quality. In the current investigation, the STROBE checklist was used to evaluate the quality of the publications. Then, 33 papers deemed medium or high quality moved on to the systematic review phase, which confirms the data gathered in the first stage. The RAIN model serves as the basis for the executive protocol of this paper. (Mohammadi et al., 2021). Numerous research studies have employed the RAIN protocol as a framework for investigating various psychological and emotional phenomena(Kiaei et al., 2022, 2023; Salari et al., 2023, 2022a, 2022b, 2021)

#### Study risk of bias assessment

At this stage, the p-value is used to determine how effectively each drug will influence human genes. In addition to **Table 3**, the results will be displayed as circular bar charts and radar charts.

### Stage III: A network meta-analysis

In this stage, we used Network meta-analysis to assess different drugs concurrently in a single study.

It Combined direct and indirect data between disease and drugs with Genes/Proteins as interface features within a network of randomized controlled trials. As it assists in measuring the comparative efficacy of commonly used drugs in clinical practice, it has gained popularity among physicians.

## RESULTS

### Stage I: Reinforcement Learning

Our RL-recommended drug combinations include salicylic acid, Tissue Plasminogen Activator, and Warfarin. Table 2 indicates the drugs mentioned above with the p-value of combining them. For instance, the p-value between Cerebrovascular accident and Salicylic Acid, Scenario 1 (S1), indicated 0.037 and decreased to 0.008 when Tissue Plasminogen Activator was added to the combination, *i.e.,* Scenario 2 (S2). Moreover, as seen in Table 2, the P-value after applying the third scenario has shown that the proposed drug combination had a good effect on managing the disease.

**Table 2:** p-value between scenarios and **Cerebrovascular accident**

| *Scenario* | *Drug Combinations* | *p-value* |
| --- | --- | --- |
| *S1* | **Salicylic Acid** | 0.036633 |
| *S2* | **S1 + Tissue Plasminogen Activator** | 0.007763 |
| *S3* | **S2 + Warfarin** | 0.003638 |

As shown in Table 3, the p-values between human genes and Cerebrovascular accidents have been changed by new scenarios. The ‘S0’ column shows the p-value between Cerebrovascular accident and the corresponding affected human genes. The “S1” column shows the combined p-value between Cerebrovascular accidents and the human genes in which Salicylic Acid is used. The p-values between Cerebrovascular accident and many human genes reach 1 in the ‘S3’ column, which indicates the weakening of the importance of the target genes.

**Table 3:** p-values between **Cerebrovascular accident** and human genes after implementing scenarios

| Association Name | S0 | S1 | S2 | S3 | Association Name | S0 | S1 | S2 | S3 |
| --- | --- | --- | --- | --- | --- | --- | --- | --- | --- |
| PLAT | 8E-06 | 1 | 1 | 1 | BDNF | 6E-04 | 0.97 | 1 | 1 |
| PLG | 1E-05 | 1 | 1 | 1 | PLA2G7 | 6E-04 | 0.98 | 1 | 1 |
| EEF1A2 | 4E-05 | 1 | 1 | 1 | F5 | 6E-04 | 1 | 1 | 1 |
| ABCD2 | 8E-05 | 0.99 | 1 | 1 | ALOX5AP | 6E-04 | 0.98 | 0.99 | 1 |
| NOTCH3 | 9E-05 | 0.65 | 0.65 | 0.65 | DGAT2 | 6E-04 | 0 | 0 | 0 |
| P2RY12 | 1E-04 | 1 | 1 | 1 | MMP9 | 7E-04 | 0 | 1 | 1 |
| F10 | 2E-04 | 1 | 1 | 1 | APP | 7E-04 | 0.98 | 0.98 | 1 |
| DOCK3 | 2E-04 | 0 | 0.21 | 0.52 | VWF | 7E-04 | 1 | 1 | 1 |
| CRP | 2E-04 | 1 | 1 | 1 | PDE4D | 7E-04 | 0 | 0.91 | 0.97 |
| MELAS | 2E-04 | 0.6 | 0.6 | 0.6 | PRH2 | 8E-04 | 1 | 1 | 1 |
| MS | 3E-04 | 0.99 | 1 | 1 | DFFA | 8E-04 | 0.84 | 0.84 | 0.84 |
| AWAT2 | 3E-04 | 0 | 0.43 | 0.43 | NINJ2 | 8E-04 | 0 | 0 | 0 |
| CIMT | 4E-04 | 0.6 | 0.84 | 0.84 | F2 | 8E-04 | 1 | 1 | 1 |
| RPSA | 4E-04 | 0.9 | 0.97 | 0.97 | PCSK9 | 9E-04 | 0.98 | 0.99 | 0.99 |
| IL4I1 | 4E-04 | 0.99 | 0.99 | 1 | SLC12A3 | 9E-04 | 0 | 1 | 1 |
| MTHFR | 5E-04 | 0.98 | 1 | 1 | GLA | 9E-04 | 0 | 0 | 1 |
| CLDN5 | 5E-04 | 0 | 1 | 1 | DCX | 9E-04 | 0 | 0 | 0 |
| RBFOX3 | 6E-04 | 0.84 | 0.84 | 0.84 | APOE | 1E-03 | 0.79 | 0.93 | 1 |
| CECR1 | 1E-03 | 0.51 | 0.51 | 0.51 | EDN1 | 0.001 | 0.93 | 1 | 1 |
| RNF213 | 1E-03 | 0 | 0 | 0 | BDNF-AS | 0.001 | 0.95 | 1 | 1 |
| GFAP | 1E-03 | 0.97 | 0.99 | 1 | KRIT1 | 0.001 | 0 | 0 | 0 |
| AQP4 | 0.001 | 0.84 | 0.99 | 0.99 | CYP2C19 | 0.001 | 1 | 1 | 1 |
| SELP | 0.001 | 1 | 1 | 1 | DGAT1 | 0.001 | 0.49 | 0.49 | 0.49 |
| SEMA5B | 0.001 | 0 | 0 | 0 | OCLN | 0.001 | 0.58 | 1 | 1 |
| CCM2 | 0.001 | 0 | 0 | 0 | GP6 | 0.001 | 1 | 1 | 1 |

### Stage II: A comprehensive Systematic Review

This stage investigates the effects of the medications mentioned above on treating cerebrovascular accidents.

Up until November 2020 and until July 2022, articles with this emphasis were gathered and systematically evaluated following PRISMA principles and the RAIN framework. (Mohammadi et al., 2021). After the preliminary investigation, two hundred forty-one potentially relevant articles were located and imported into the EndNote reference management system. 99 of the 241 listed studies were deemed duplicates and were therefore disregarded. Following a review of the titles and abstracts of the remaining 142 papers and consideration of the inclusion and exclusion criteria, 41 of the remaining studies were eliminated during the research’s screening phase. At the time that the eligibility of the studies was being evaluated, there were a total of 101 left, 56 were eliminated after their entire texts were reviewed and the inclusion and exclusion criteria were taken into account. At the stage of quality evaluation, based on the evaluation of the full text of the articles and the score obtained from the STROBE checklist for each paper, 12 of the remaining 45 studies were eliminated because they were deemed to be of poor methodological quality, leaving 33 cross-sectional studies for the stage of final analysis. This was achieved by analyzing the full texts of the articles and scoring each paper using the STROBE checklist. (Please see Figure 3). Details and characteristics of these articles are also provided in Table 4. The structures of these drugs are shown in Figure 4. Moreover, Table 5 shows the properties of these drugs.

**Figure 3:**
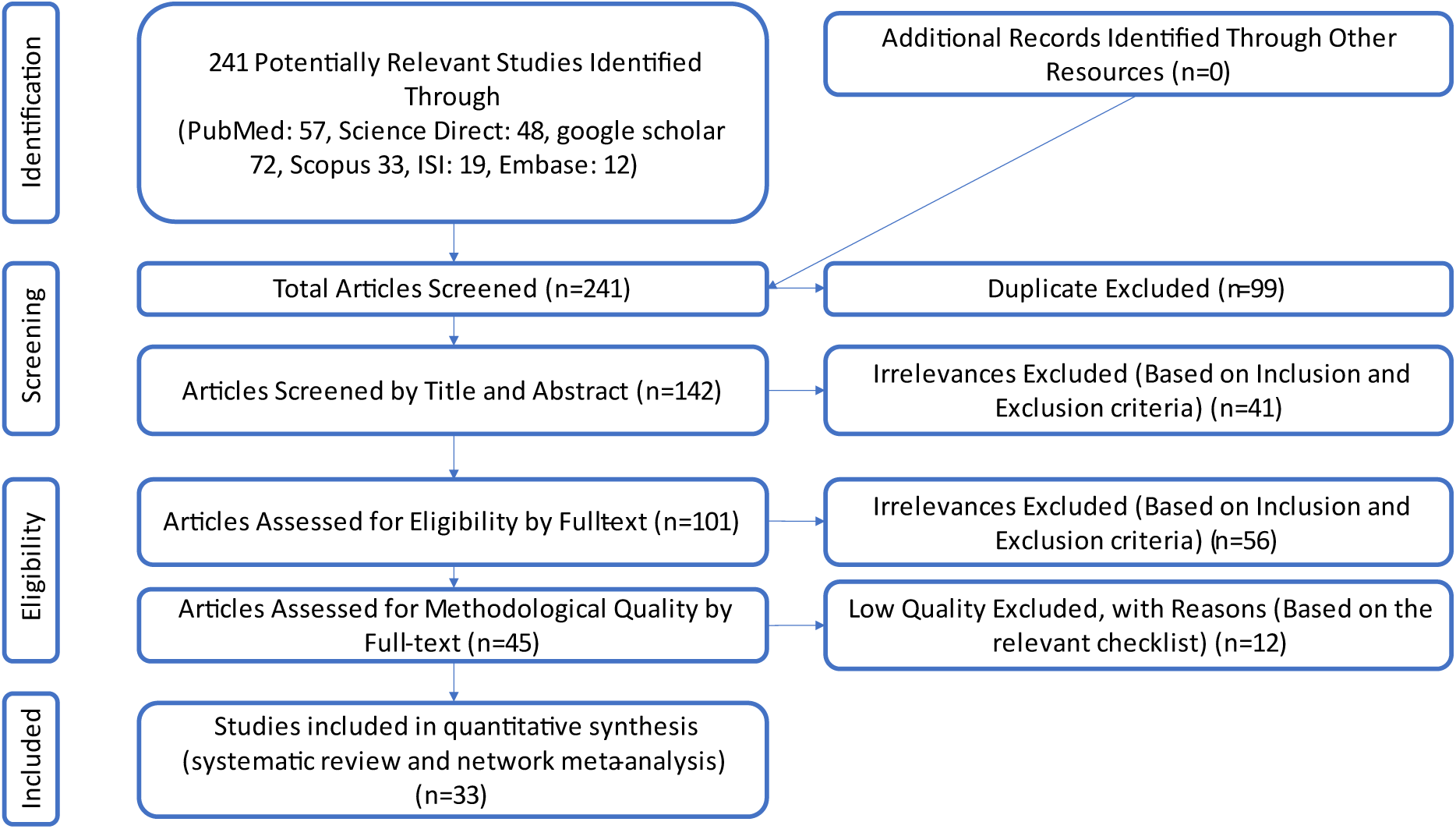
PRISMA (2020) flow diagram indicating the stages of sieving articles in this systematic review and network meta- analysis

**Figure 4:**
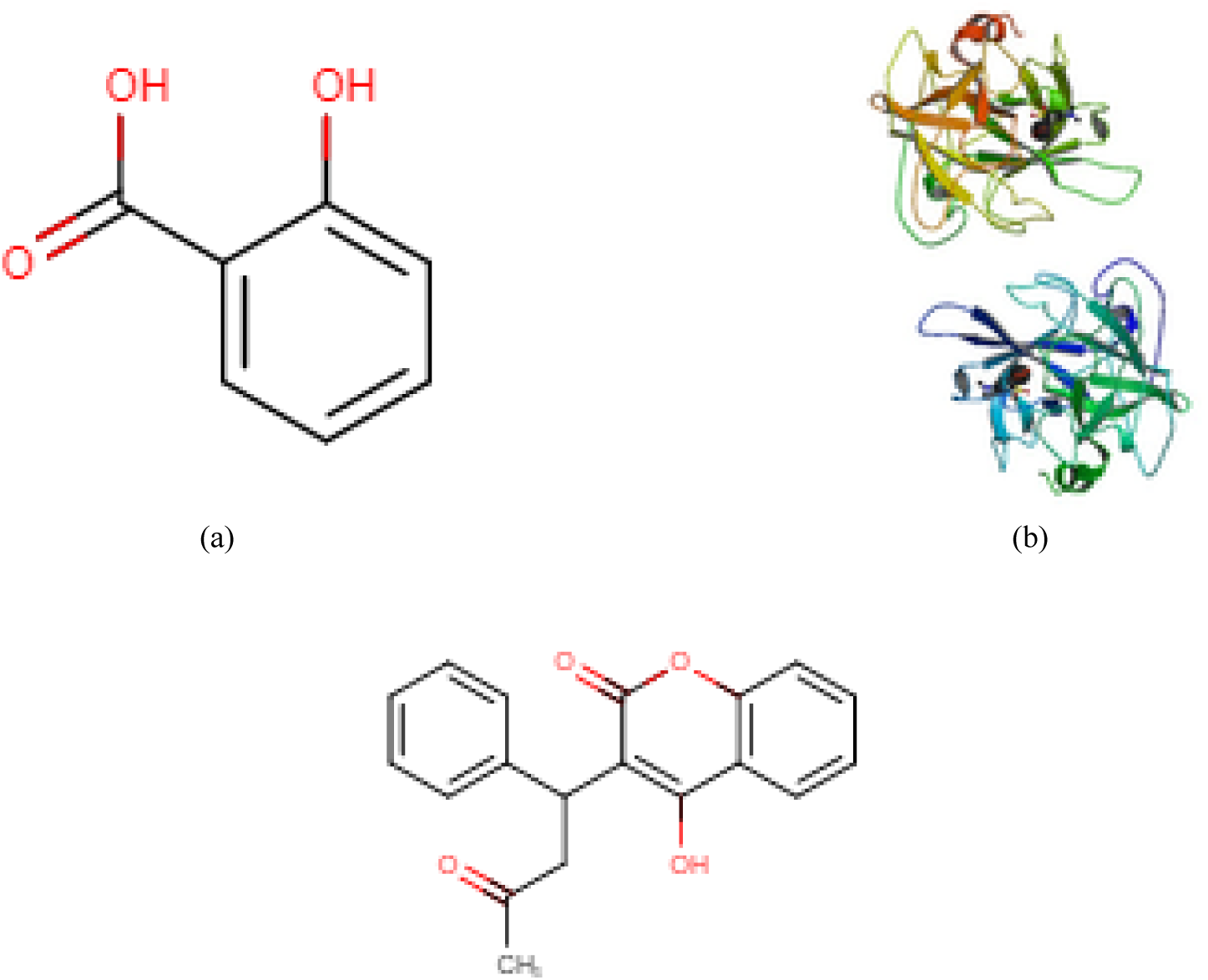
Drug structure for (a) Salicylic Acid, (b) Tissue Plasminogen Activator, (c) Warfarin from https://www.drugbank.com/

**Table 4:**
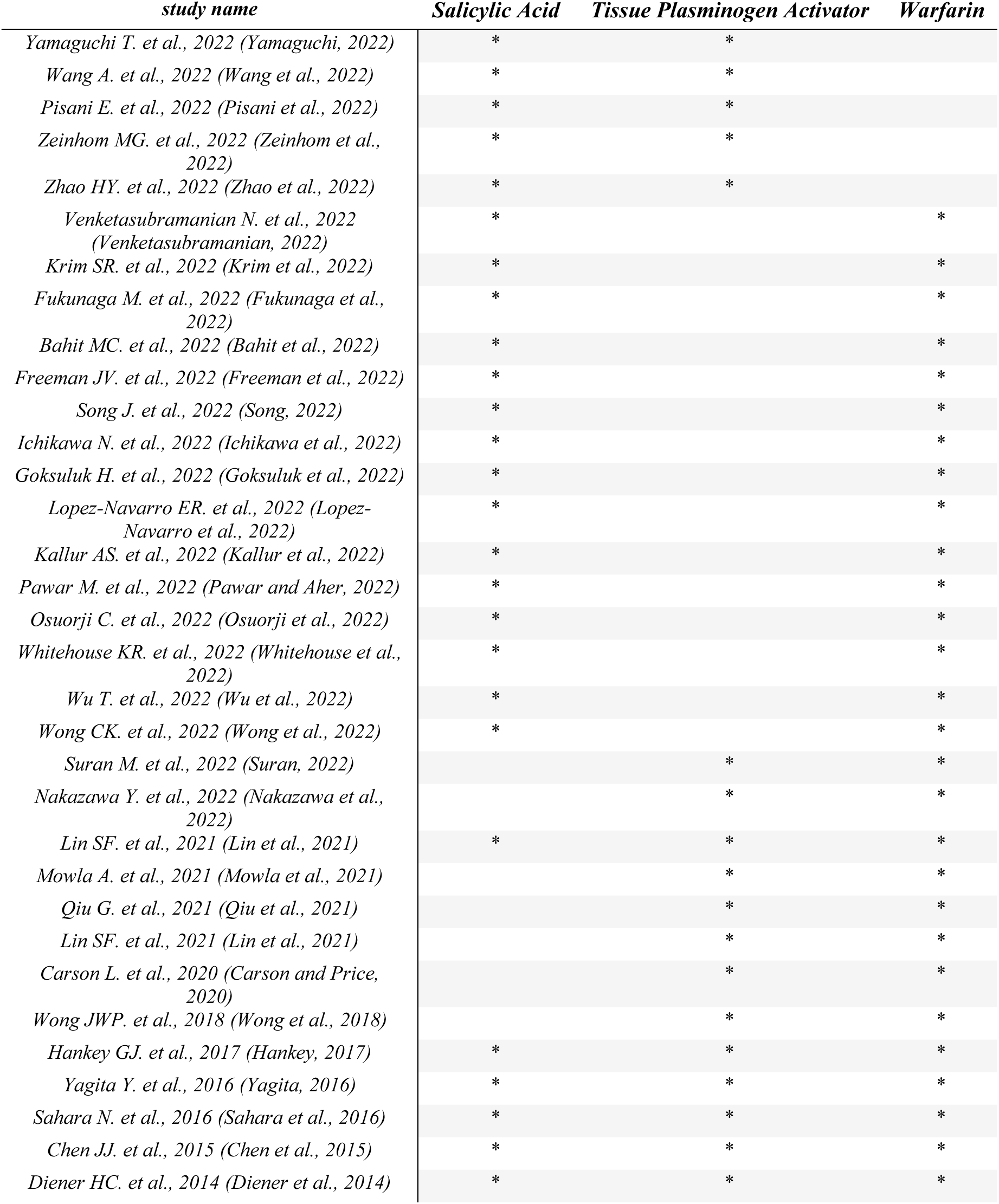
some important research studies for proposed drugs in Cerebrovascular accident managements

**Table 5:**
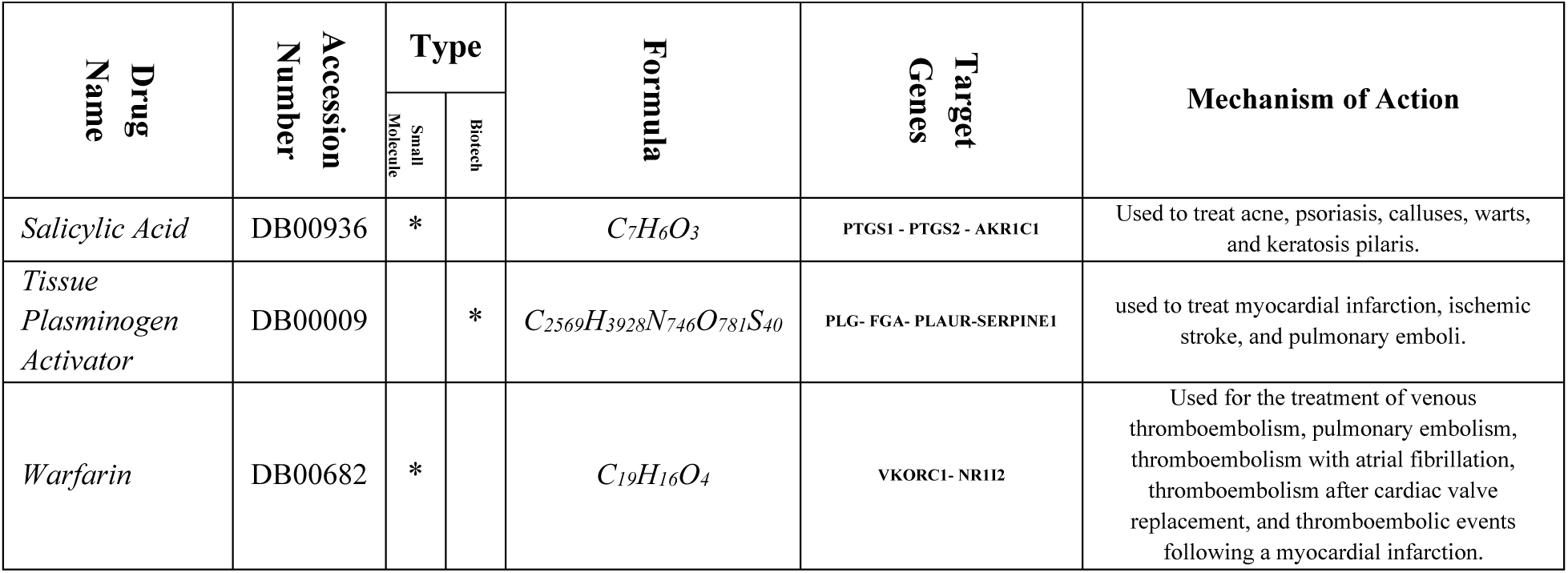
properties of proposed drugs as effective drugs to Cerebrovascular accident management.

### Stage III: Network Meta-Analysis

While Figure 5(a) indicates the p-values between affected human genes and Cerebrovascular accident, Figure 5(b) shows the p-values between them after implementing the third scenario. Moreover, Figure 6 shows a radar chart to indicate the efficiency of the drug selected using the drug selection algorithm by demonstrating the p-values between Cerebrovascular accident and human genes after consuming selected drugs. Each colored line shows the effectiveness of the corresponding drug in that scenario.

**Figure 5:**
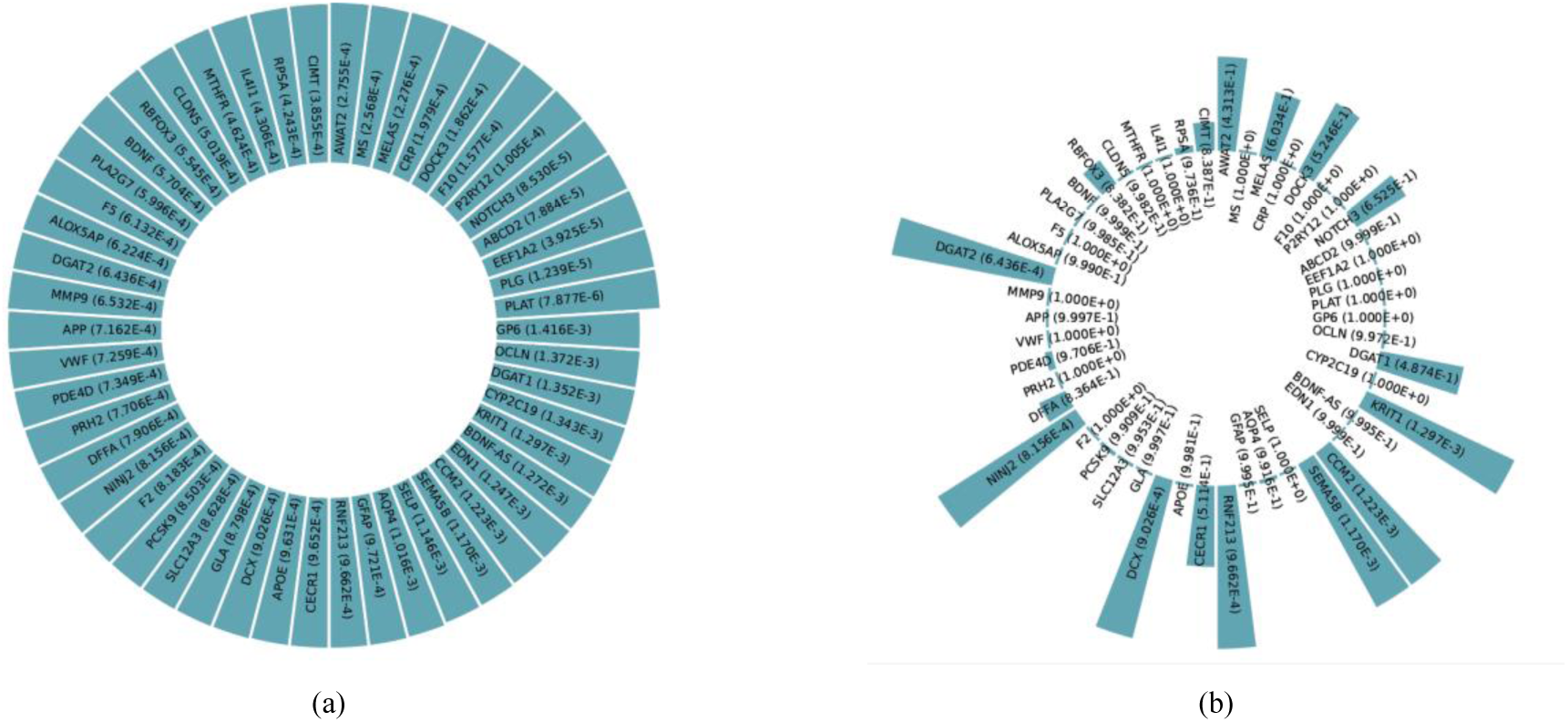
p-values between affected human genes and Cerebrovascular accident {(a)before/ (b)after} implementing Scenario 4.

**Figure 6:**
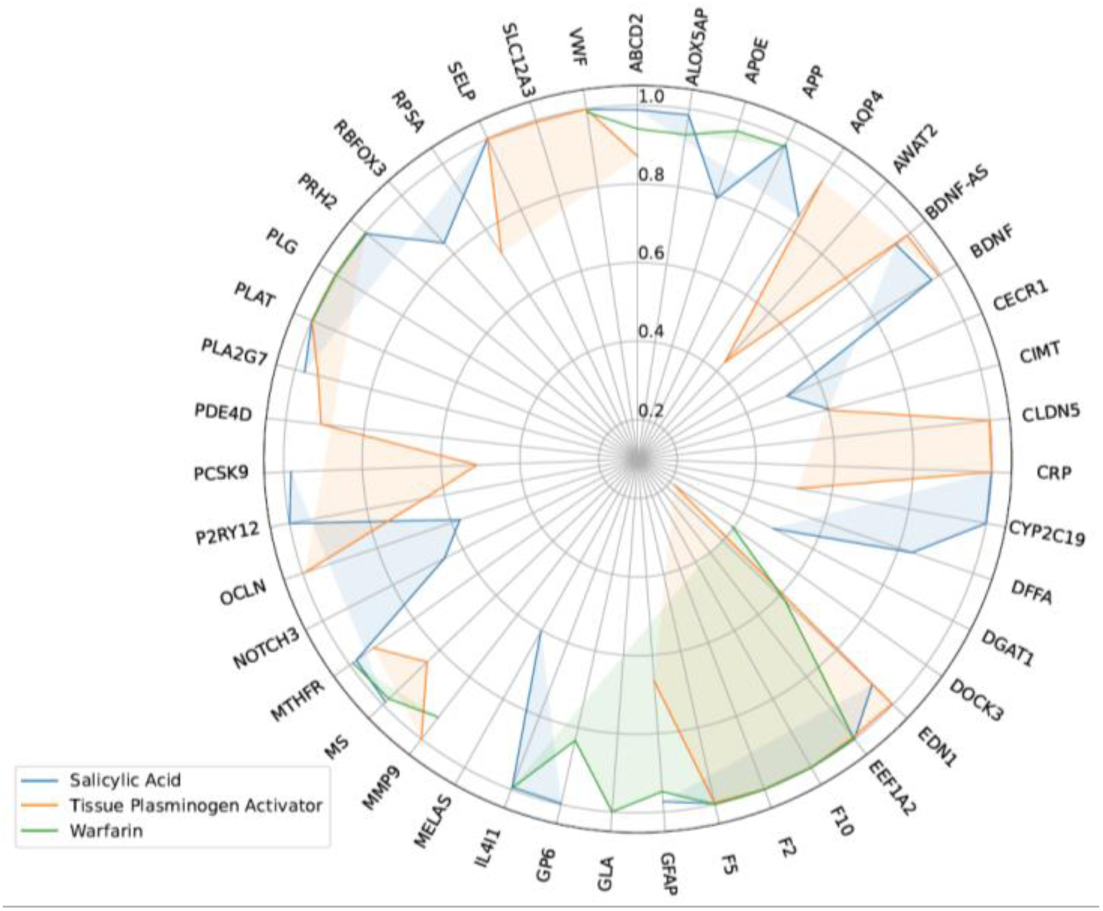
radar chart for p-values between Cerebrovascular accident and affected genes, after consumption of each drug.

## DISCUSSION

In addition to the aforementioned systematic review and network meta-analysis, two additional components were included. The initial step is to identify appropriate medication combinations to use as inputs in the systematic review. To accomplish this, we have applied RL to online databases. Various studies have utilized RL for medical applications (Jafari et al., 2022; Kiaei et al., 2019; Mohammadi et al., 2021). To write prescriptions for various medication combinations, the final step entails searching the internet for relevant information. In light of this, before the systematic review, a native RL model suggested several medication combinations to treat cerebrovascular accidents. In the second phase of the procedure, a systematic review of the recommended medication combinations was conducted to validate the first step’s preliminary findings (Salari et al., 2022a, 2021). In the third phase of this procedure, a network meta-analysis is conducted to determine the effectiveness of the proposed medication combination on human genes and proteins. The prescription drug information for each combination is researched as a final step (such as drug interactions, side effects, drugs, foods completions, etc.)

### Related genes/proteins

Numerous studies and online biological databases have confirmed the relevance of the genes identified as potential targets for the progression of cerebrovascular accidents. According to these studies, cerebrovascular accident disease has led to the involvement of numerous genes and proteins. Among the cases with the lowest p-value, the following can be mentioned: PLAT, PLG, EEF1A2, ABCD2, NOTCH3, P2RY12, F10, DOCK3, CRP, MELAS, MS, AWAT2, CIMT, RPSA, IL4I1, MTHFR, CLDN5, RBFOX3, BDNF, PLA2G7, F5, ALOX5AP, DGAT2, MMP9, APP, VWF, PDE4D, PRH2, DFFA, NINJ2, F2, PCSK9, SLC12A3, GLA, DCX, APOE, CECR1, RNF213, GFAP, AQP4, SELP, SEMA5B, CCM2, EDN1, BDNF-AS, KRIT1, CYP2C19, DGAT1, OCLN, GP6.

### Prescription drug information

Some of the available information on prescription drugs is utilized to investigate drug interactions, drug and food combinations, adverse effects, and concerns regarding serious conditions.

Several reputable websites, including Medscape, WebMD, Drugs, and Drugbank, were consulted during drug interaction research. In every one of these databases, medications were compared side-by-side.

Internet-accessible drug interaction databases, such as Medscape, WebMD, Drugs, and Drugbank, examined the drugs in pairs. Based on the information provided by these databases, we discovered that some drug interactions can occur between pairs of medications.

#### Salicylic Acid and Tissue Plasminogen Activator interactions (Moderate)

There is a correlation between anticoagulants and non-steroidal anti-inflammatory drugs, which both increase the risk of bleeding episodes. Simultaneous use of anticoagulants and over-the-counter NSAIDs may greatly raise the risk for gastrointestinal bleeding. In contrast, concomitant use of anticoagulants and acetaminophen may lead to an elevated risk for general all-site bleeding events. Ibuprofen and other nonsteroidal anti-inflammatory drugs (NSAIDs) are substrates for the CYP2C9 enzyme, which can potentially interfere with the metabolism of S-warfarin and further raise the risk for bleeding associated with warfarin use.(Chan, 1995; Choi et al., 2010; Moore et al., 2015; Teklay et al., 2014)

#### Salicylic Acid and Warfarin interactions (Moderate)

Both anticoagulants and nonsteroidal anti-inflammatory drugs can cause bleeding. Concomitant use of anticoagulants with over-the-counter NSAIDs may raise the risk of gastrointestinal bleeding. In contrast, concomitant use of anticoagulants with acetaminophen may increase the risk for all-site bleeding. NSAIDs such as ibuprofen are substrates of CYP2C9, which may interfere with the metabolism of S-warfarin and increase the risk of warfarin-associated bleeding.(Chan, 1995; Choi et al., 2010; Moore et al., 2015; Teklay et al., 2014)

#### Tissue Plasminogen Activator and Warfarin interactions (Moderate)

Because of how they work, anticoagulant drugs make patients more likely to experience bleeding complications. Likely, the administration of numerous such medications will raise the risk of bleeding while providing the patient with a little benefit. (Levi, 2016)

## CONCLUSION

Cerebrovascular accident is caused by blood clots and ruptured brain blood arteries. Symptoms include vertigo, numbness, weakness on one side of the body, and difficulties speaking, writing, or comprehending language. High blood pressure, older age, smoking, diabetes, high cholesterol, heart disease, atherosclerosis (a buildup of fatty deposits and plaque inside the coronary arteries), and a family history of cerebrovascular accident enhance the risk of cerebrovascular accident. It is also known as a CVA and a stroke. This Systematic Review and Network Meta-Analysis proposes a drug combination consisting of Salicylic Acid, Tissue Plasminogen Activator, and Warfarin for the treatment of Cerebrovascular accidents. *(Strengths and limitations)* The p-value between cerebrovascular accident and human genes associated with this combination has reached 0.003638, indicating that this drug combination is extremely effective. Additional high-quality clinical trials are required to determine the efficacy and safety of other treatments.

## Data Availability

Not applicable

