## Supplementary material for "Recommending Drug Combinations using Reinforcement Learning to target Genes/proteins that cause Stroke: A comprehensive Systematic Review and Network Meta-analysis": v36_sm2.pdf

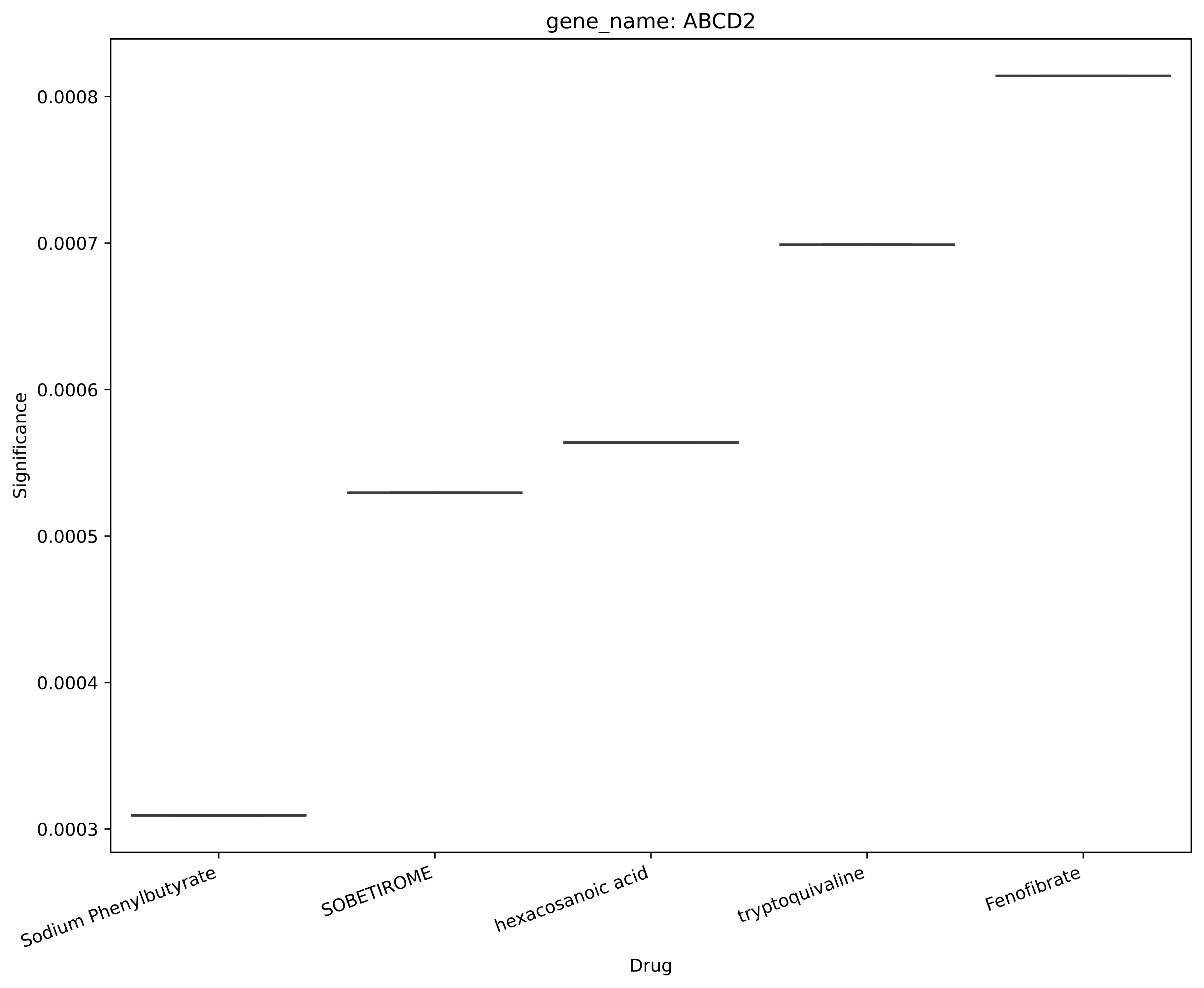
gene_name: ABCD2
Significance
0.0008
0.0007
0.0006
0.0005
0.0004
0.0003
Sodium Phenylbutyrate
SOBETIROME
hexacosanoic acid
tryptoquivaline
Fenofibrate
Drug

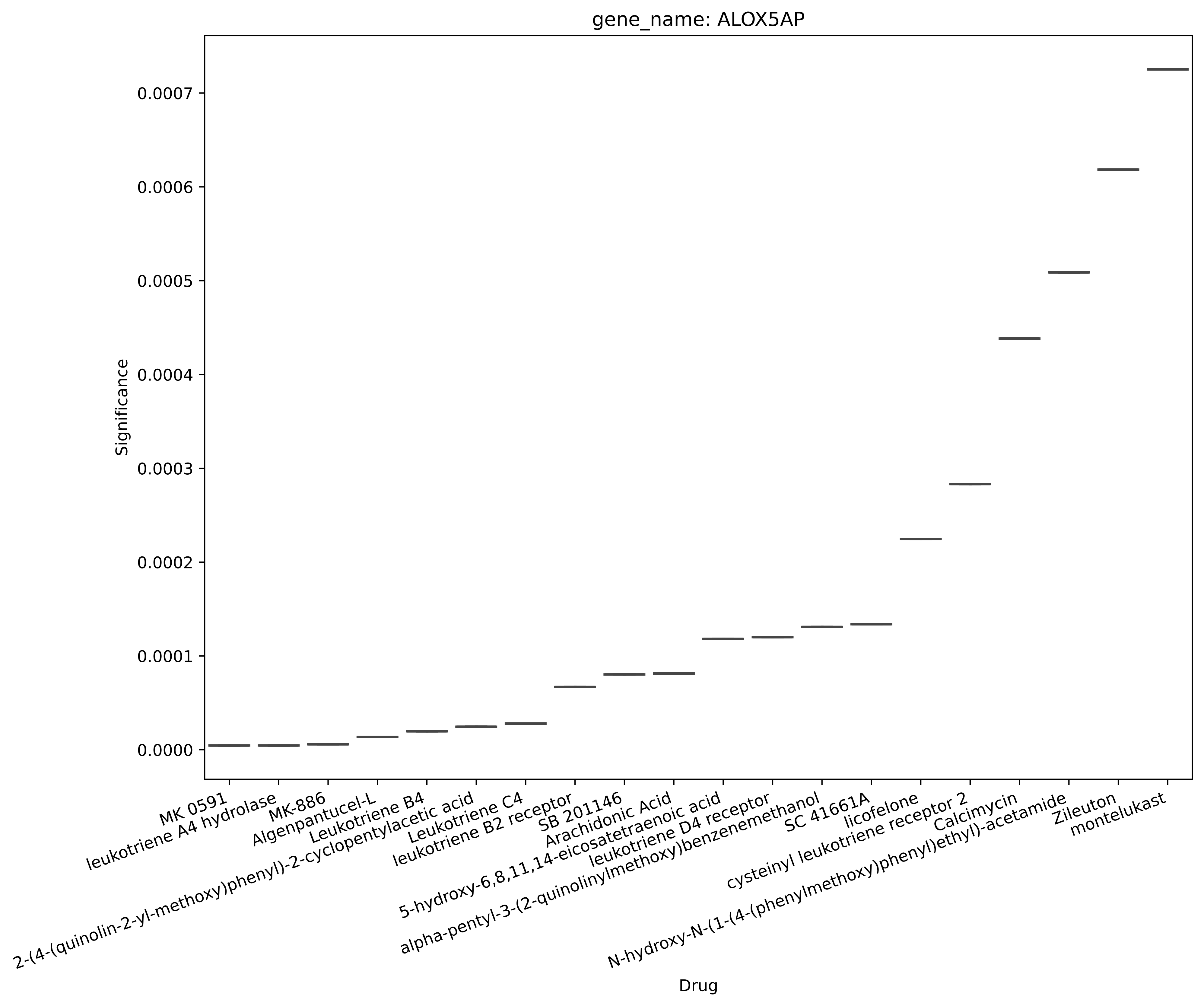
gene_name: ALOX5AP
Significance
0.0007
0.0006
0.0005
0.0004
0.0003
0.0002
0.0001
0.0000
Drug
MK 0591
leukotriene A4 hydrolase
MK-886
Algenpantucel-L
Leukotriene B4
2-(4-(quinolin-2-yl-methoxy)phenyl)-2-cyclopentylacetic acid
Leukotriene C4
leukotriene B2 receptor
SB 201146
5-hydroxy-6,8,11,14-eicosatetraenoic acid
Arachidonic Acid
leukotriene D4 receptor
alpha-pentyl-3-(2-quinolinylmethoxy)benzenemethanol
SC 41661A
licofelone
cysteinyl leukotriene receptor 2
Calcimycin
N-hydroxy-N-(1-(4-(phenylmethoxy)phenyl)ethyl)-acetamide
Zileuton
montelukast

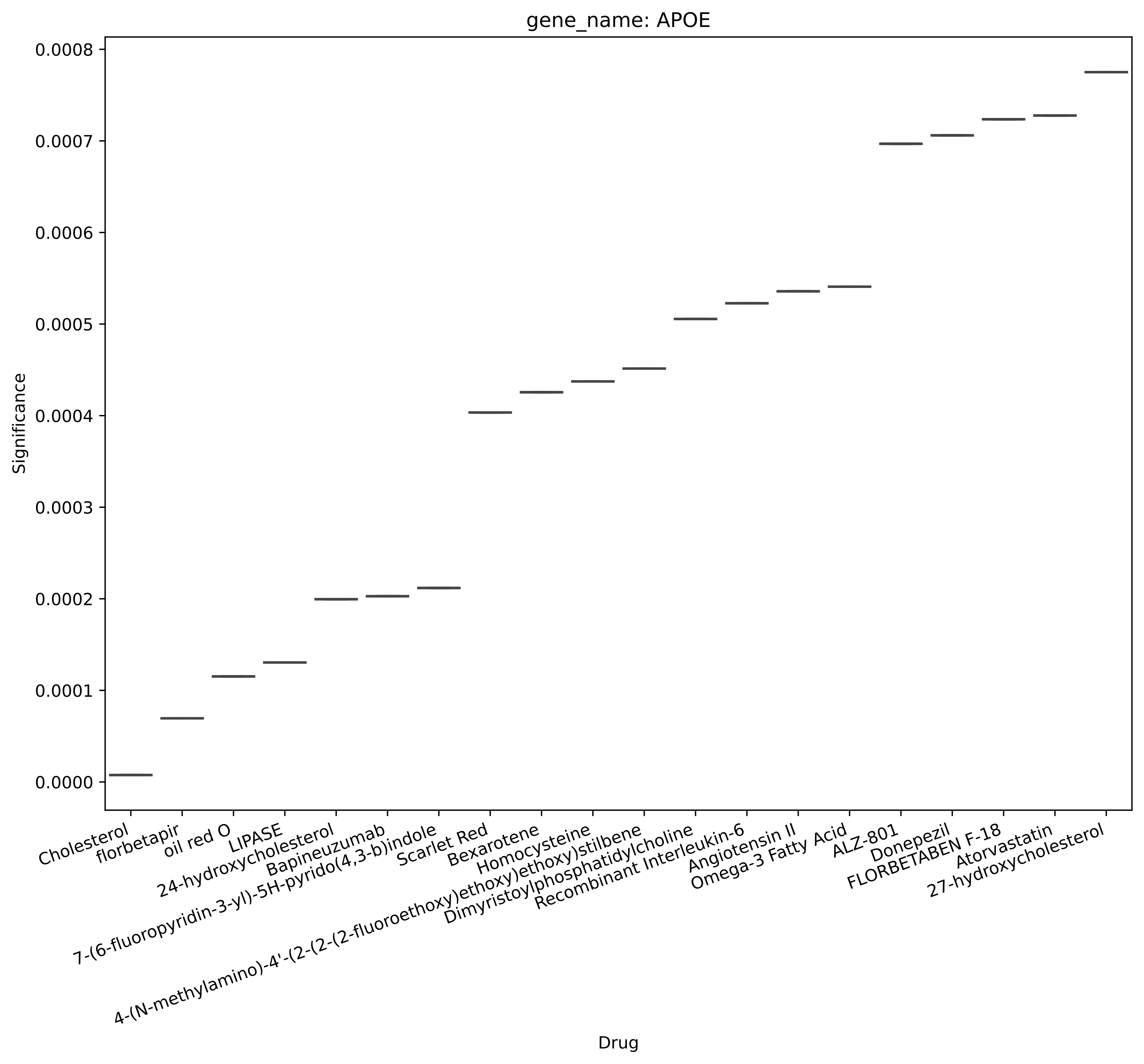

gene_name: APOE
Significance
0.0000
0.0001
0.0002
0.0003
0.0004
0.0005
0.0006
0.0007
0.0008
Cholesterol
florbetapir
oil red O
LIPASE
24-hydroxycholesterol
Bapineuzumab
7-(6-fluoropyridin-3-yl)-5H-pyrido(4,3-b)indole
Scarlet Red
Bexarotene
Homocysteine
4-(N-methylamino)-4'-(2-(2-(2-fluoroethoxy)ethoxy)ethoxy)stilbene
Dimyristoylphosphatidylcholine
Recombinant Interleukin-6
Angiotensin II
Omega-3 Fatty Acid
ALZ-801
Donepezil
FLORBETABEN F-18
Atorvastatin
27-hydroxycholesterol
Drug

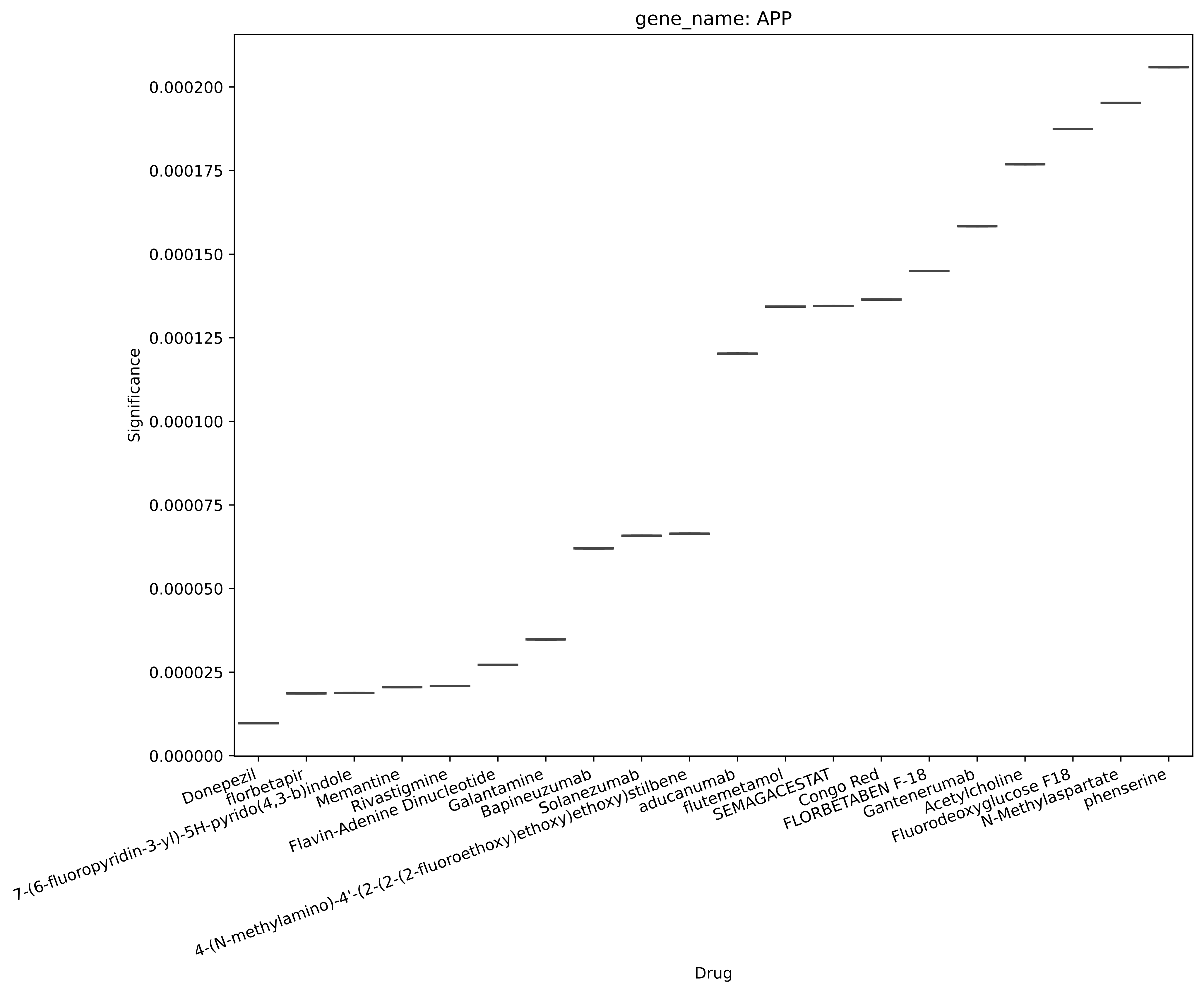
gene_name: APP
Significance
0.000000
0.000025
0.000050
0.000075
0.000100
0.000125
0.000150
0.000175
0.000200
Donepezil
florbetapir
7-(6-fluoropyridin-3-yl)-5H-pyrido(4,3-b)indole
Memantine
Rivastigmine
Flavin-Adenine Dinucleotide
Galantamine
Bapineuzumab
Solanezumab
4-(N-methylamino)-4'-(2-(2-(2-fluoroethoxy)ethoxy)ethoxy)stilbene
aducanumab
flutemetamol
SEMAGACESTAT
Congo Red
FLORBETABEN F-18
Gantenerumab
Acetylcholine
Fluorodeoxyglucose F18
N-Methylaspartate
phenserine
Drug

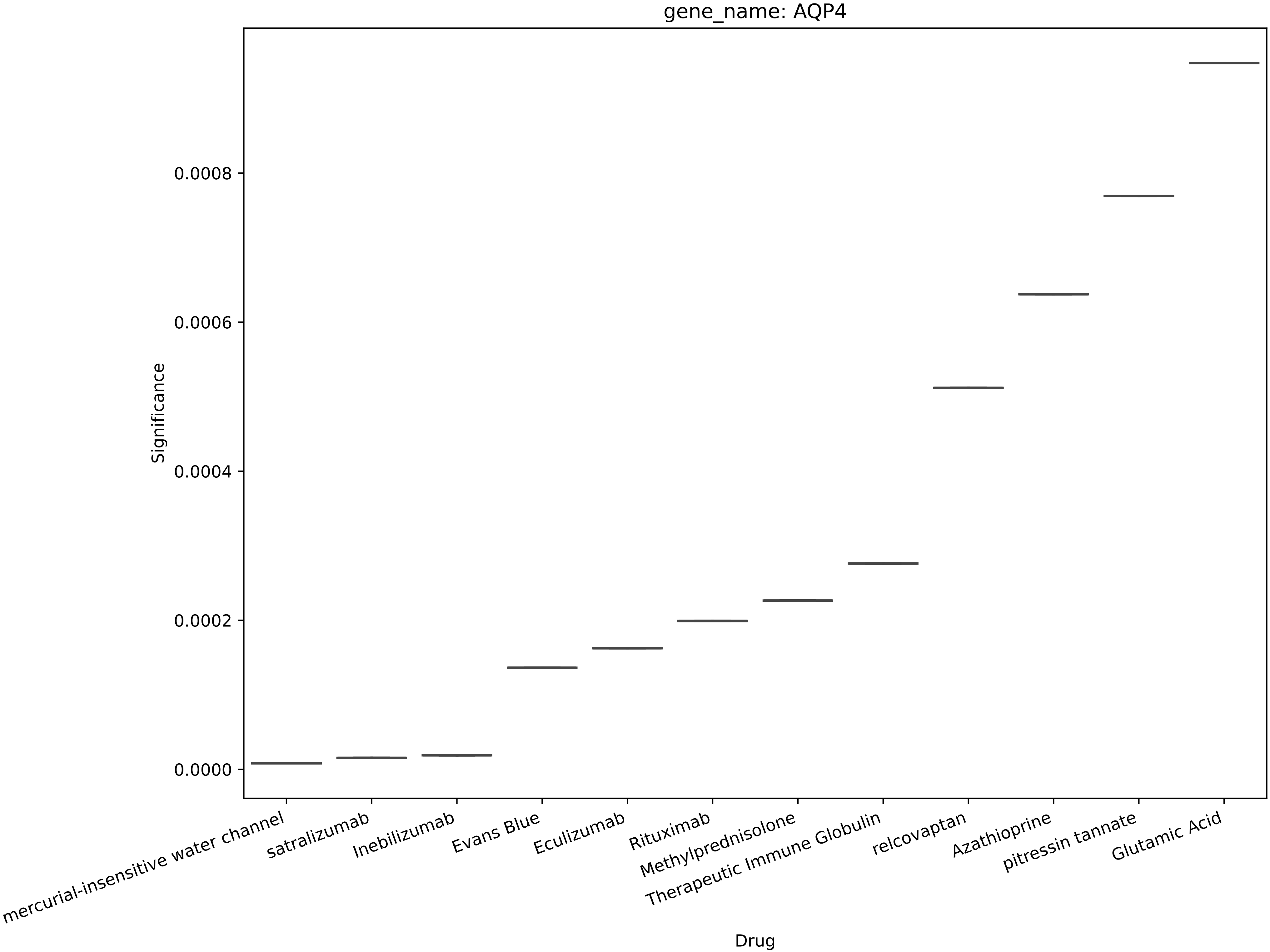
gene_name: AQP4
Significance
0.0000
0.0002
0.0004
0.0006
0.0008
mercurial-insensitive water channel
satralizumab
Inebilizumab
Evans Blue
Eculizumab
Rituximab
Methylprednisolone
Therapeutic Immune Globulin
relcovaptan
Azathioprine
pitressin tannate
Glutamic Acid
Drug

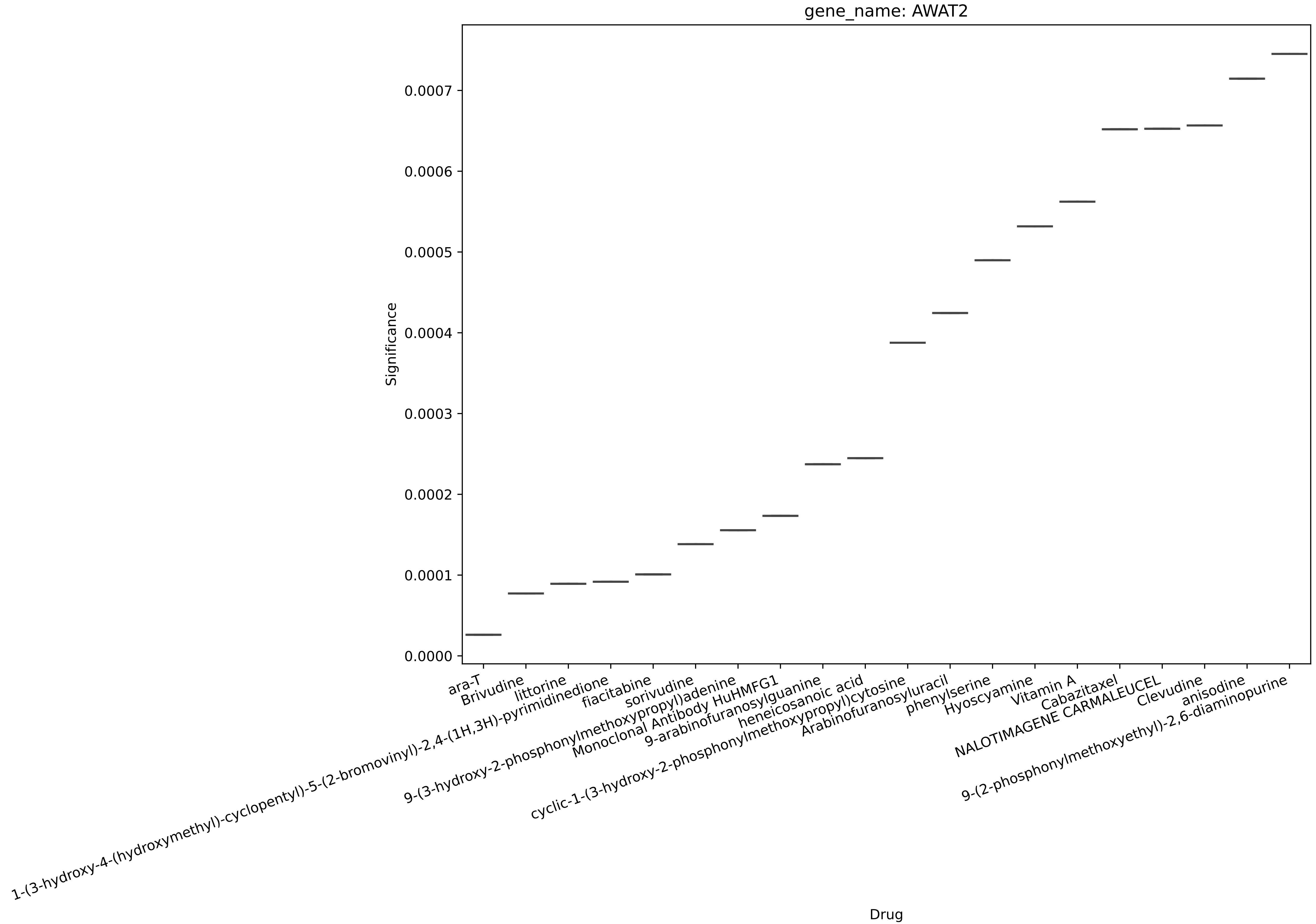

gene_name: AWAT2
Significance
0.0000
0.0001
0.0002
0.0003
0.0004
0.0005
0.0006
0.0007
Drug
ara-T
Brivudine
littorine
1-(3-hydroxy-4-(hydroxymethyl)-cyclopentyl)-5-(2-bromovinyl)-2,4-(1H,3H)-pyrimidinedione
fiacitabine
sorivudine
9-(3-hydroxy-2-phosphonylmethoxypropyl)adenine
Monoclonal Antibody HuHMFG1
9-arabinofuranosylguanine
heneicosanoic acid
cyclic-1-(3-hydroxy-2-phosphonylmethoxypropyl)cytosine
Arabinofuranosyluracil
phenylserine
Hyoscyamine
Vitamin A
Cabazitaxel
NALOTIMAGENE CARMALEUCEL
Clevudine
anisodine
9-(2-phosphonylmethoxyethyl)-2,6-diaminopurine

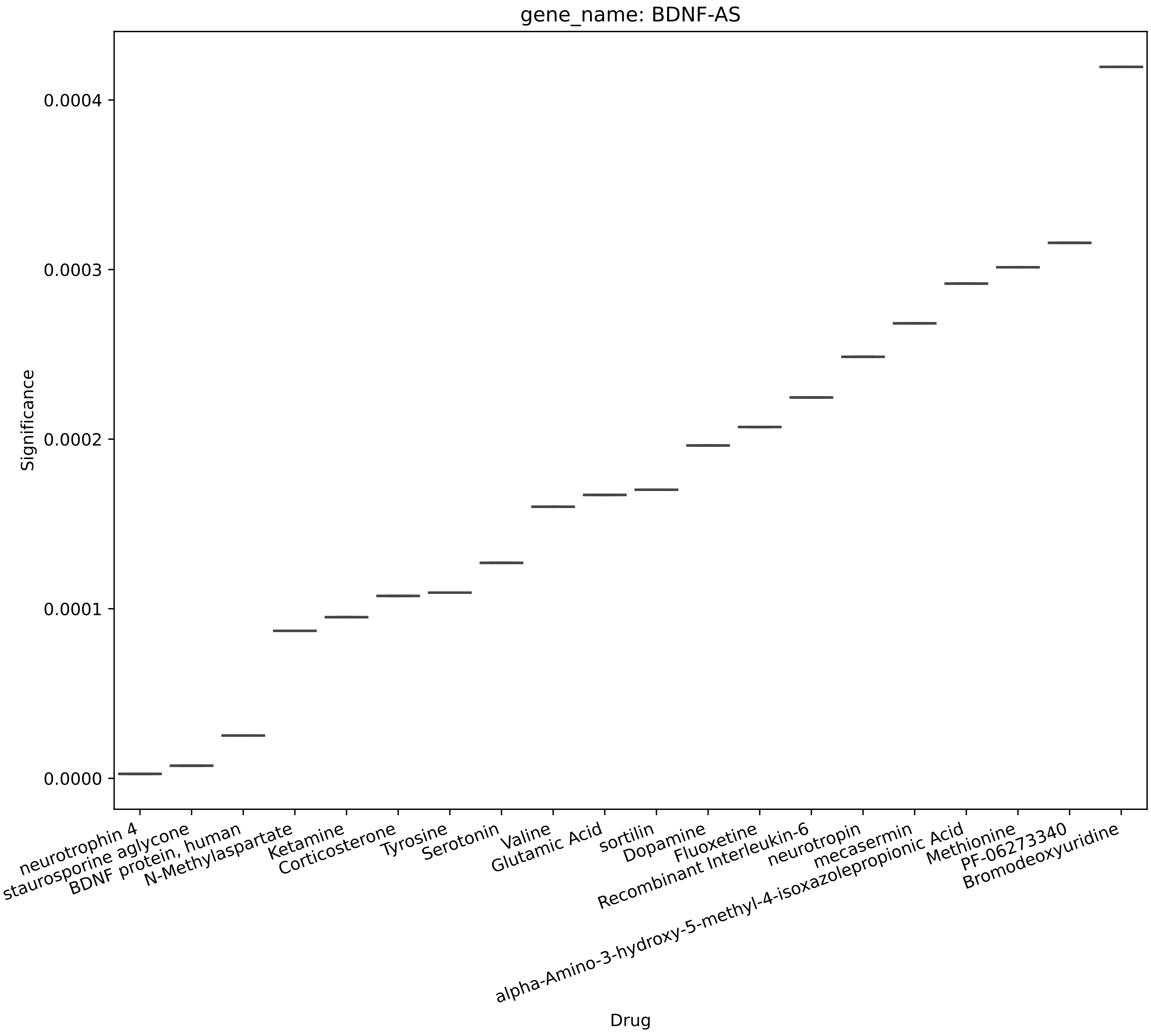
gene_name: BDNF-AS
Significance
0.0000
0.0001
0.0002
0.0003
0.0004
Drug
neurotrophin 4
staurosporine aglycone
BDNF protein, human
N-Methylaspartate
Ketamine
Corticosterone
Tyrosine
Serotonin
Valine
Glutamic Acid
sortilin
Dopamine
Fluoxetine
Recombinant Interleukin-6
neurotropin
alpha-Amino-3-hydroxy-5-methyl-4-isoxazolepropionic Acid
mecasermin
Methionine
PF-06273340
Bromodeoxyuridine

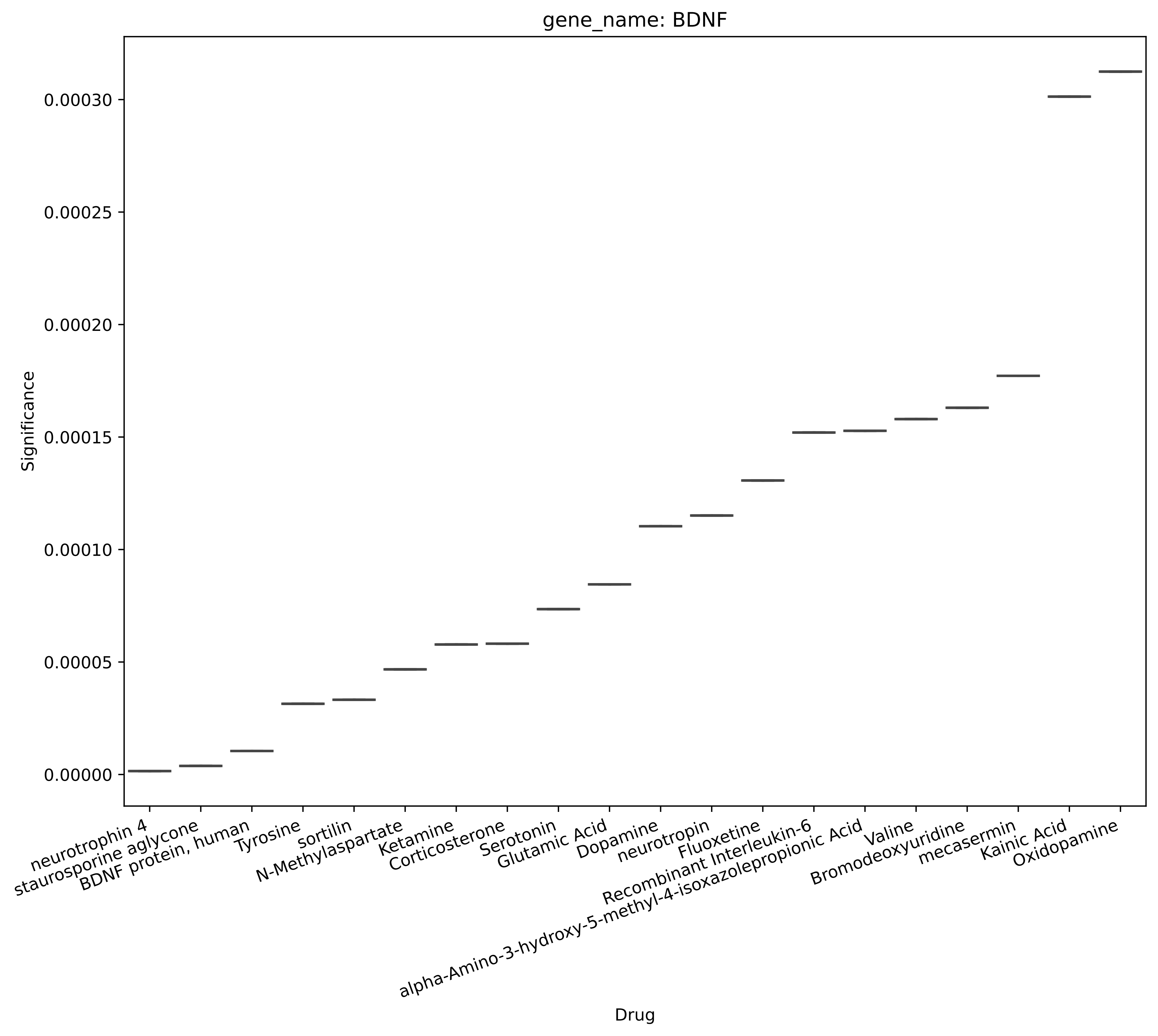
gene_name: BDNF
Significance
0.00030
0.00025
0.00020
0.00015
0.00010
0.00005
0.00000
neurotrophin 4
staurosporine aglycone
BDNF protein, human
Tyrosine
sortilin
N-Methylaspartate
Ketamine
Corticosterone
Serotonin
Glutamic Acid
Dopamine
neurotropin
Fluoxetine
Recombinant Interleukin-6
alpha-Amino-3-hydroxy-5-methyl-4-isoxazolepropionic Acid
Valine
Bromodeoxyuridine
mecasermin
Kainic Acid
Oxidopamine
Drug

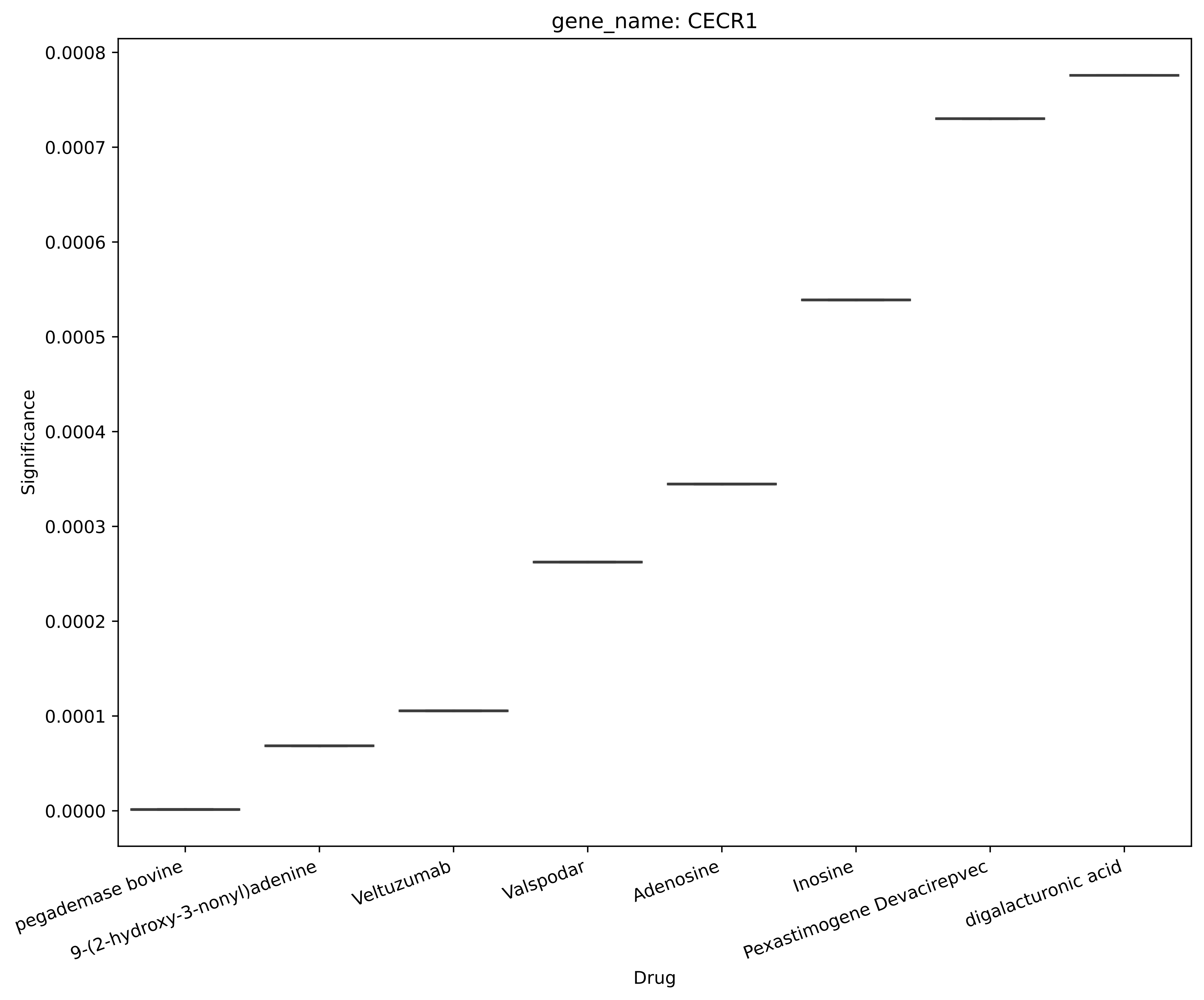
gene_name: CECR1
Significance
0.0008
0.0007
0.0006
0.0005
0.0004
0.0003
0.0002
0.0001
0.0000
pegademase bovine
9-(2-hydroxy-3-nonyl)adenine
Veltuzumab
Valspodar
Adenosine
Inosine
Pexastimogene Devacirepvec
digalacturonic acid
Drug

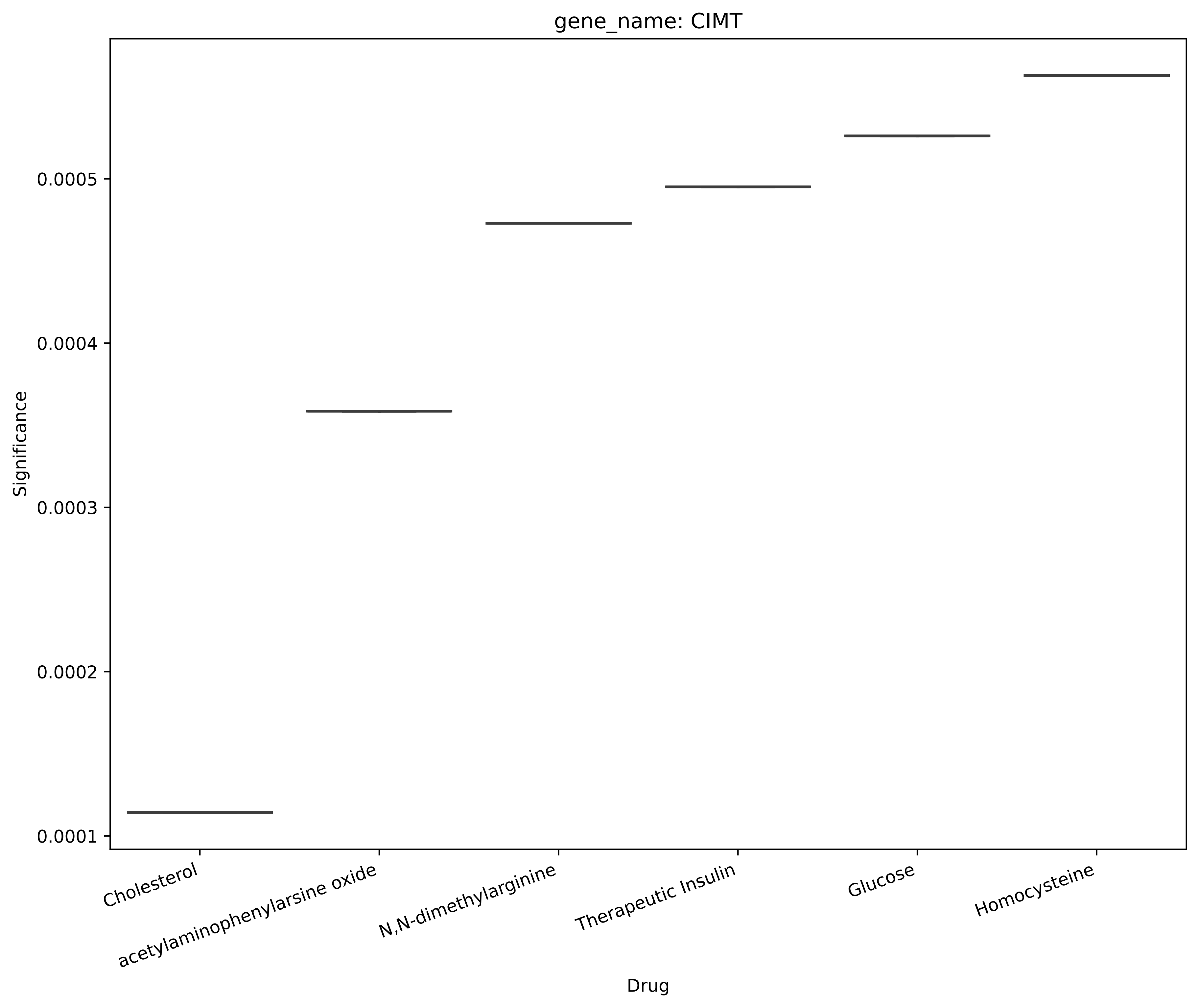
gene_name: CIMT
Significance
0.0005
0.0004
0.0003
0.0002
0.0001
Cholesterol
acetylaminophenylarsine oxide
N,N-dimethylarginine
Therapeutic Insulin
Glucose
Homocysteine
Drug

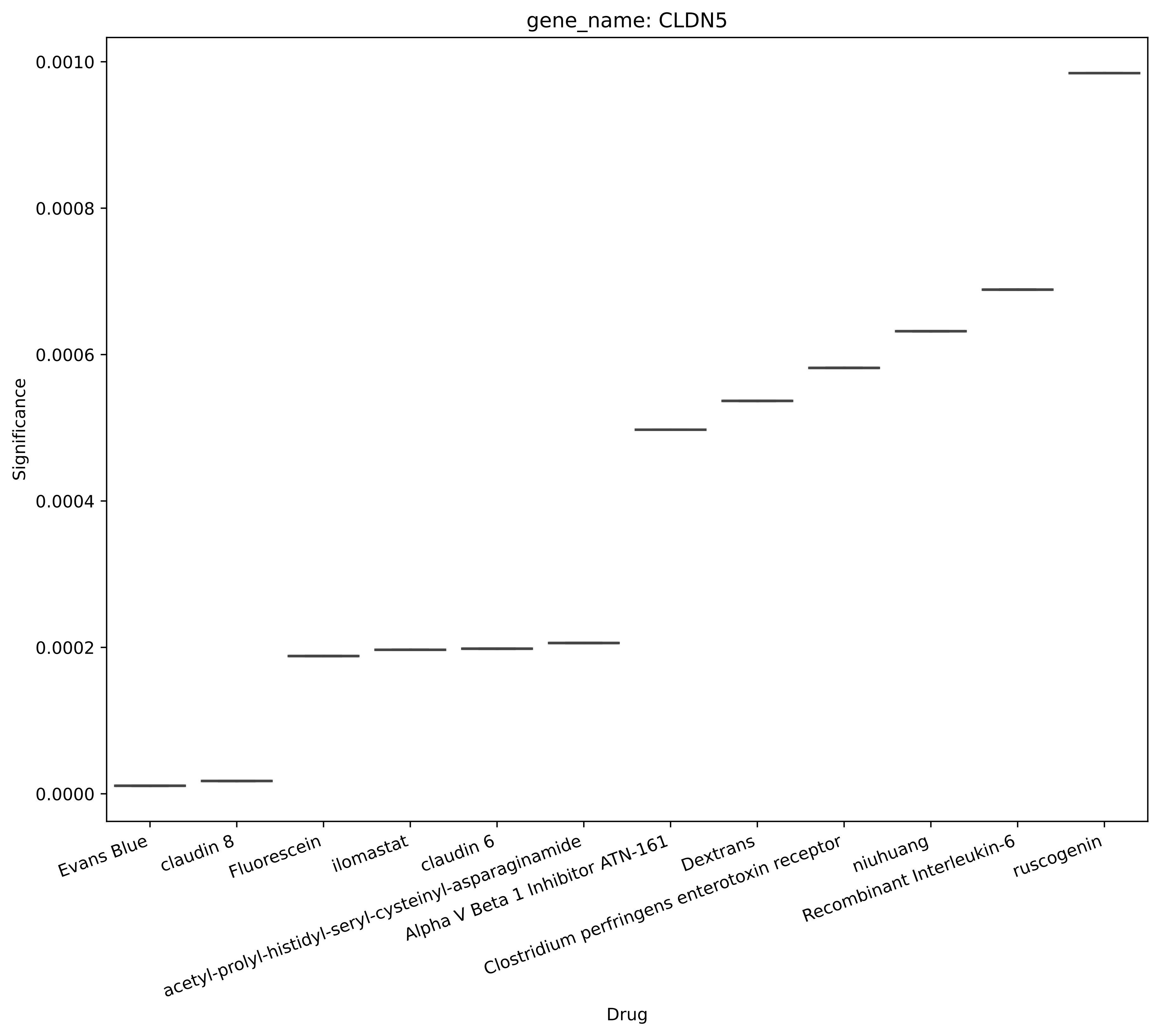
gene_name: CLDN5
Significance
0.0010
0.0008
0.0006
0.0004
0.0002
0.0000
Evans Blue
claudin 8
Fluorescein
ilomastat
claudin 6
acetyl-prolyl-histidyl-seryl-cysteinyl-asparaginamide
Alpha V Beta 1 Inhibitor ATN-161
Dextrans
Clostridium perfringens enterotoxin receptor
niuhuang
Recombinant Interleukin-6
ruscogenin
Drug

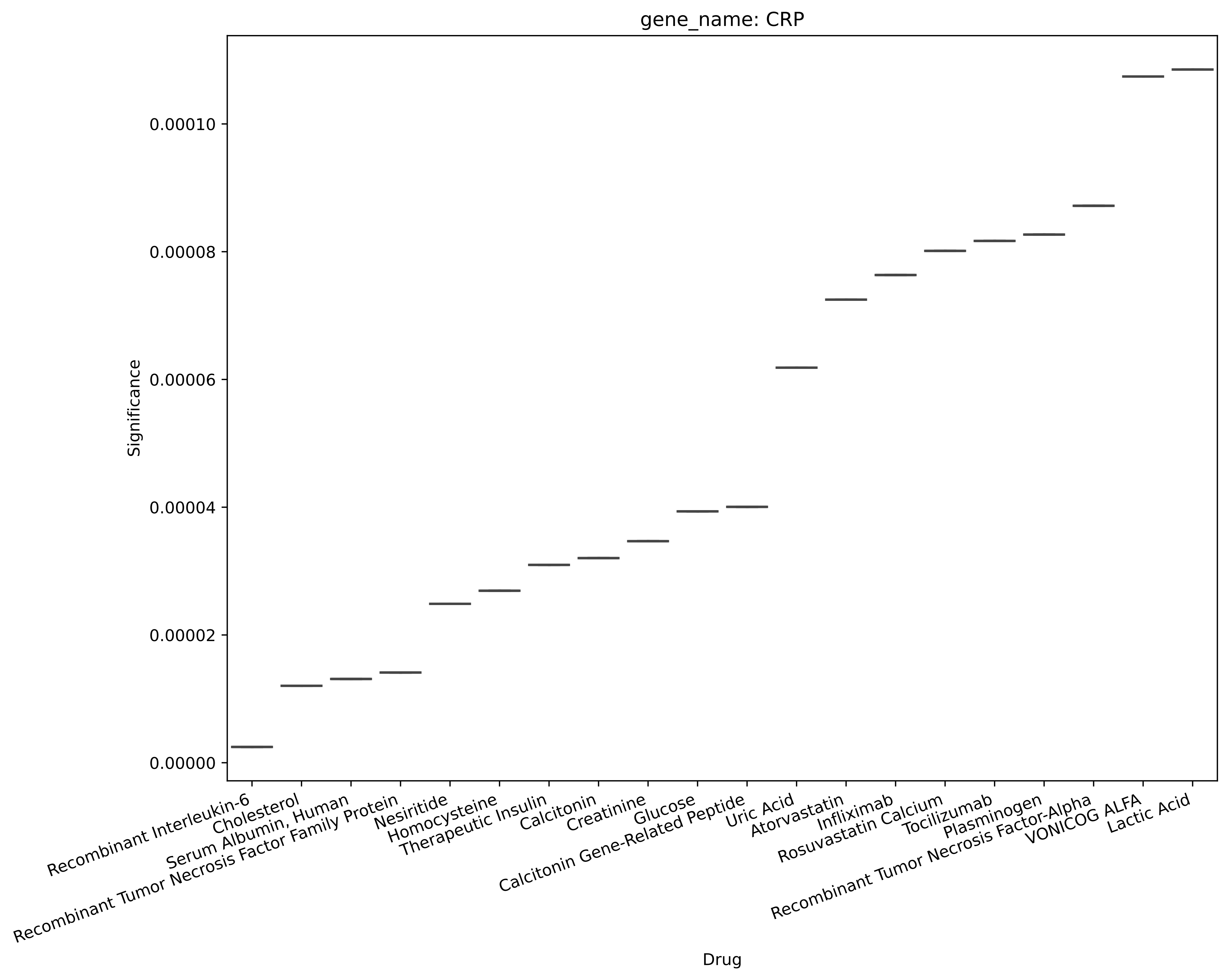
gene_name: CRP
Significance
0.00000
0.00002
0.00004
0.00006
0.00008
0.00010
Recombinant Interleukin-6
Cholesterol
Serum Albumin, Human
Recombinant Tumor Necrosis Factor Family Protein
Nesiritide
Homocysteine
Therapeutic Insulin
Calcitonin
Creatinine
Glucose
Calcitonin Gene-Related Peptide
Uric Acid
Atorvastatin
Infliximab
Rosuvastatin Calcium
Tocilizumab
Plasminogen
Recombinant Tumor Necrosis Factor-Alpha
VONICOG ALFA
Lactic Acid
Drug

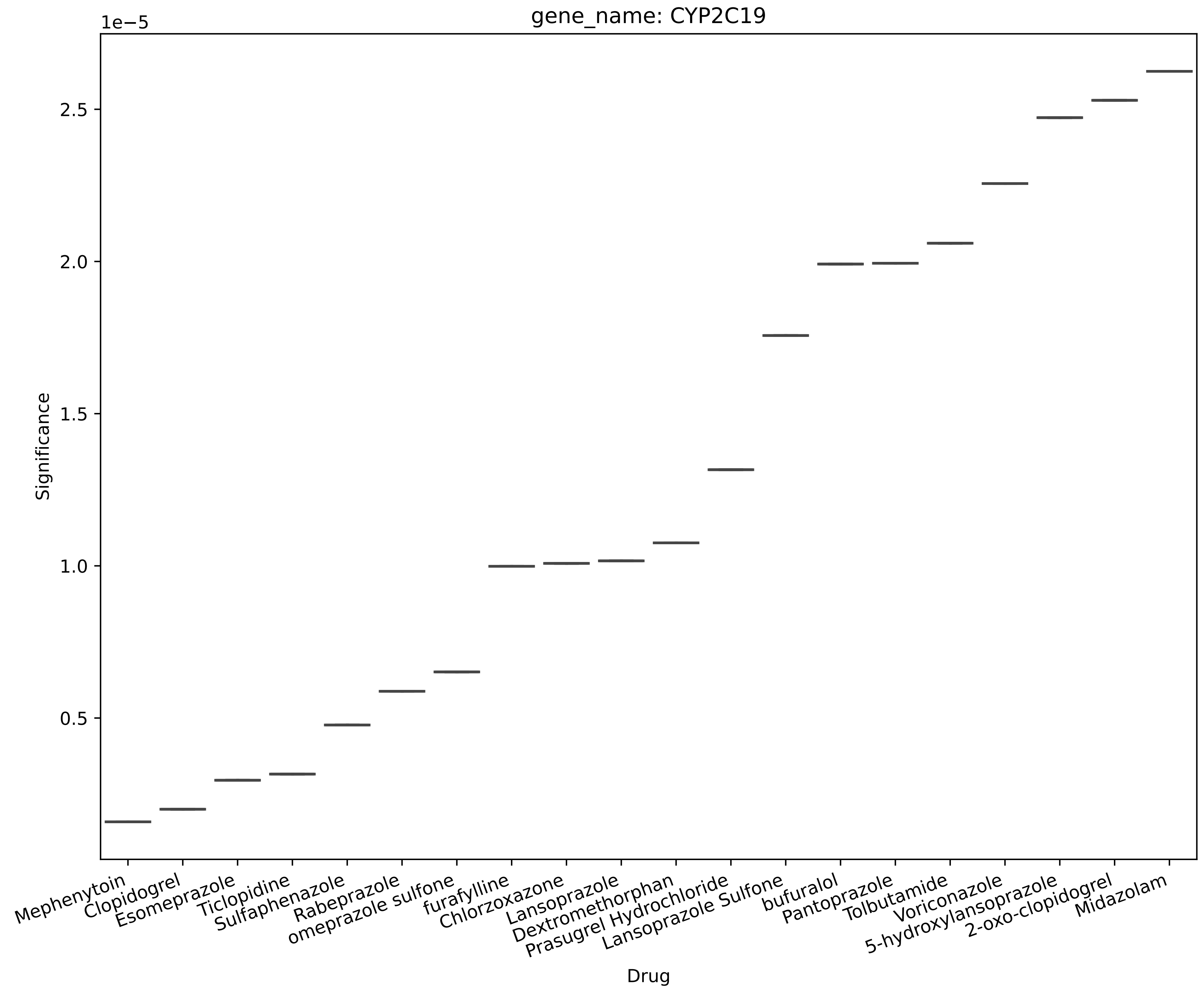

gene_name: CYP2C19
1e−5
2.5
2.0
1.5
1.0
0.5
Significance
Mephenytoin
Clopidogrel
Esomeprazole
Ticlopidine
Sulfaphenazole
Rabeprazole
omeprazole sulfone
furafylline
Chlorzoxazone
Lansoprazole
Dextromethorphan
Prasugrel Hydrochloride
Lansoprazole Sulfone
bufuralol
Pantoprazole
Tolbutamide
Voriconazole
5-hydroxylansoprazole
2-oxo-clopidogrel
Midazolam
Drug

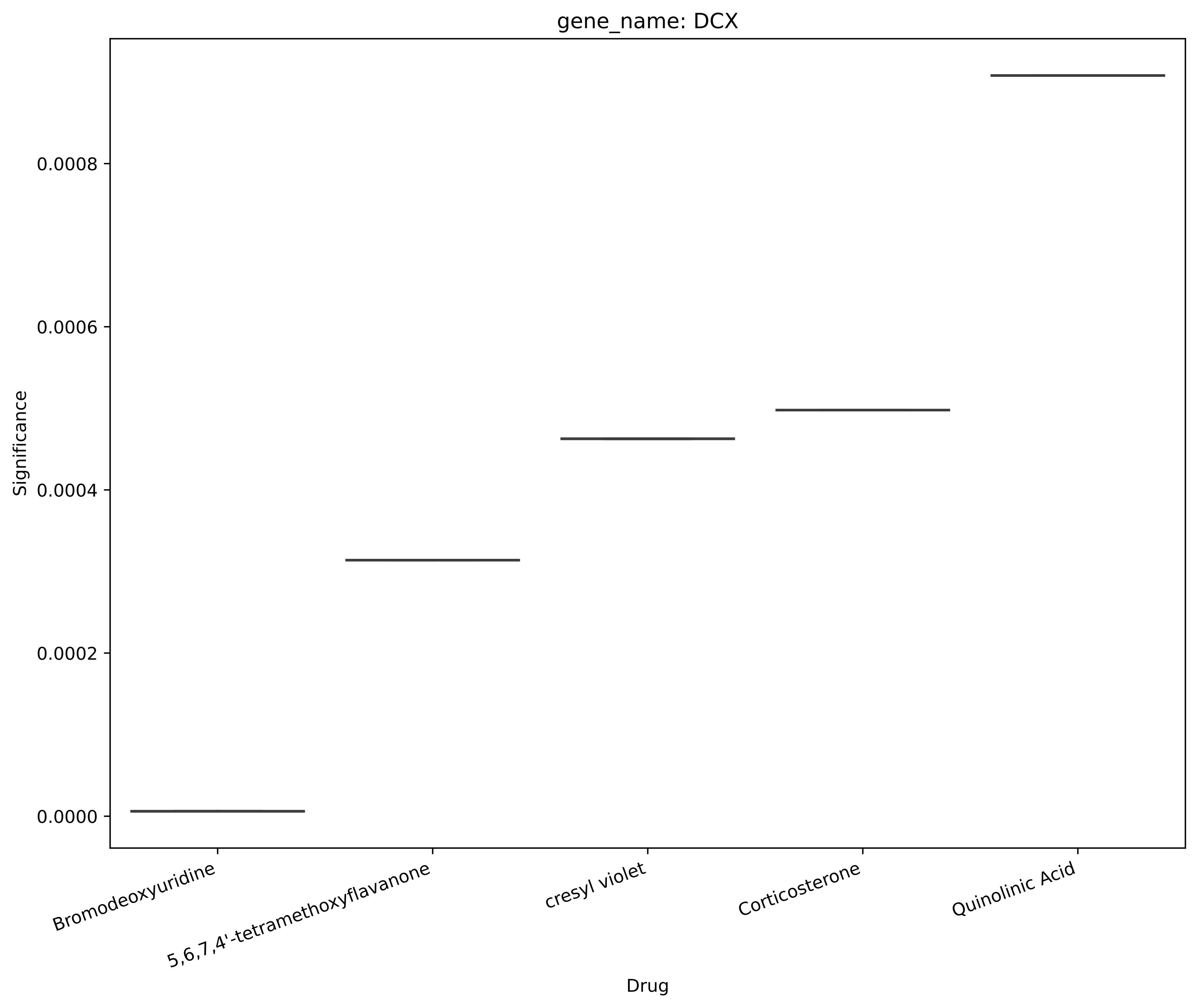

gene_name: DCX
Significance
0.0000
0.0002
0.0004
0.0006
0.0008
Bromodeoxyuridine
5,6,7,4'-tetramethoxyflavanone
cresyl violet
Corticosterone
Quinolinic Acid
Drug

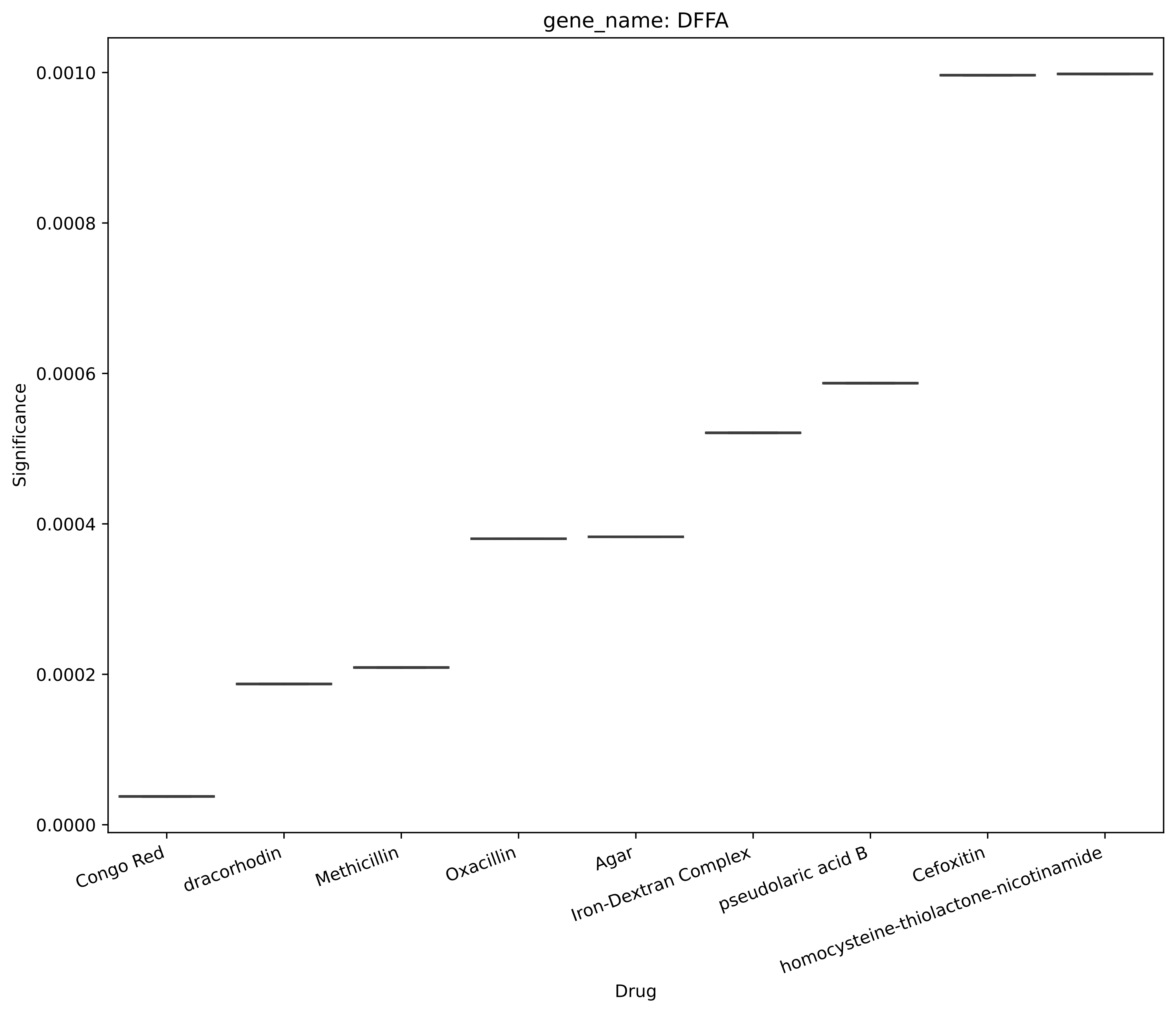
gene_name: DFFA
Significance
0.0010
0.0008
0.0006
0.0004
0.0002
0.0000
Congo Red
dracorhodin
Methicillin
Oxacillin
Agar
Iron-Dextran Complex
pseudolaric acid B
Cefoxitin
homocysteine-thiolactone-nicotinamide
Drug

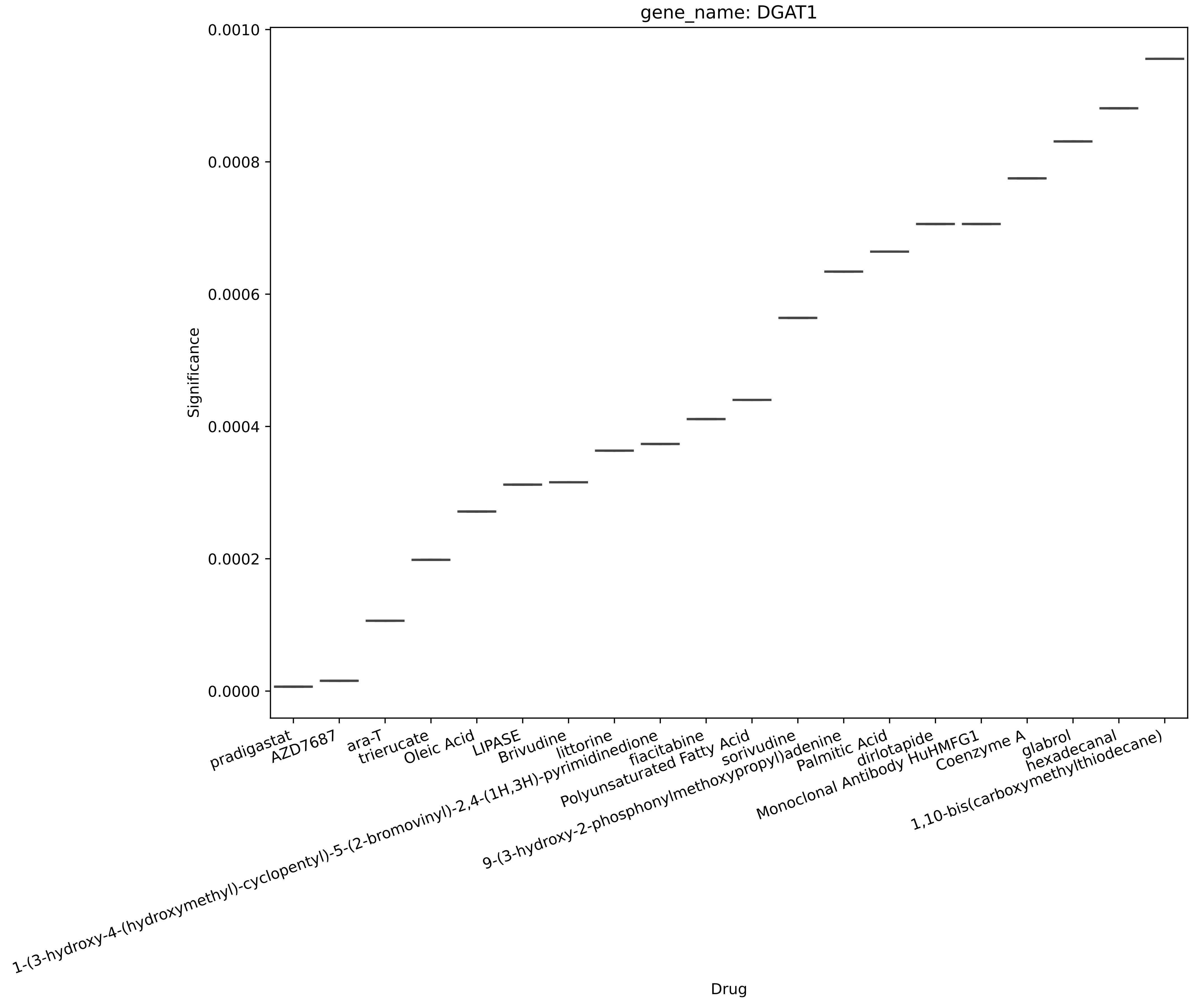

gene_name: DGAT1
0.0010
0.0008
0.0006
0.0004
0.0002
0.0000
Significance
pradigastat
AZD7687
ara-T
trierucate
Oleic Acid
LIPASE
Brivudine
littorine
1-(3-hydroxy-4-(hydroxymethyl)-cyclopentyl)-5-(2-bromovinyl)-2,4-(1H,3H)-pyrimidinedione
fiacitabine
Polyunsaturated Fatty Acid
sorivudine
9-(3-hydroxy-2-phosphonylmethoxypropyl)adenine
Palmitic Acid
dirlotapide
Monoclonal Antibody HuHMFG1
Coenzyme A
glabrol
hexadecanal
1,10-bis(carboxymethylthiodecane)
Drug

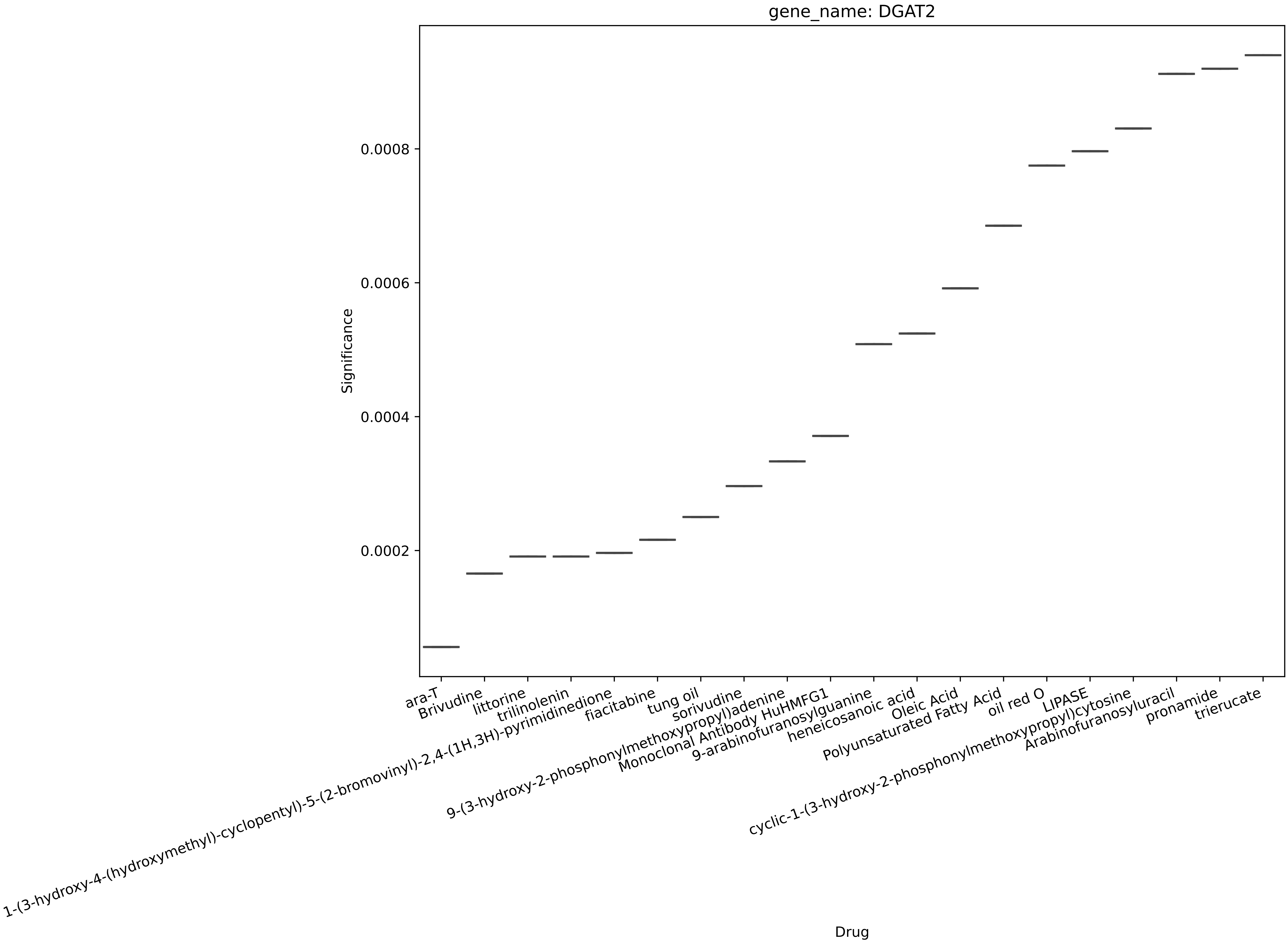
gene_name: DGAT2
Significance
0.0002
0.0004
0.0006
0.0008
Drug
ara-T
Brivudine
littorine
trilinolenin
1-(3-hydroxy-4-(hydroxymethyl)-cyclopentyl)-5-(2-bromovinyl)-2,4-(1H,3H)-pyrimidinedione
fiacitabine
tung oil
sorivudine
9-(3-hydroxy-2-phosphonylmethoxypropyl)adenine
Monoclonal Antibody HuHMFG1
9-arabinofuranosylguanine
heneicosanoic acid
Oleic Acid
Polyunsaturated Fatty Acid
oil red O
LIPASE
cyclic-1-(3-hydroxy-2-phosphonylmethoxypropyl)cytosine
Arabinofuranosyluracil
pronamide
trierucate

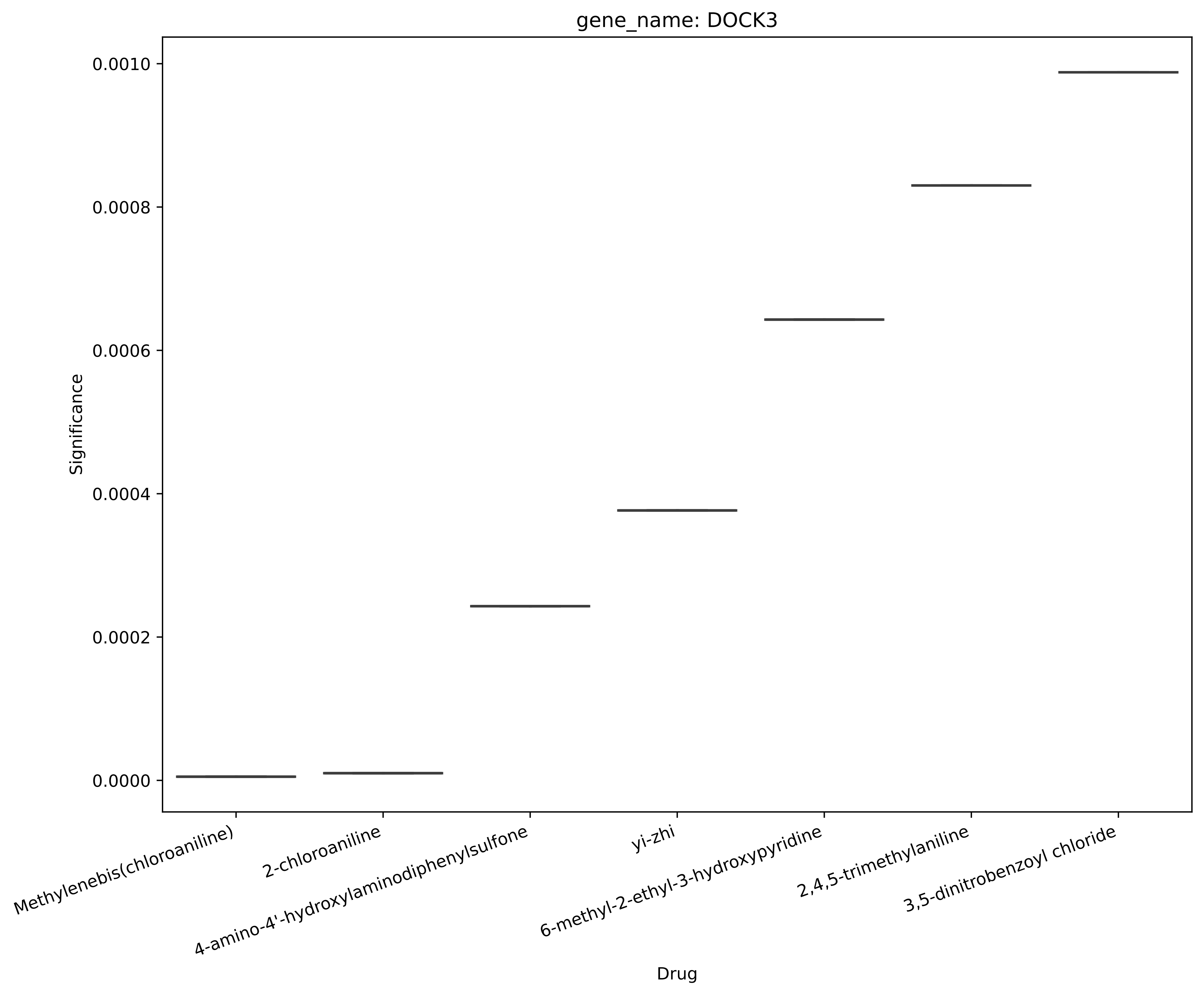
gene_name: DOCK3
Significance
0.0010
0.0008
0.0006
0.0004
0.0002
0.0000
Methylenebis(chloroaniline)
2-chloroaniline
4-amino-4'-hydroxylaminodiphenylsulfone
yi-zhi
6-methyl-2-ethyl-3-hydroxypyridine
2,4,5-trimethylaniline
3,5-dinitrobenzoyl chloride
Drug

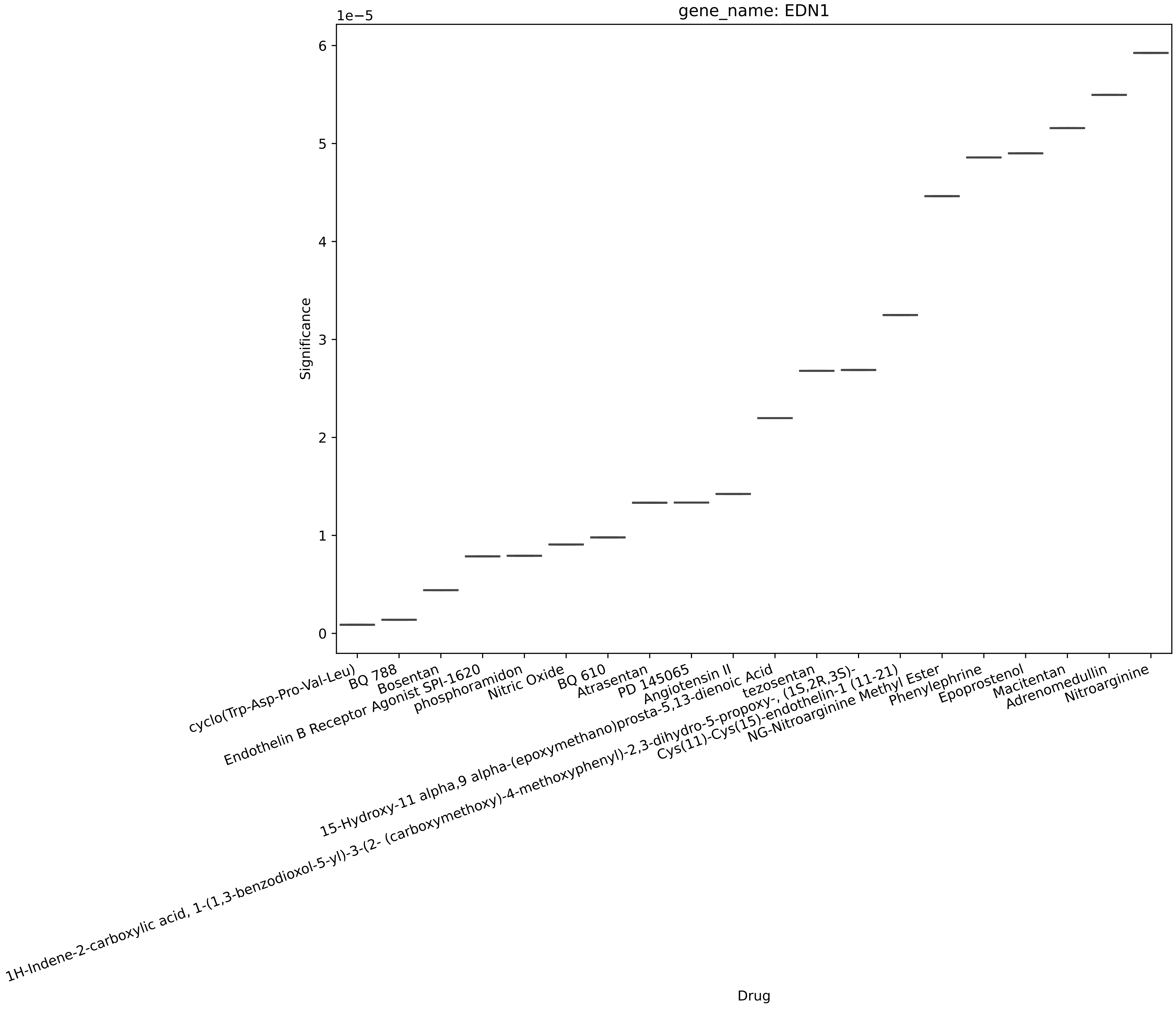
gene_name: EDN1
1e−5
Significance
0
1
2
3
4
5
6
Drug
cyclo(Trp-Asp-Pro-Val-Leu)
BQ 788
Bosentan
Endothelin B Receptor Agonist SPI-1620
phosphoramidon
Nitric Oxide
BQ 610
Atrasentan
PD 145065
Angiotensin II
15-Hydroxy-11 alpha,9 alpha-(epoxymethano)prosta-5,13-dienoic Acid
tezosentan
1H-Indene-2-carboxylic acid, 1-(1,3-benzodioxol-5-yl)-3-(2- (carboxymethoxy)-4-methoxyphenyl)-2,3-dihydro-5-propoxy-, (1S,2R,3S)-
Cys(11)-Cys(15)-endothelin-1 (11-21)
NG-Nitroarginine Methyl Ester
Phenylephrine
Epoprostenol
Macitentan
Adrenomedullin
Nitroarginine

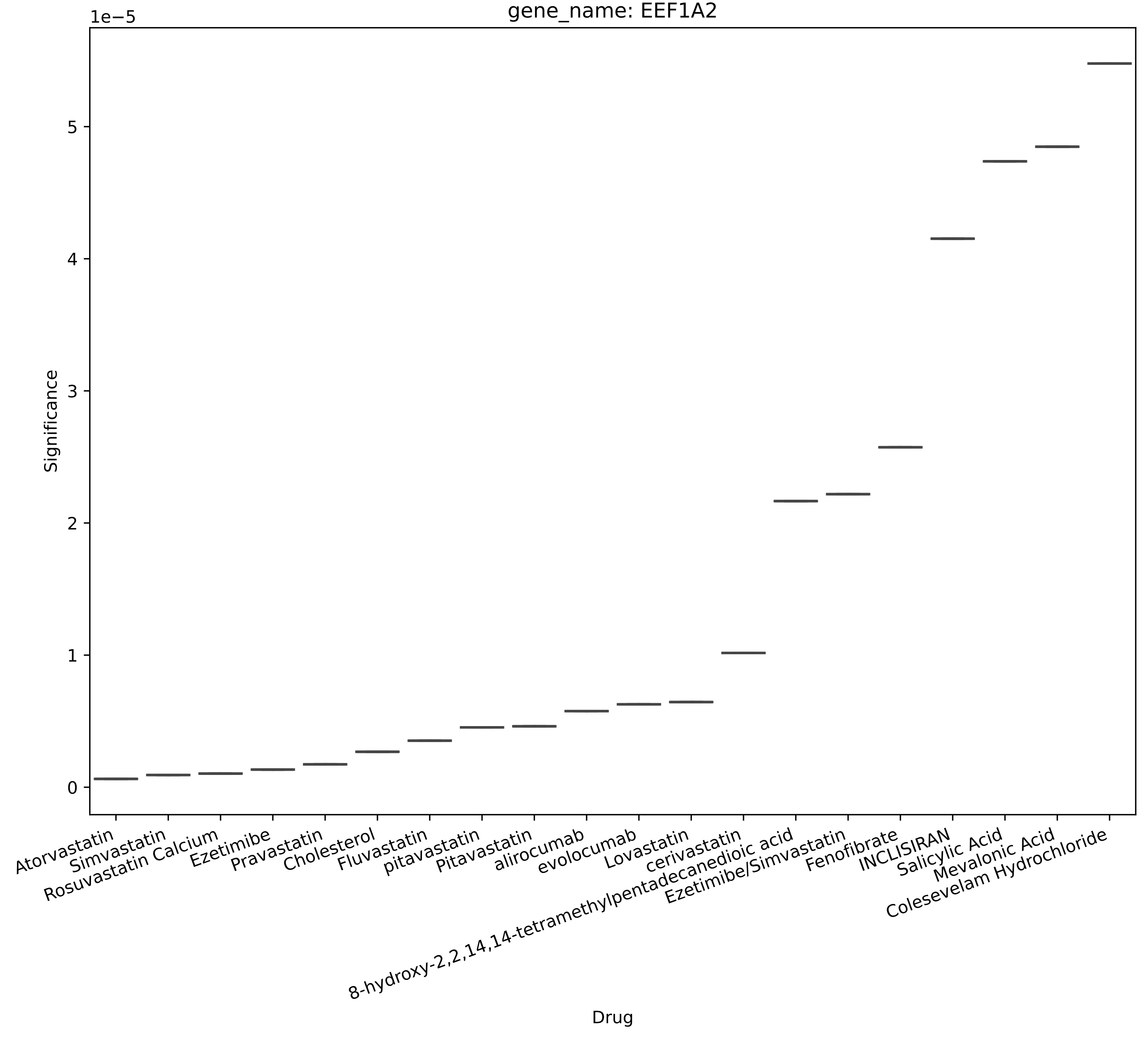

gene_name: EEF1A2
1e−5
Significance
0
1
2
3
4
5
Drug
Atorvastatin
Simvastatin
Rosuvastatin Calcium
Ezetimibe
Pravastatin
Cholesterol
Fluvastatin
pitavastatin
Pitavastatin
alirocumab
evolocumab
Lovastatin
cerivastatin
8-hydroxy-2,2,14,14-tetramethylpentadecanedioic acid
Ezetimibe/Simvastatin
Fenofibrate
INCLISIRAN
Salicylic Acid
Mevalonic Acid
Colesevelam Hydrochloride

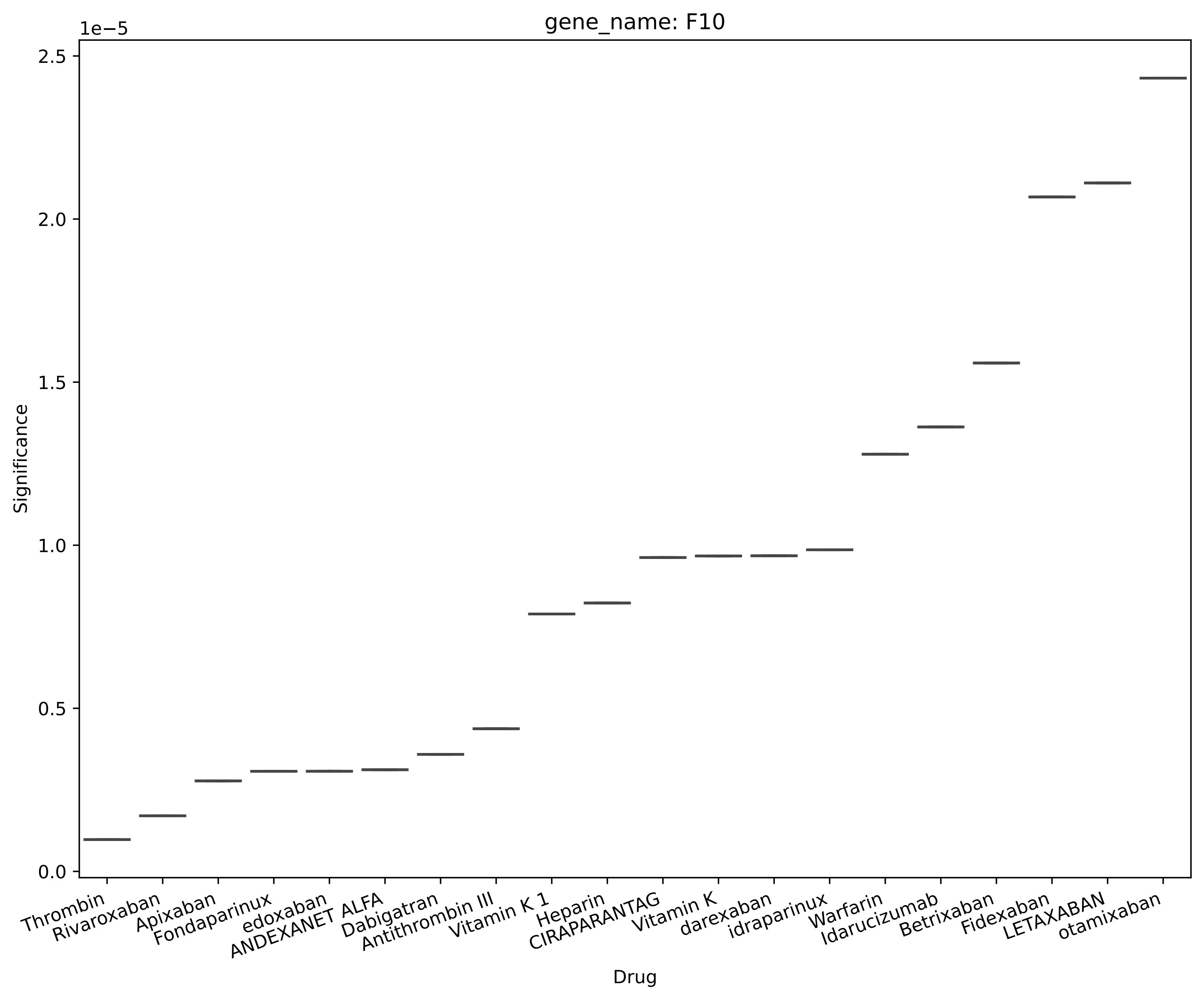
gene_name: F10
1e−5
2.5
2.0
1.5
1.0
0.5
0.0
Significance
Thrombin
Rivaroxaban
Apixaban
Fondaparinux
edoxaban
ANDEXANET ALFA
Dabigatran
Antithrombin III
Vitamin K 1
Heparin
CIRAPARANTAG
Vitamin K
darexaban
idraparinux
Warfarin
Idarucizumab
Betrixaban
Fidexaban
LETAXABAN
otamixaban
Drug

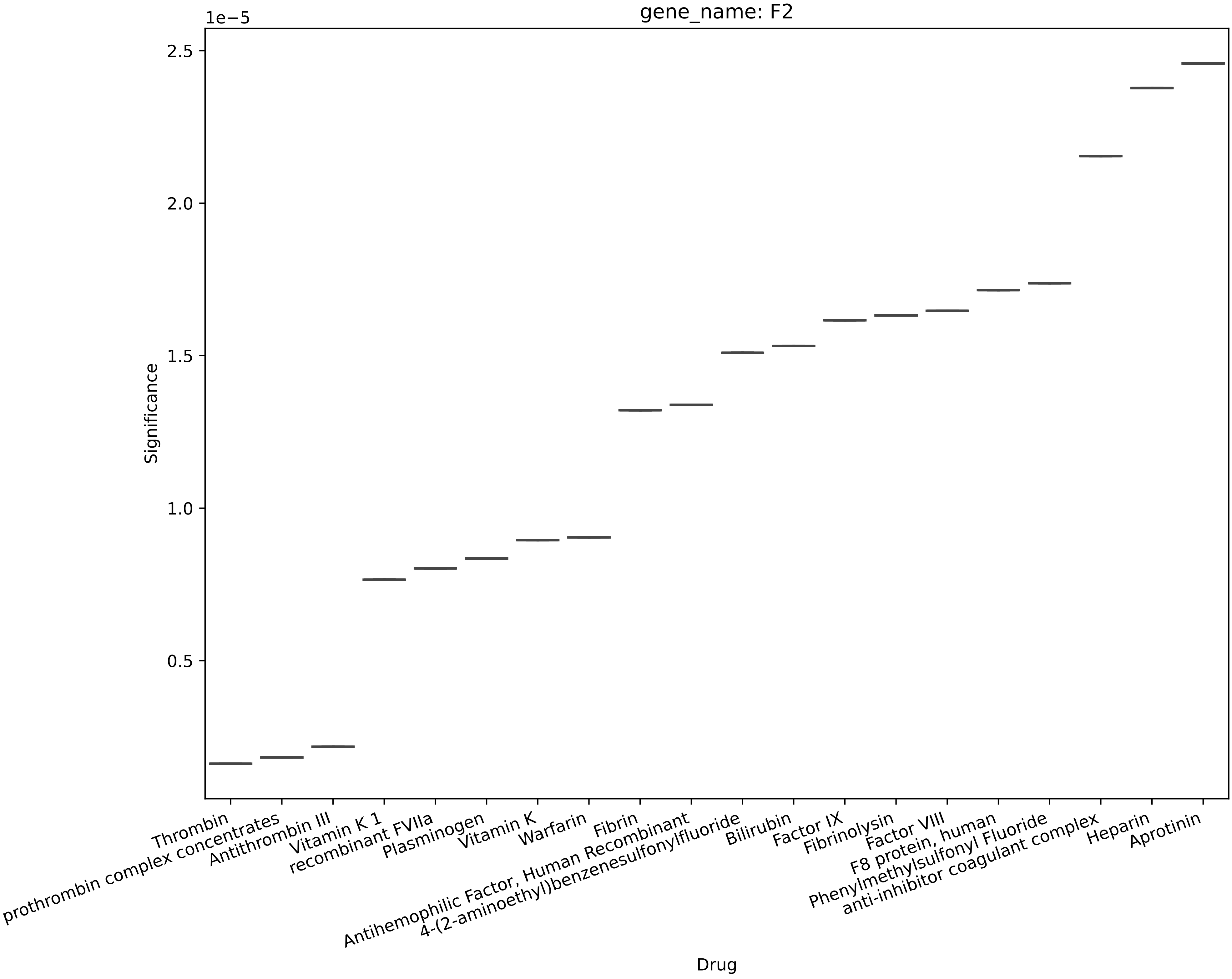

gene_name: F2
1e−5
2.5
2.0
1.5
1.0
0.5
Significance
Thrombin
prothrombin complex concentrates
Antithrombin III
Vitamin K 1
recombinant FVIIa
Plasminogen
Vitamin K
Warfarin
Fibrin
Antihemophilic Factor, Human Recombinant
4-(2-aminoethyl)benzenesulfonylfluoride
Bilirubin
Factor IX
Fibrinolysin
Factor VIII
F8 protein, human
Phenylmethylsulfonyl Fluoride
anti-inhibitor coagulant complex
Heparin
Aprotinin
Drug

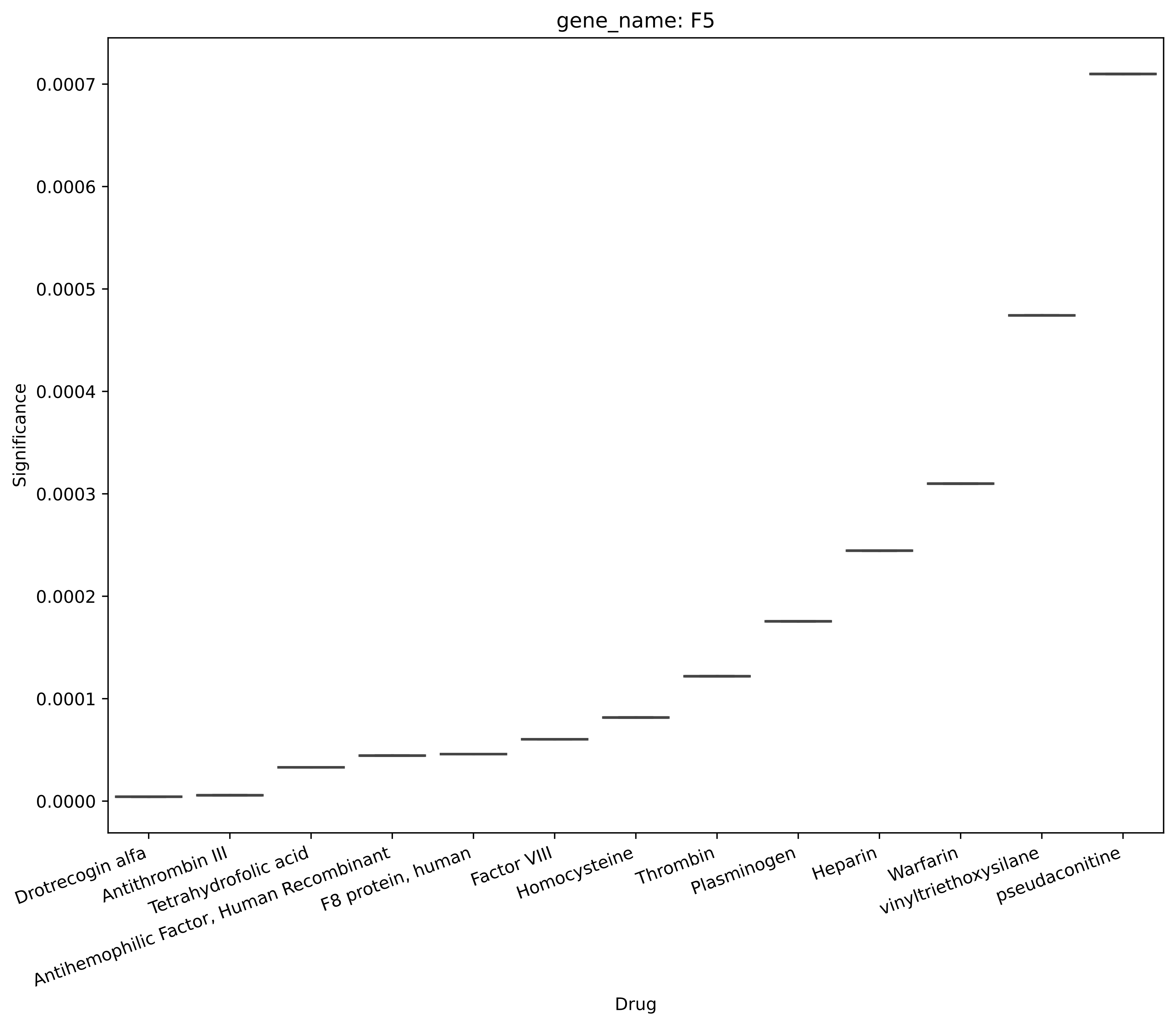
gene_name: F5
Significance
0.0007
0.0006
0.0005
0.0004
0.0003
0.0002
0.0001
0.0000
Drotrecogin alfa
Antithrombin III
Tetrahydrofolic acid
Antihemophilic Factor, Human Recombinant
F8 protein, human
Factor VIII
Homocysteine
Thrombin
Plasminogen
Heparin
Warfarin
vinyltriethoxysilane
pseudaconitine
Drug

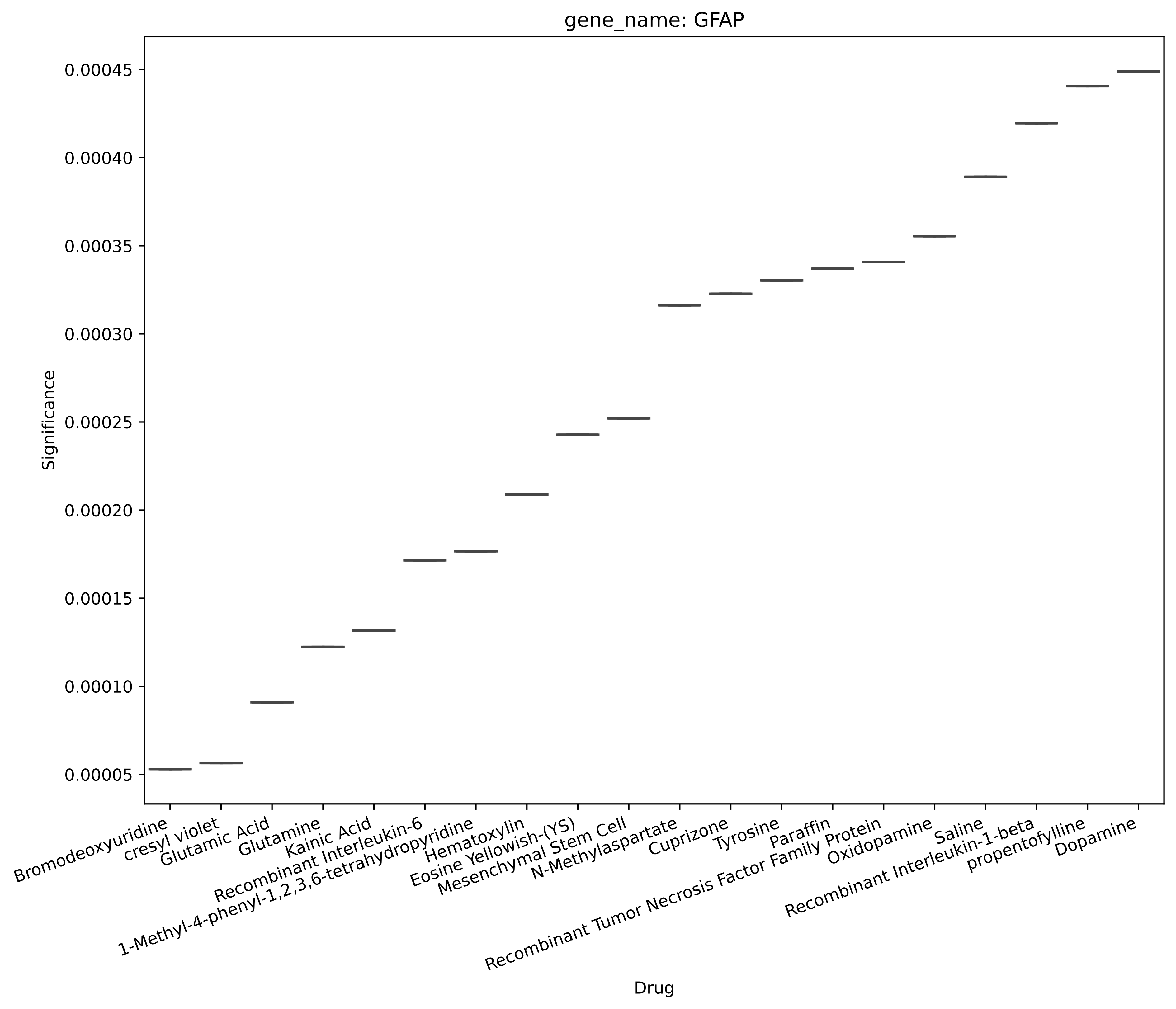
gene_name: GFAP
Significance
0.00005
0.00010
0.00015
0.00020
0.00025
0.00030
0.00035
0.00040
0.00045
Drug
Bromodeoxyuridine
cresyl violet
Glutamic Acid
Glutamine
Kainic Acid
Recombinant Interleukin-6
1-Methyl-4-phenyl-1,2,3,6-tetrahydropyridine
Hematoxylin
Eosine Yellowish-(YS)
Mesenchymal Stem Cell
N-Methylaspartate
Cuprizone
Tyrosine
Paraffin
Recombinant Tumor Necrosis Factor Family Protein
Oxidopamine
Saline
Recombinant Interleukin-1-beta
propentofylline
Dopamine

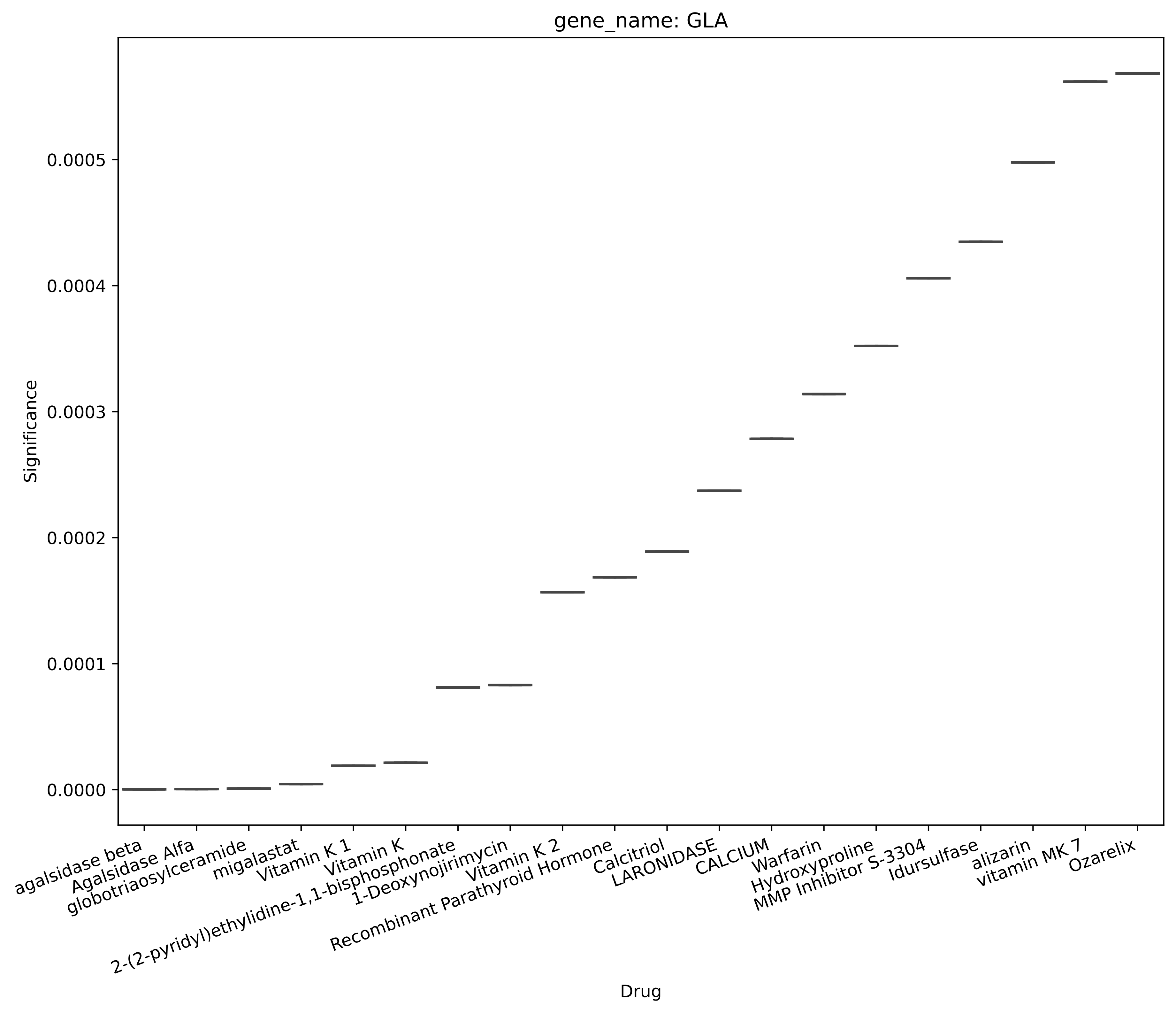
gene_name: GLA
Significance
0.0000
0.0001
0.0002
0.0003
0.0004
0.0005
Drug
agalsidase beta
Agalsidase Alfa
globotriaosylceramide
migalastat
Vitamin K 1
Vitamin K
2-(2-pyridyl)ethylidine-1,1-bisphosphonate
1-Deoxynojirimycin
Vitamin K 2
Recombinant Parathyroid Hormone
Calcitriol
LARONIDASE
CALCIUM
Warfarin
Hydroxyproline
MMP Inhibitor S-3304
Idursulfase
alizarin
vitamin MK 7
Ozarelix

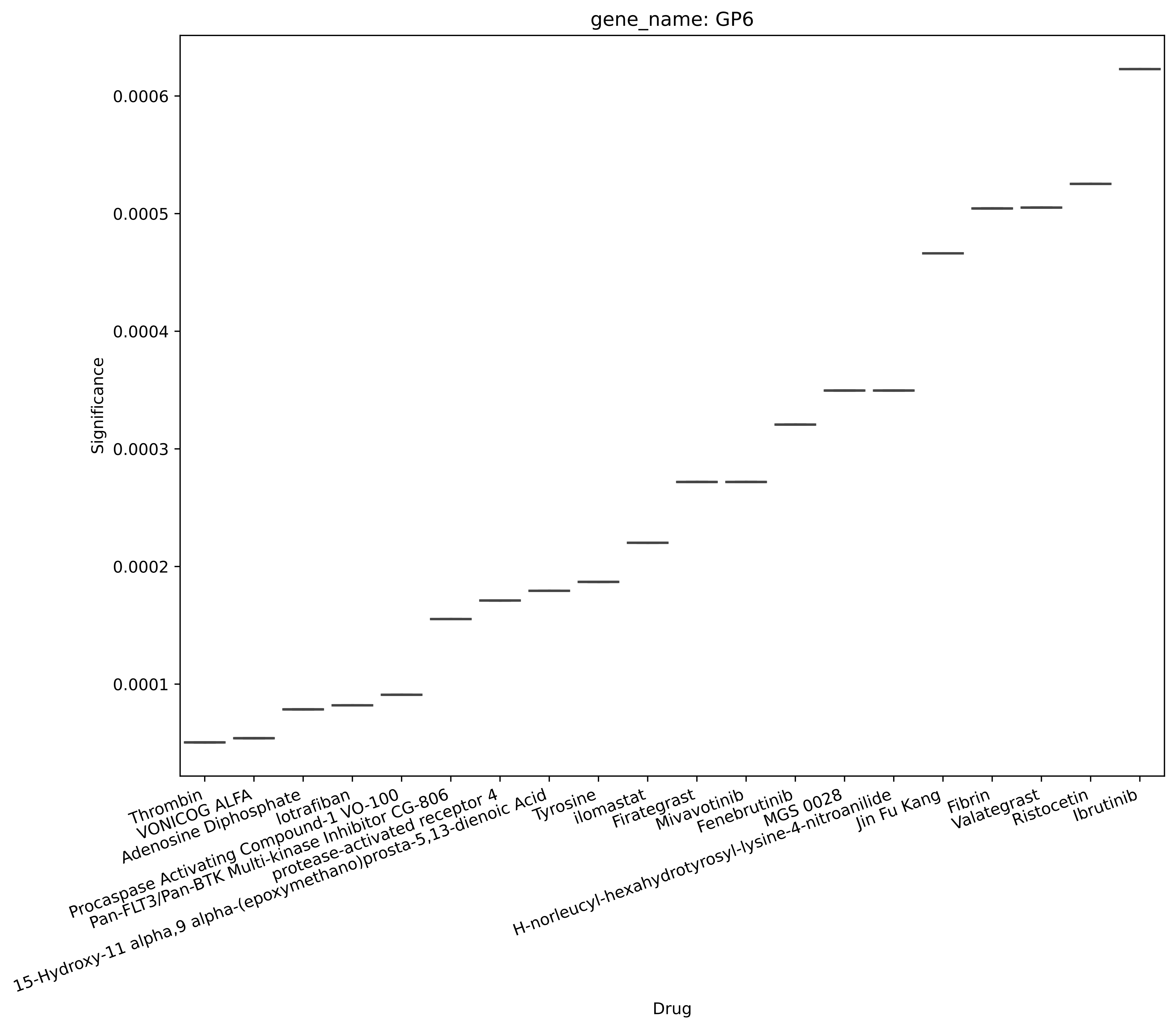

gene_name: GP6
Significance
0.0006
0.0005
0.0004
0.0003
0.0002
0.0001
Thrombin
VONICOG ALFA
Adenosine Diphosphate
lotrafiban
Procaspase Activating Compound-1 VO-100
Pan-FLT3/Pan-BTK Multi-kinase Inhibitor CG-806
protease-activated receptor 4
15-Hydroxy-11 alpha,9 alpha-(epoxymethano)prosta-5,13-dienoic Acid
Tyrosine
ilomastat
Firategrast
Mivavotinib
Fenebrutinib
MGS 0028
H-norleucyl-hexahydrotyrosyl-lysine-4-nitroanilide
Jin Fu Kang
Fibrin
Valategrast
Ristocetin
Ibrutinib
Drug

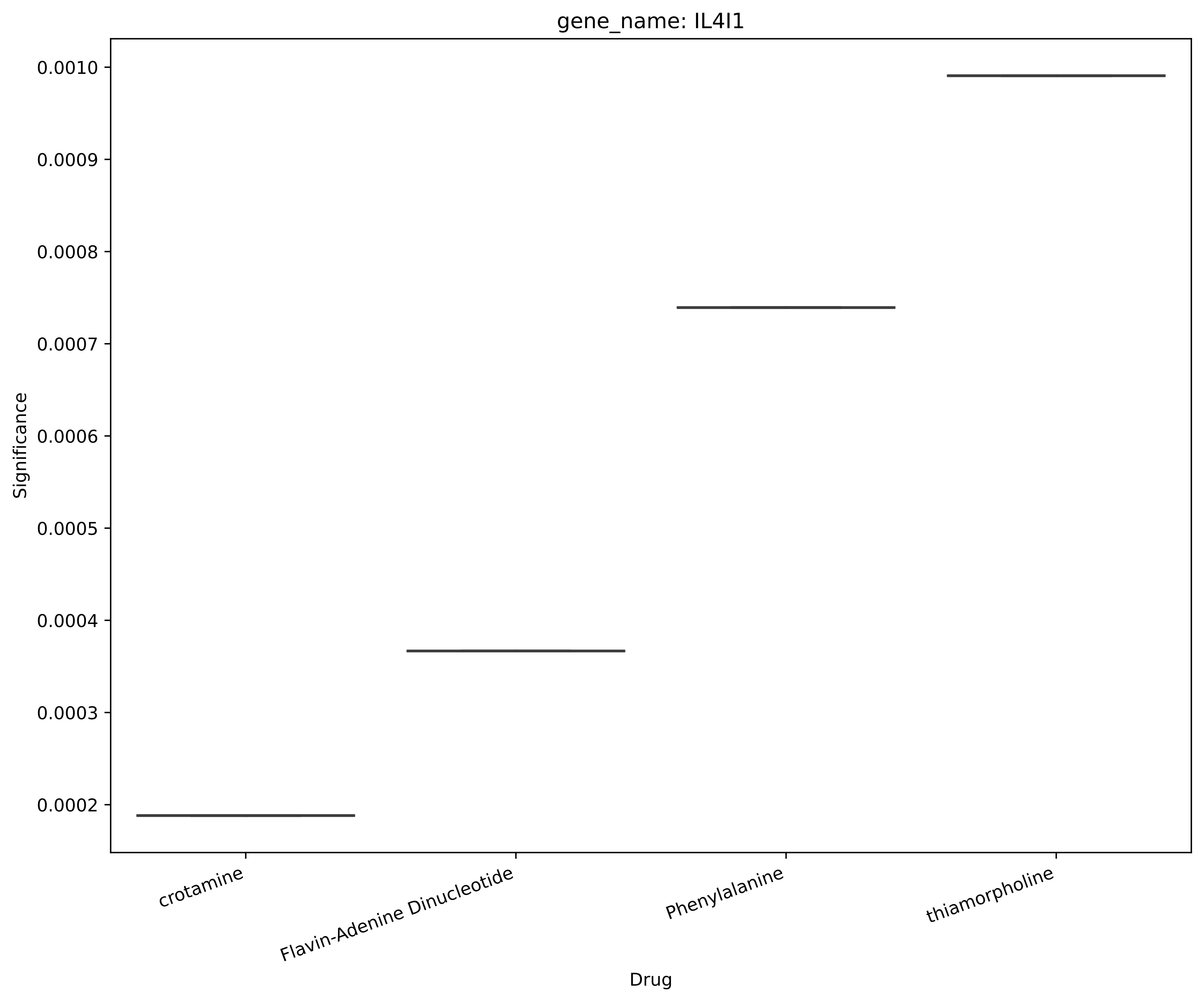
gene_name: IL4I1
Significance
0.0010
0.0009
0.0008
0.0007
0.0006
0.0005
0.0004
0.0003
0.0002
crotamine
Flavin-Adenine Dinucleotide
Phenylalanine
thiamorpholine
Drug

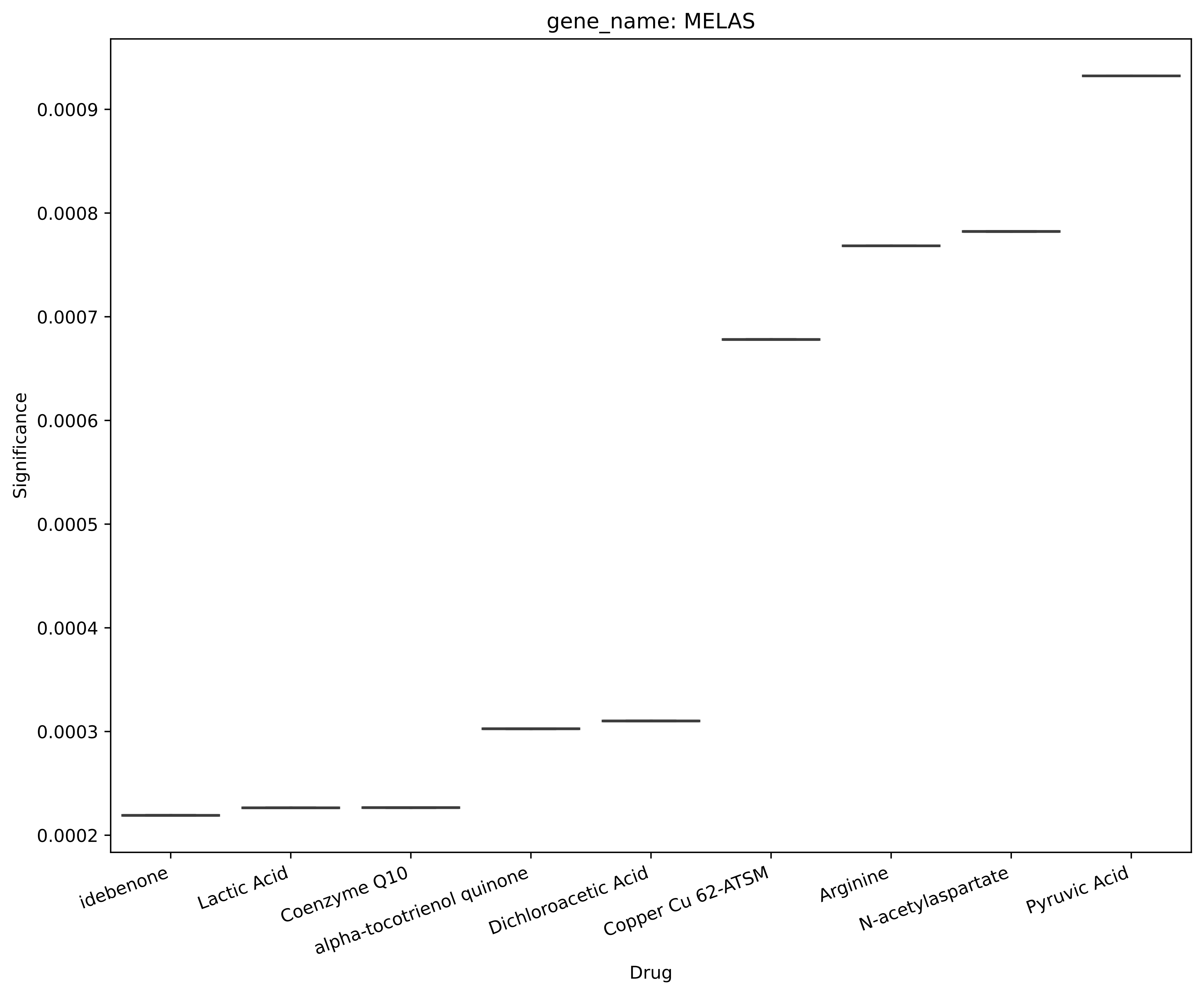
gene_name: MELAS
Significance
0.0009
0.0008
0.0007
0.0006
0.0005
0.0004
0.0003
0.0002
idebenone
Lactic Acid
Coenzyme Q10
alpha-tocotrienol quinone
Dichloroacetic Acid
Copper Cu 62-ATSM
Arginine
N-acetylaspartate
Pyruvic Acid
Drug

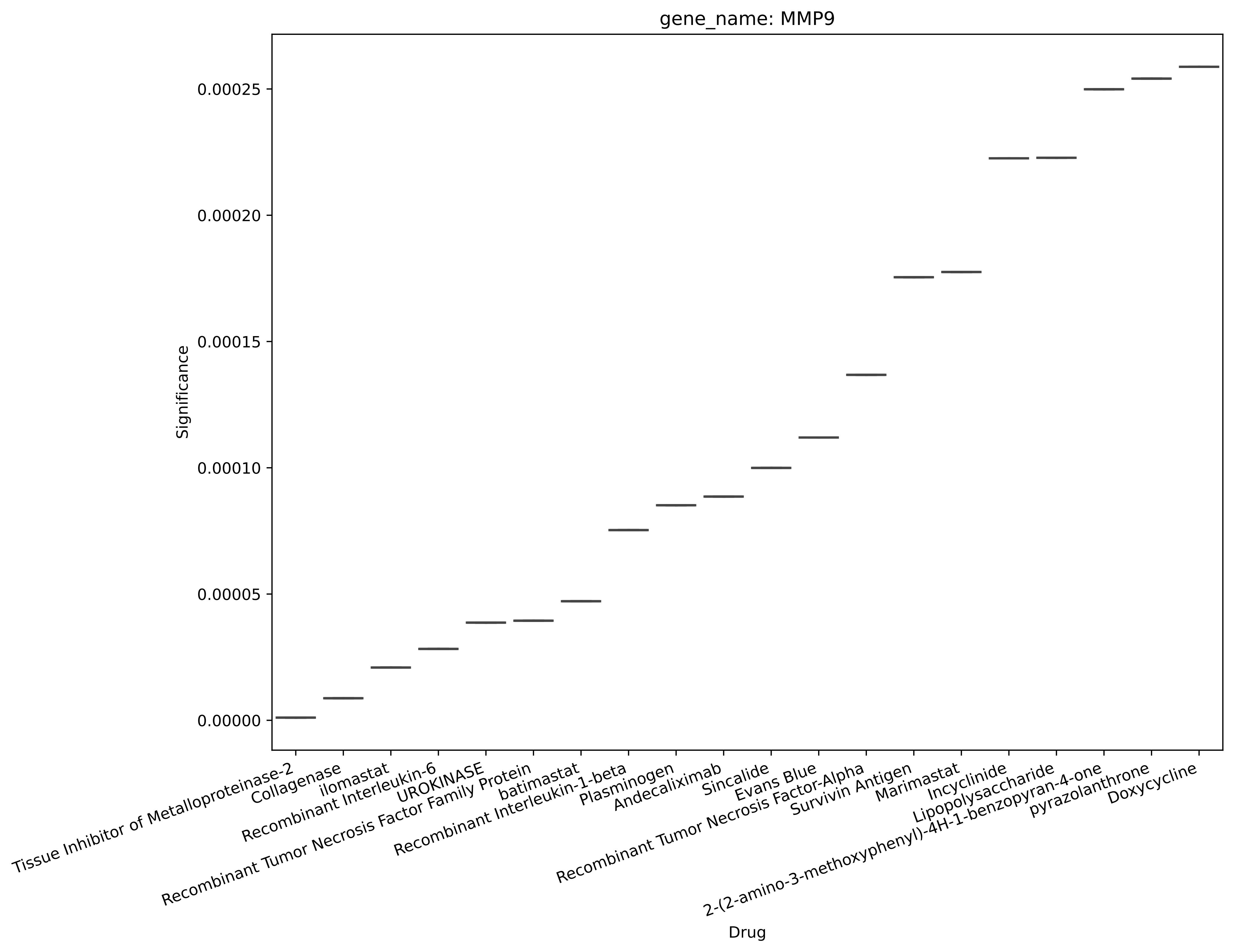
gene_name: MMP9
Significance
0.00025
0.00020
0.00015
0.00010
0.00005
0.00000
Tissue Inhibitor of Metalloproteinase-2
Collagenase
ilomastat
Recombinant Interleukin-6
UROKINASE
Recombinant Tumor Necrosis Factor Family Protein
batimastat
Recombinant Interleukin-1-beta
Plasminogen
Andecaliximab
Sincalide
Evans Blue
Recombinant Tumor Necrosis Factor-Alpha
Survivin Antigen
Marimastat
Incyclinide
Lipopolysaccharide
2-(2-amino-3-methoxyphenyl)-4H-1-benzopyran-4-one
pyrazolanthrone
Doxycycline
Drug

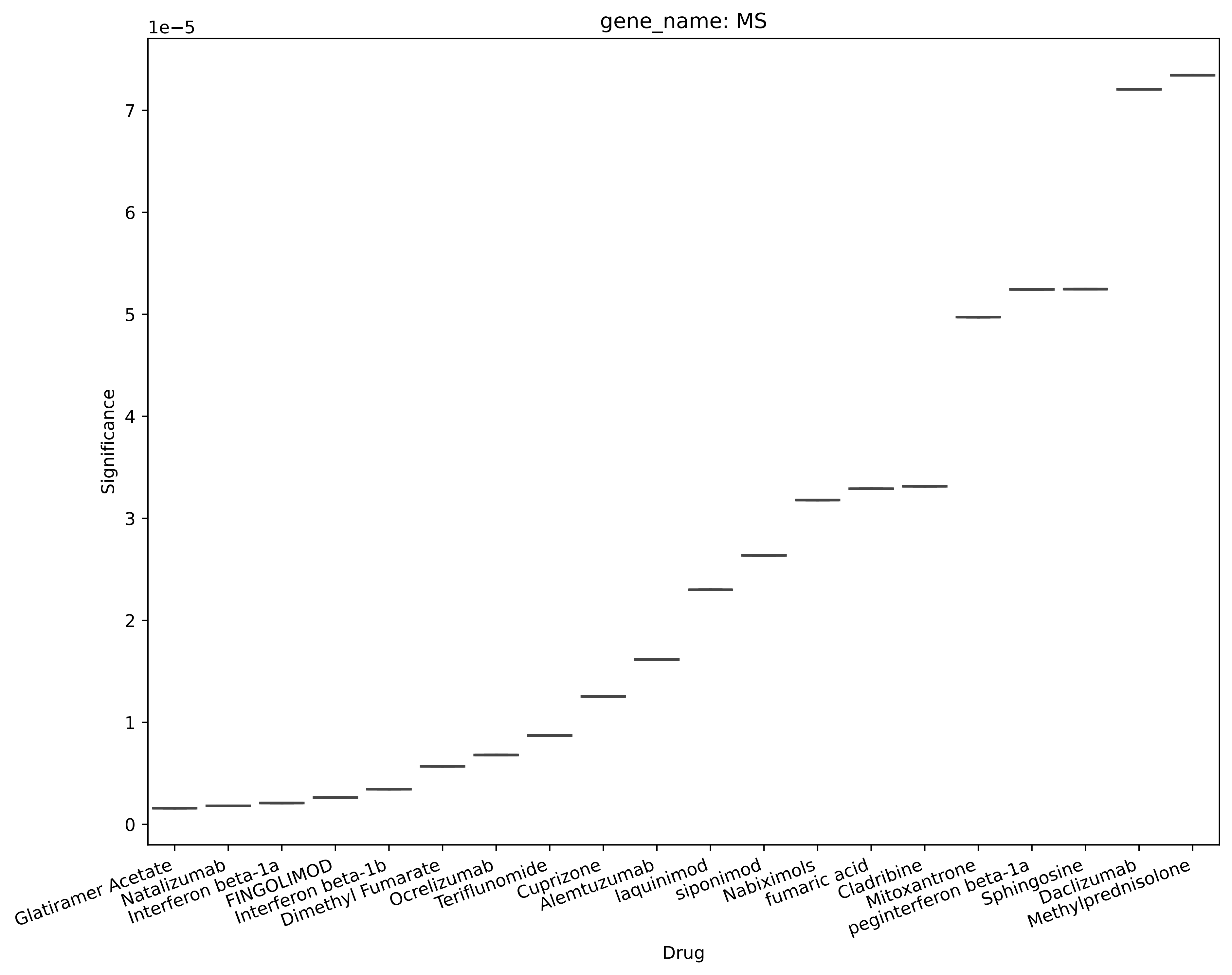
gene_name: MS
1e−5
Significance
0
1
2
3
4
5
6
7
Drug
Glatiramer Acetate
Natalizumab
Interferon beta-1a
FINGOLIMOD
Interferon beta-1b
Dimethyl Fumarate
Ocrelizumab
Teriflunomide
Cuprizone
Alemtuzumab
laquinimod
siponimod
Nabiximols
fumaric acid
Cladribine
Mitoxantrone
peginterferon beta-1a
Sphingosine
Daclizumab
Methylprednisolone

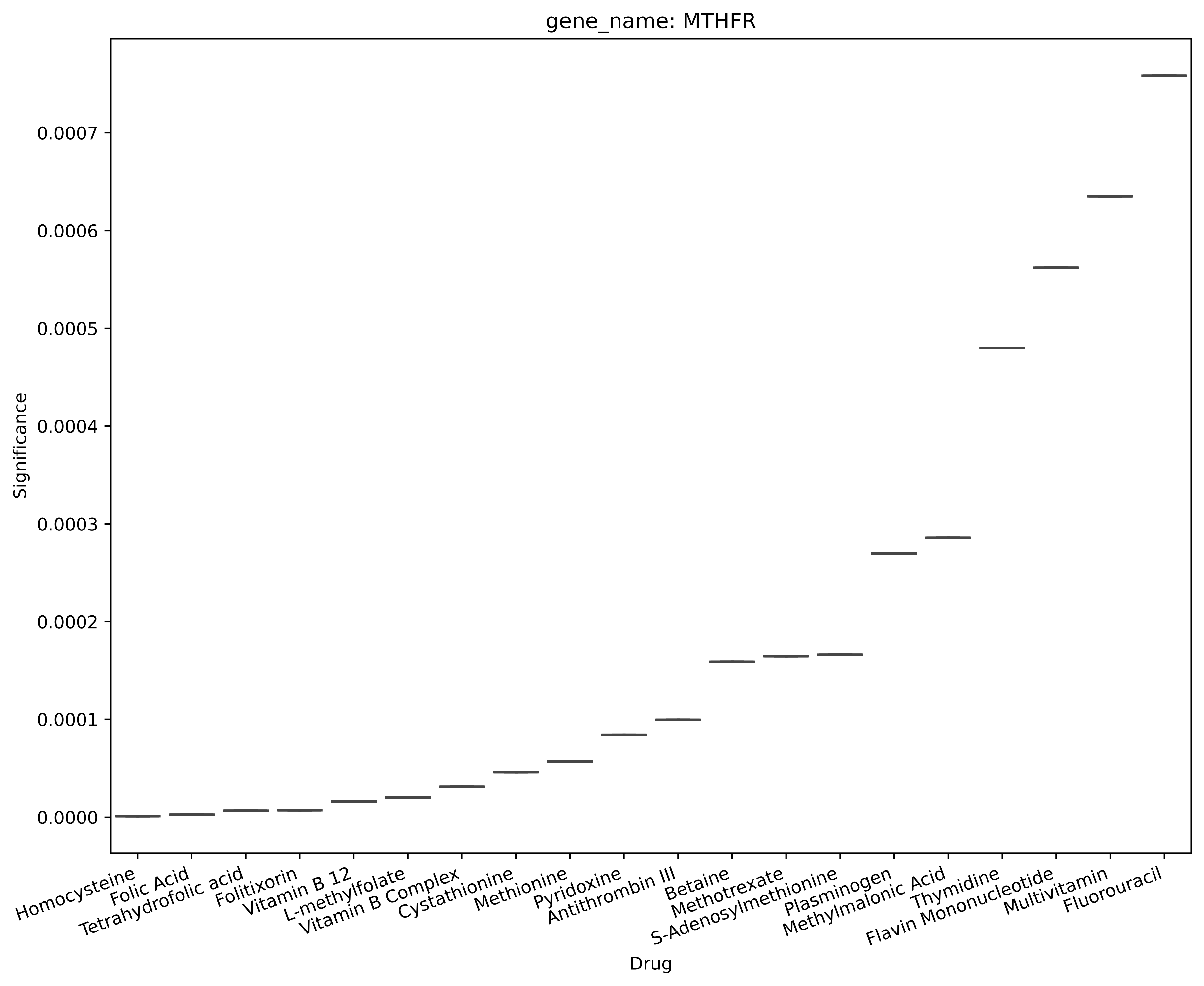
gene_name: MTHFR
Significance
0.0000
0.0001
0.0002
0.0003
0.0004
0.0005
0.0006
0.0007
Drug
Homocysteine
Folic Acid
Tetrahydrofolic acid
Folitixorin
Vitamin B 12
L-methylfolate
Vitamin B Complex
Cystathionine
Methionine
Pyridoxine
Antithrombin III
Betaine
Methotrexate
S-Adenosylmethionine
Plasminogen
Methylmalonic Acid
Thymidine
Flavin Mononucleotide
Multivitamin
Fluorouracil

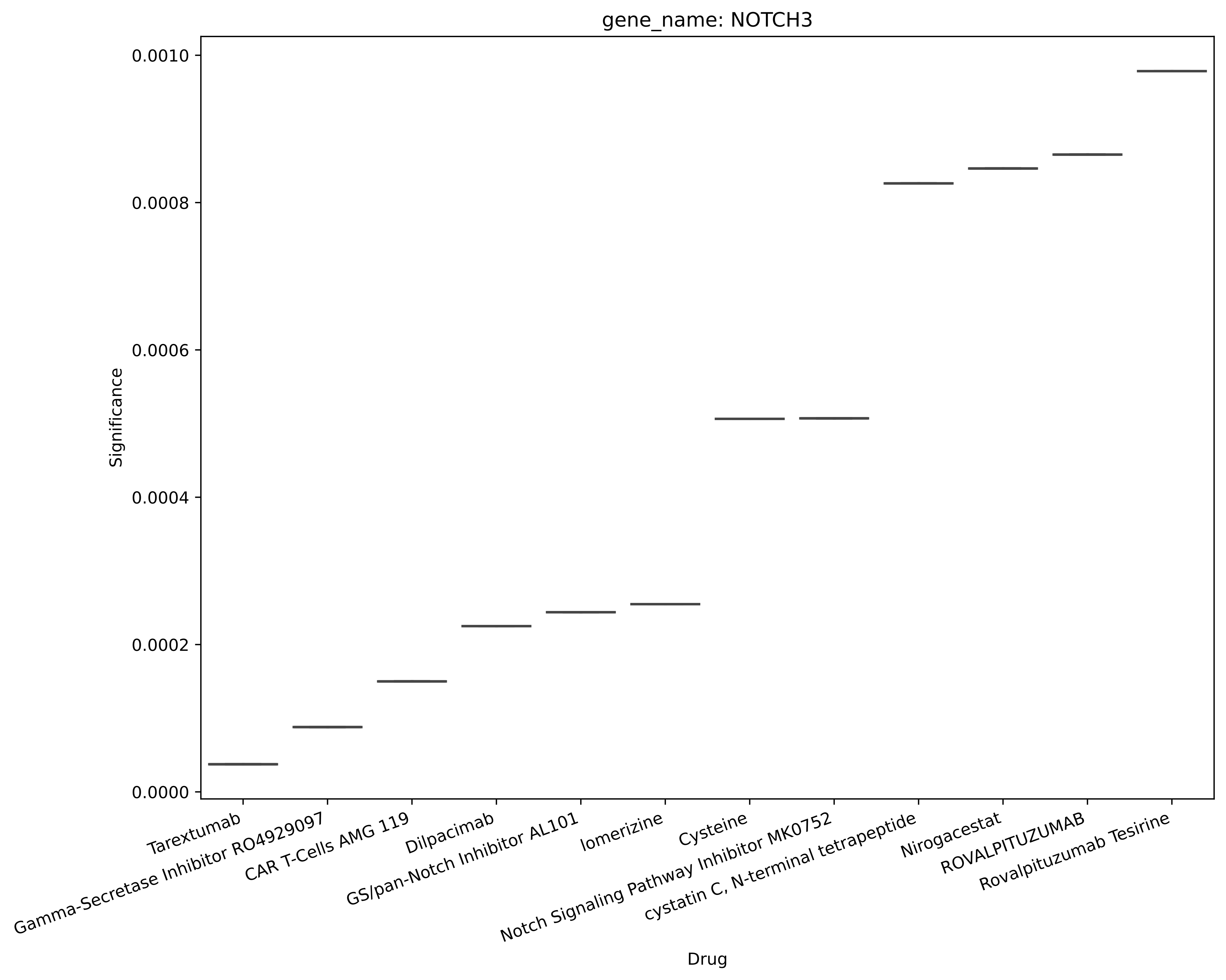

gene_name: NOTCH3
Significance
0.0010
0.0008
0.0006
0.0004
0.0002
0.0000
Tarextumab
Gamma-Secretase Inhibitor RO4929097
CAR T-Cells AMG 119
Dilpacimab
GS/pan-Notch Inhibitor AL101
lomerizine
Cysteine
Notch Signaling Pathway Inhibitor MK0752
cystatin C, N-terminal tetrapeptide
Nirogacestat
ROVALPITUZUMAB
Rovalpituzumab Tesirine
Drug

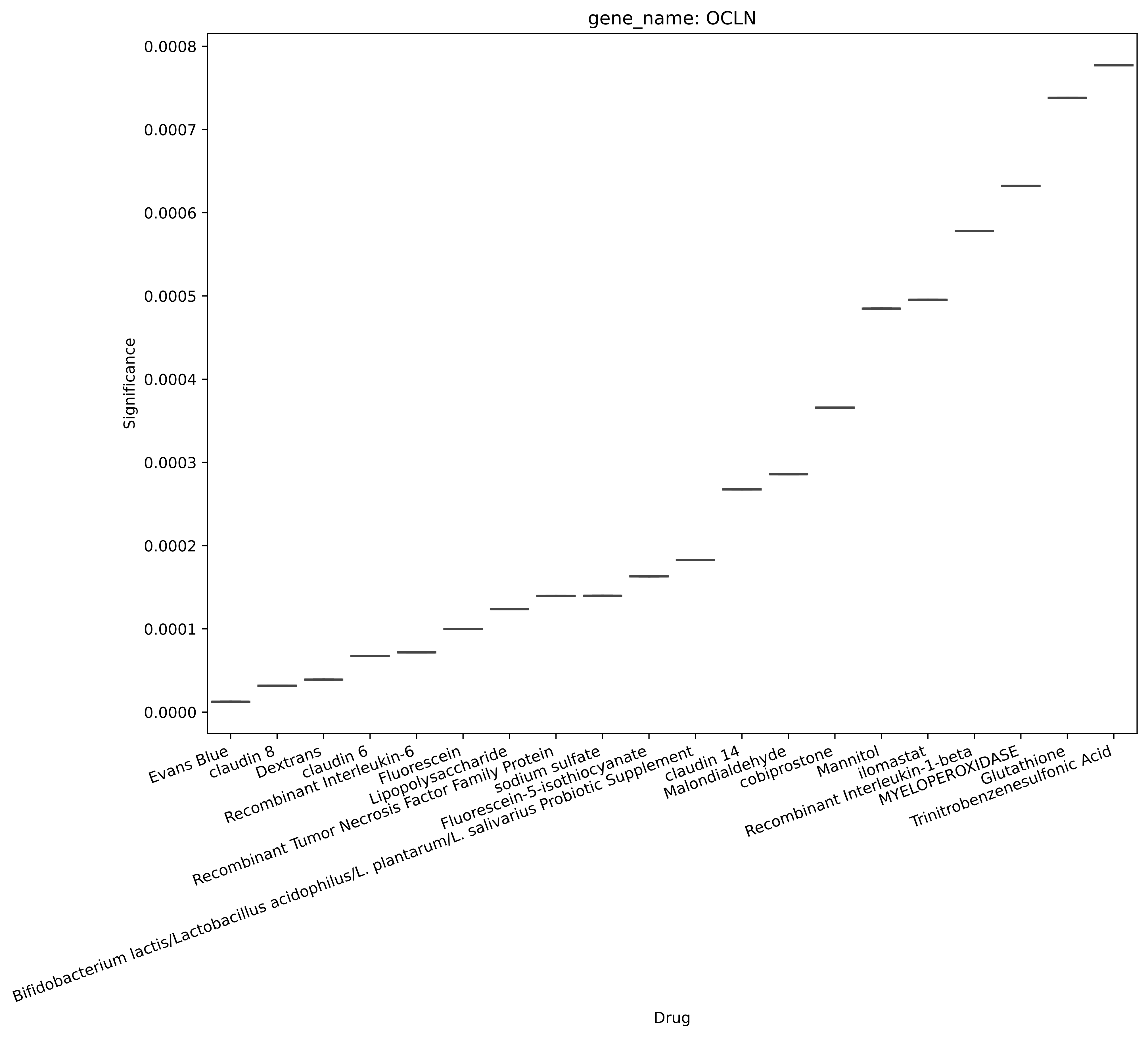
gene_name: OCLN
Significance
0.0008
0.0007
0.0006
0.0005
0.0004
0.0003
0.0002
0.0001
0.0000
Evans Blue
claudin 8
Dextrans
claudin 6
Recombinant Interleukin-6
Fluorescein
Lipopolysaccharide
Recombinant Tumor Necrosis Factor Family Protein
sodium sulfate
Fluorescein-5-isothiocyanate
Bifidobacterium lactis/Lactobacillus acidophilus/L. plantarum/L. salivarius Probiotic Supplement
claudin 14
Malondialdehyde
cobiprostone
Mannitol
ilomastat
Recombinant Interleukin-1-beta
MYELOPEROXIDASE
Glutathione
Trinitrobenzenesulfonic Acid
Drug

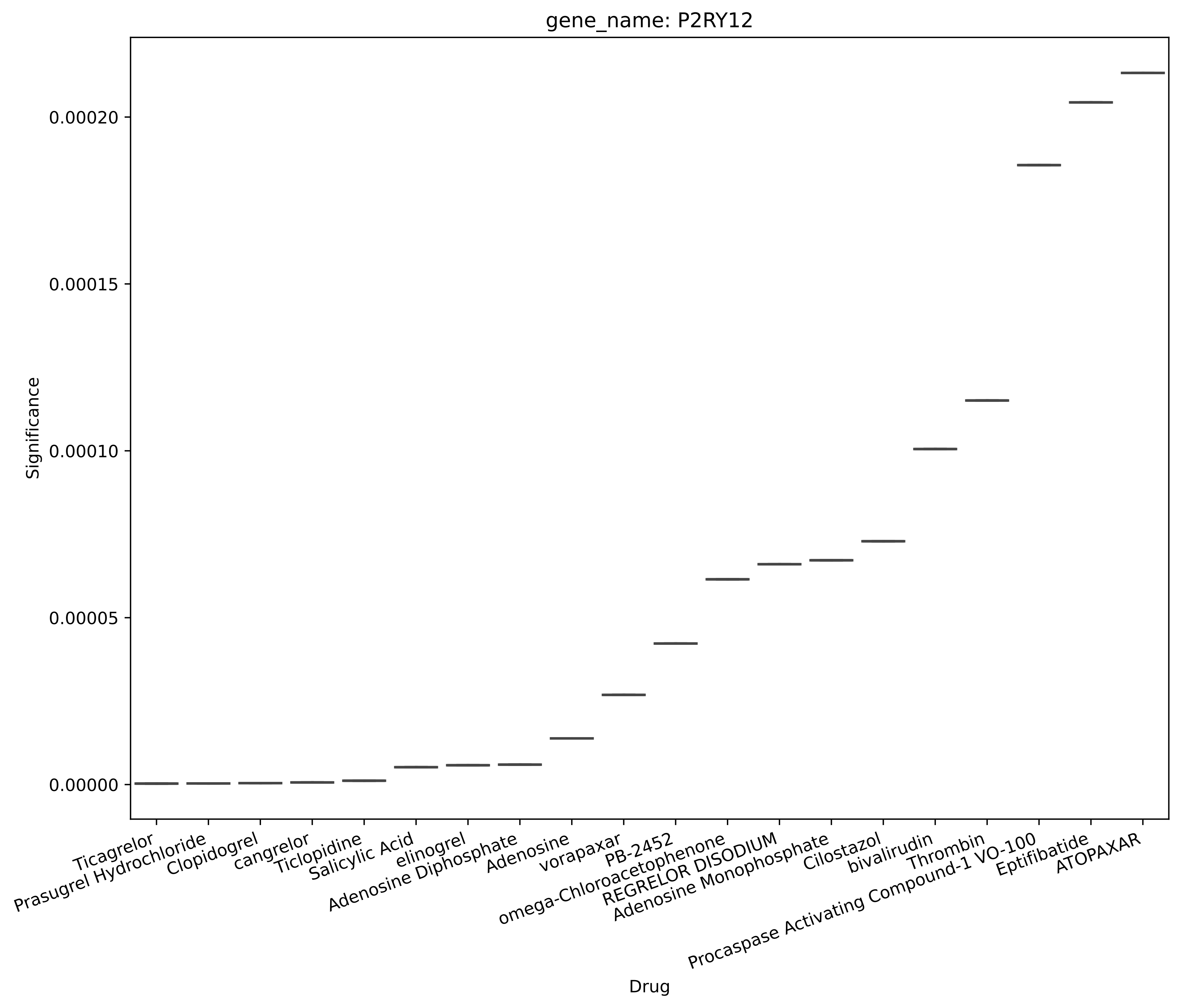
gene_name: P2RY12
Significance
0.00020
0.00015
0.00010
0.00005
0.00000
Ticagrelor
Prasugrel Hydrochloride
Clopidogrel
cangrelor
Ticlopidine
Salicylic Acid
elinogrel
Adenosine Diphosphate
Adenosine
vorapaxar
PB-2452
omega-Chloroacetophenone
REGRELOR DISODIUM
Adenosine Monophosphate
Cilostazol
bivalirudin
Thrombin
Procaspase Activating Compound-1 VO-100
Eptifibatide
ATOPAXAR
Drug

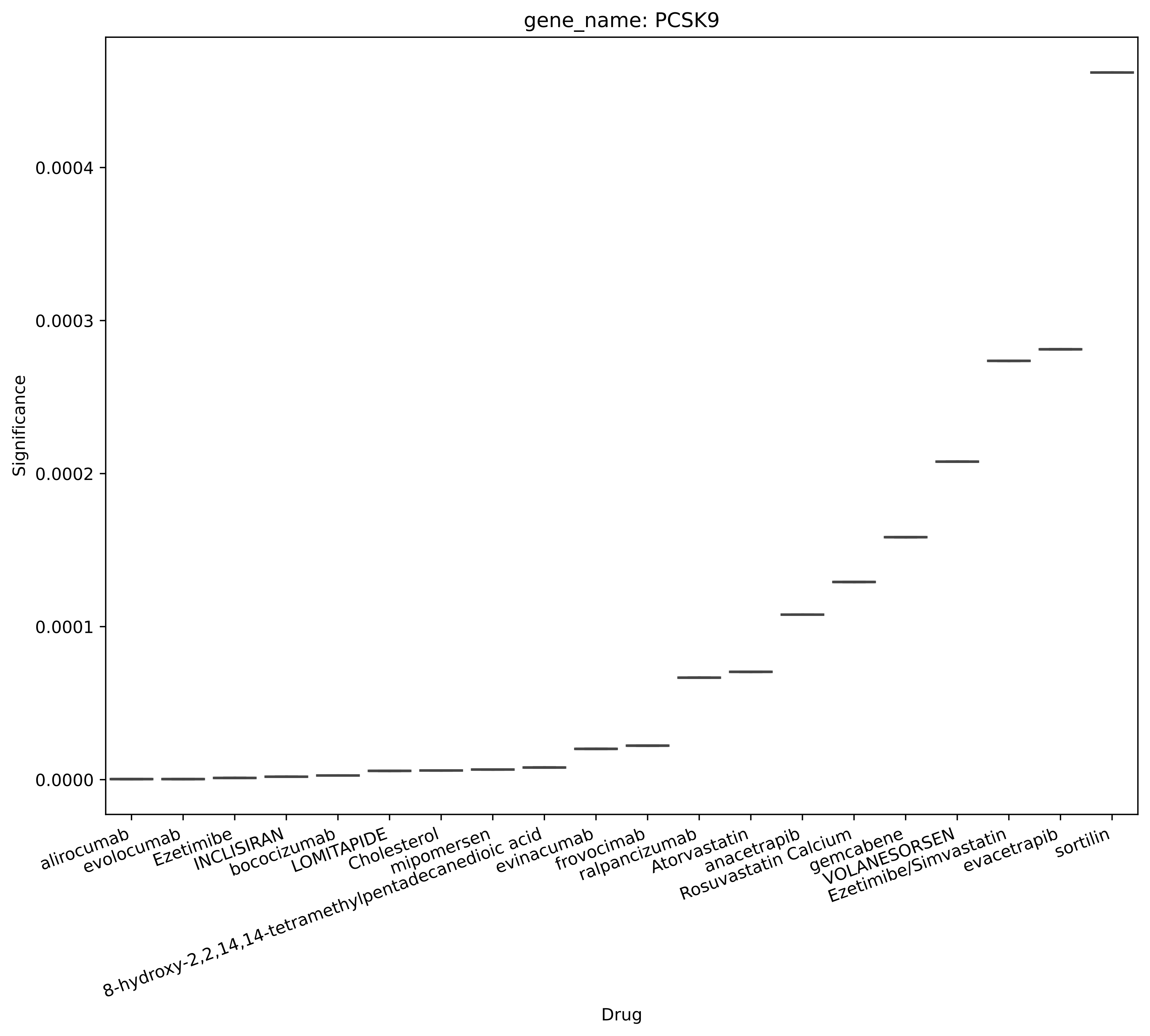
gene_name: PCSK9
Significance
0.0004
0.0003
0.0002
0.0001
0.0000
Drug
alirocumab
evolocumab
Ezetimibe
INCLISIRAN
bococizumab
LOMITAPIDE
Cholesterol
mipomersen
8-hydroxy-2,2,14,14-tetramethylpentadecanedioic acid
evinacumab
frovocimab
ralpancizumab
Atorvastatin
anacetrapib
Rosuvastatin Calcium
gemcabene
VOLANESORSEN
Ezetimibe/Simvastatin
evacetrapib
sortilin

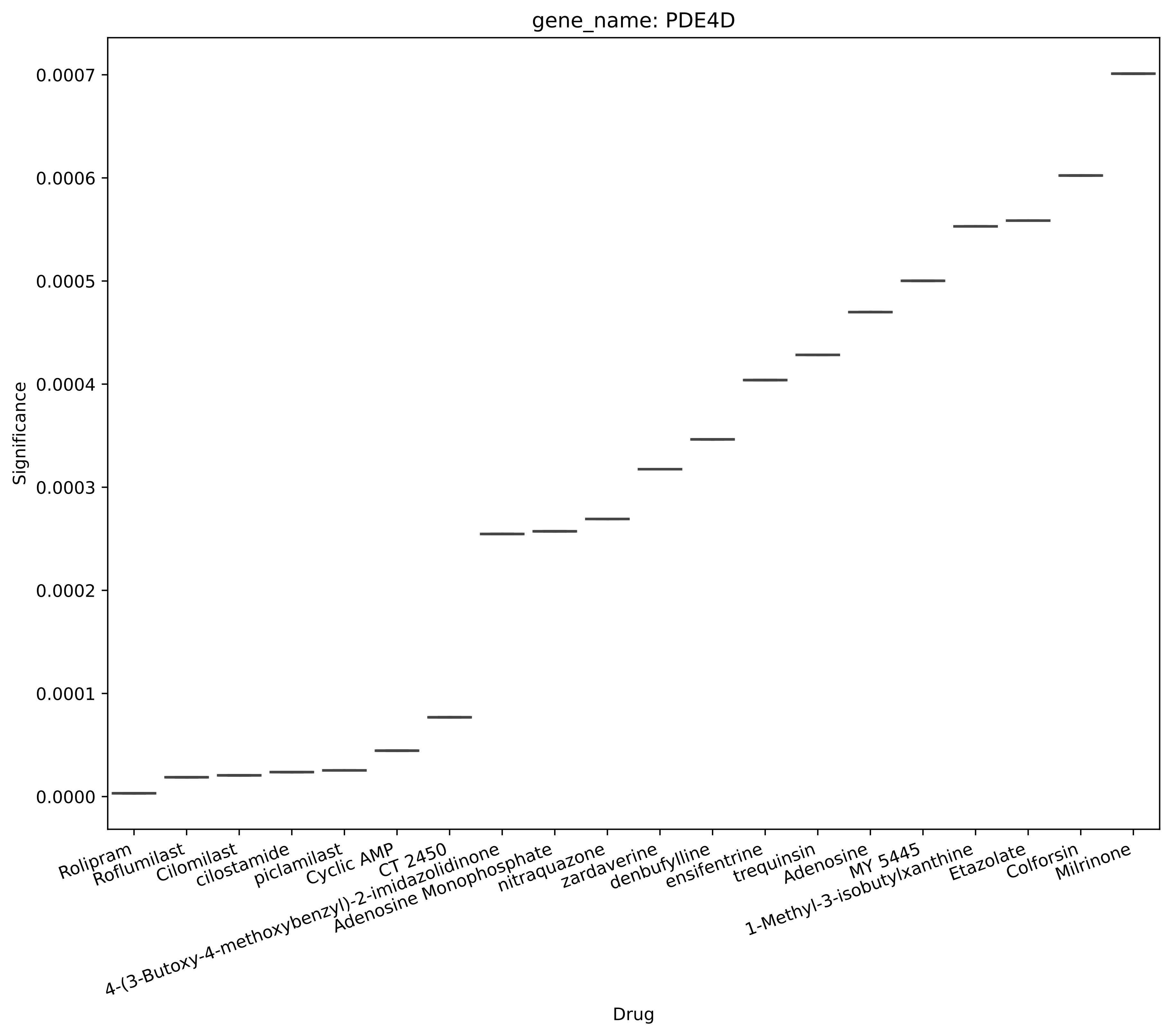
gene_name: PDE4D
Significance
0.0000
0.0001
0.0002
0.0003
0.0004
0.0005
0.0006
0.0007
Drug
Rolipram
Roflumilast
Cilomilast
cilostamide
piclamilast
Cyclic AMP
CT 2450
4-(3-Butoxy-4-methoxybenzyl)-2-imidazolidinone
Adenosine Monophosphate
nitraquazone
zardaverine
denbufylline
ensifentrine
trequinsin
Adenosine
MY 5445
1-Methyl-3-isobutylxanthine
Etazolate
Colforsin
Milrinone

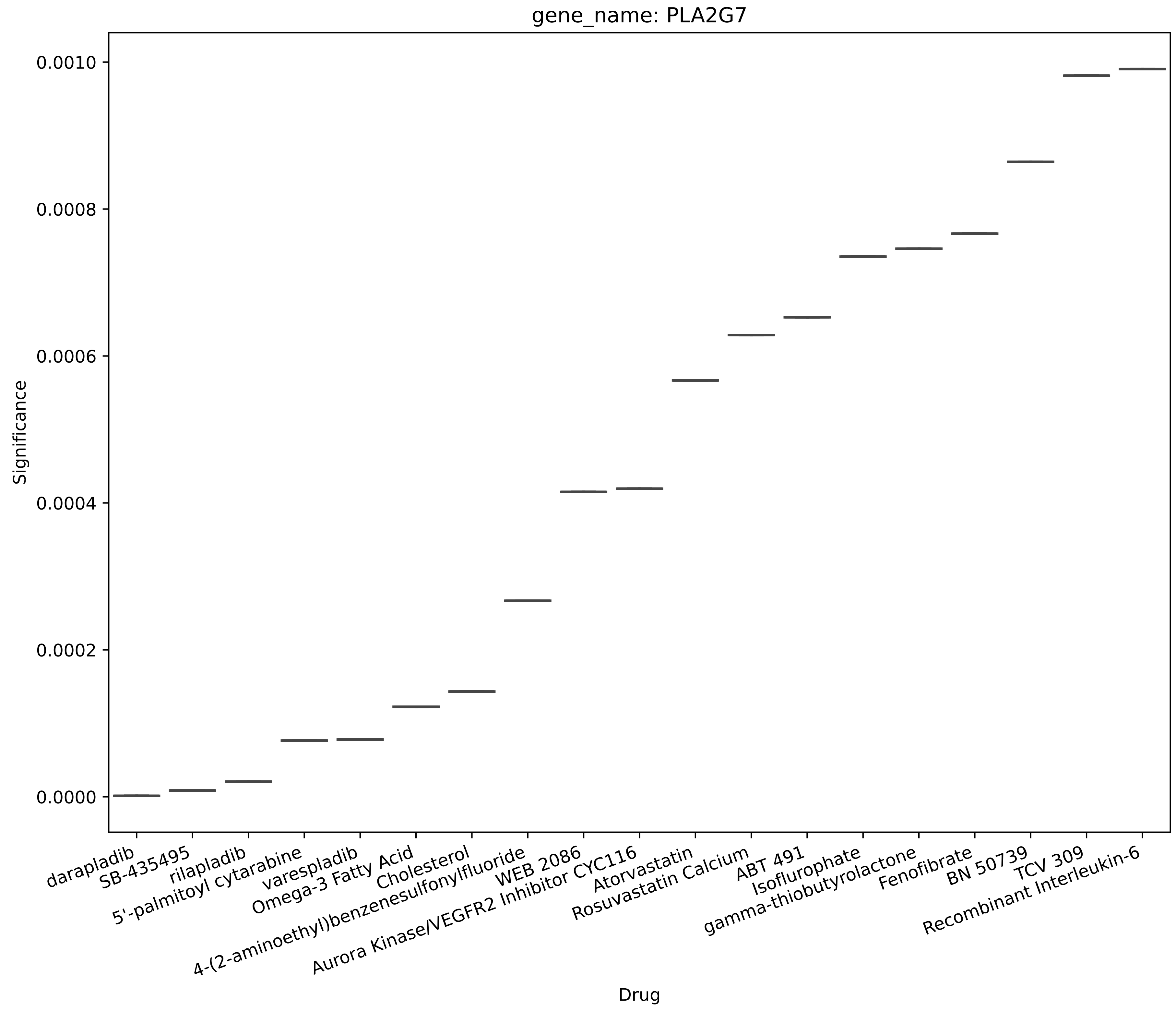
gene_name: PLA2G7
Significance
0.0010
0.0008
0.0006
0.0004
0.0002
0.0000
darapladib
SB-435495
rilapladib
5'-palmitoyl cytarabine
varespladib
Omega-3 Fatty Acid
Cholesterol
4-(2-aminoethyl)benzenesulfonylfluoride
WEB 2086
Aurora Kinase/VEGFR2 Inhibitor CYC116
Atorvastatin
Rosuvastatin Calcium
ABT 491
Isoflurophate
gamma-thiobutyrolactone
Fenofibrate
BN 50739
TCV 309
Recombinant Interleukin-6
Drug

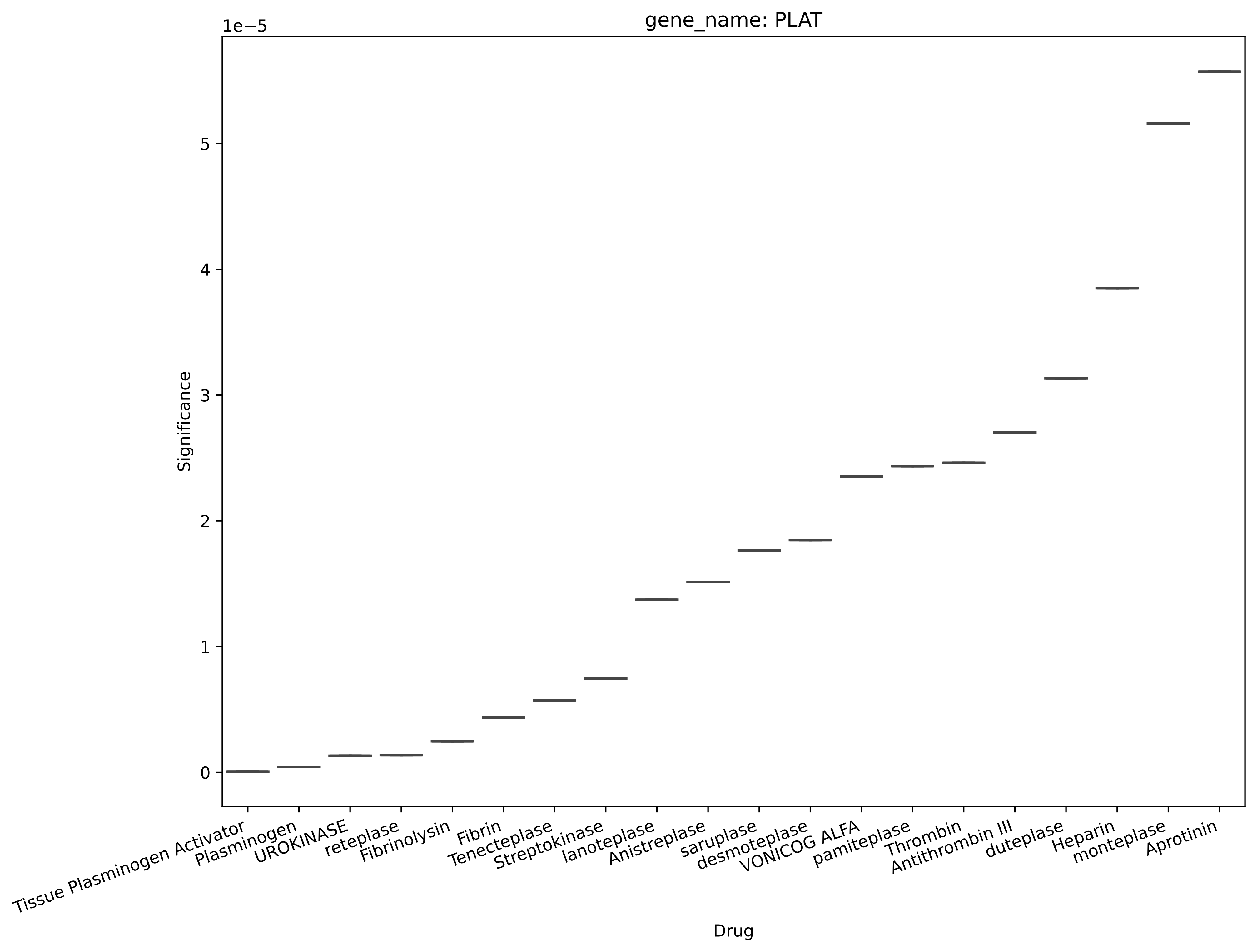
gene_name: PLAT
1e−5
Significance
0
1
2
3
4
5
Drug
Tissue Plasminogen Activator
Plasminogen
UROKINASE
reteplase
Fibrinolysin
Fibrin
Tenecteplase
Streptokinase
lanoteplase
Anistreplase
saruplase
desmoteplase
VONICOG ALFA
pamiteplase
Thrombin
Antithrombin III
duteplase
Heparin
monteplase
Aprotinin

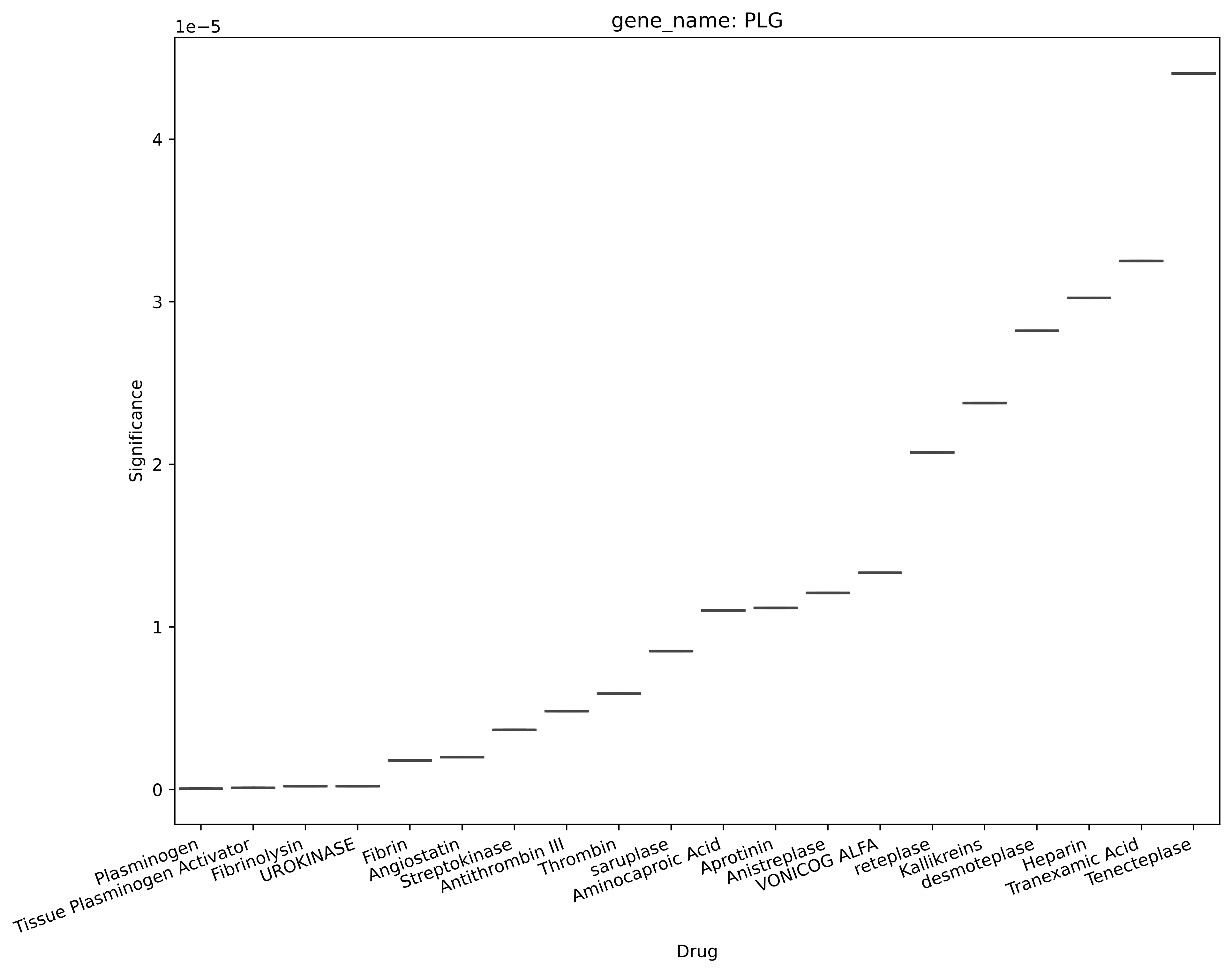

gene_name: PLG
1e−5
4
3
2
1
0
Significance
Plasminogen
Tissue Plasminogen Activator
Fibrinolysin
UROKINASE
Fibrin
Angiostatin
Streptokinase
Antithrombin III
Thrombin
saruplase
Aminocaproic Acid
Aprotinin
Anistreplase
VONICOG ALFA
reteplase
Kallikreins
desmoteplase
Heparin
Tranexamic Acid
Tenecteplase
Drug

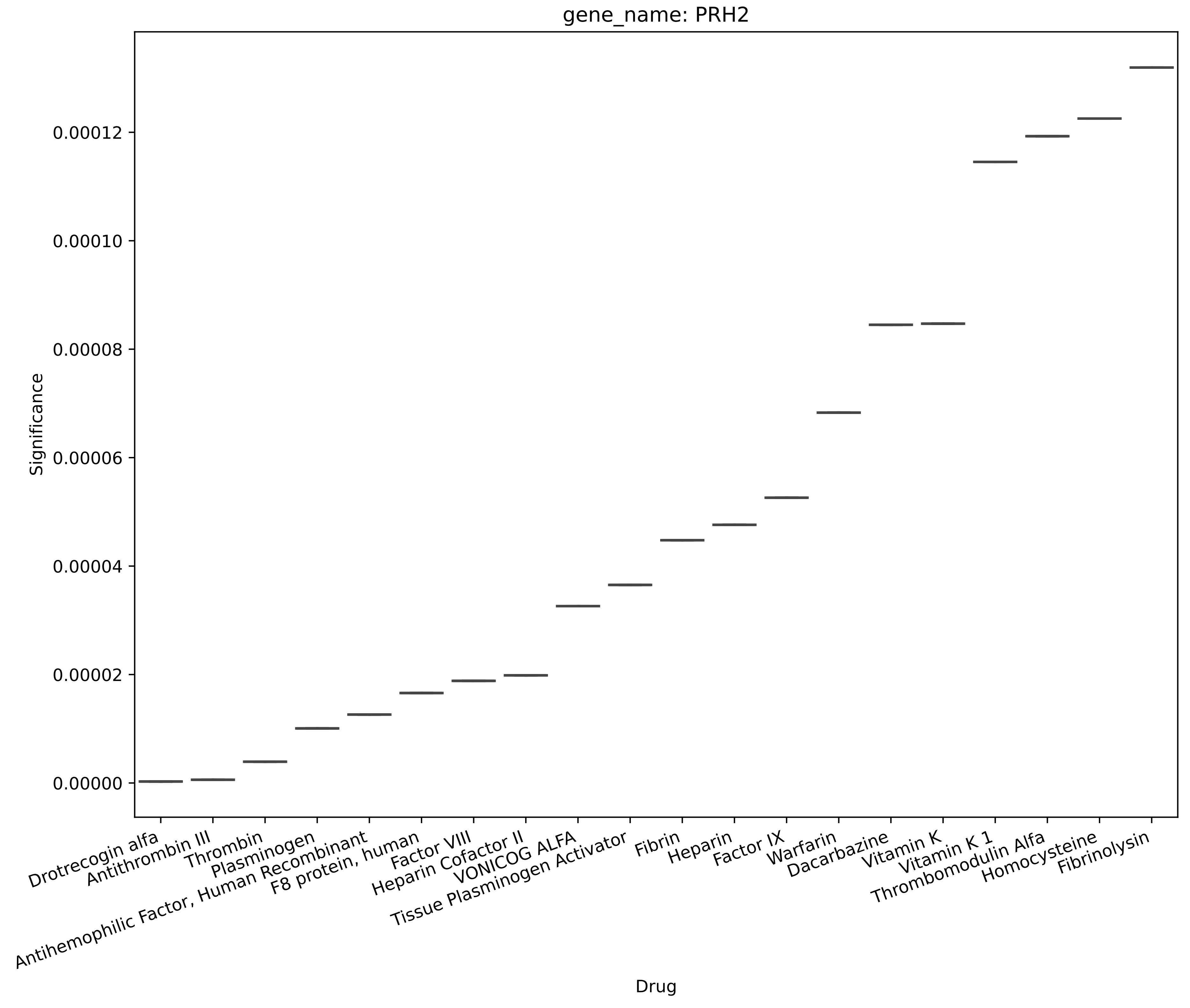
gene_name: PRH2
Significance
0.00012
0.00010
0.00008
0.00006
0.00004
0.00002
0.00000
Drotrecogin alfa
Antithrombin III
Thrombin
Plasminogen
Antihemophilic Factor, Human Recombinant
F8 protein, human
Factor VIII
Heparin Cofactor II
VONICOG ALFA
Tissue Plasminogen Activator
Fibrin
Heparin
Factor IX
Warfarin
Dacarbazine
Vitamin K
Vitamin K 1
Thrombomodulin Alfa
Homocysteine
Fibrinolysin
Drug

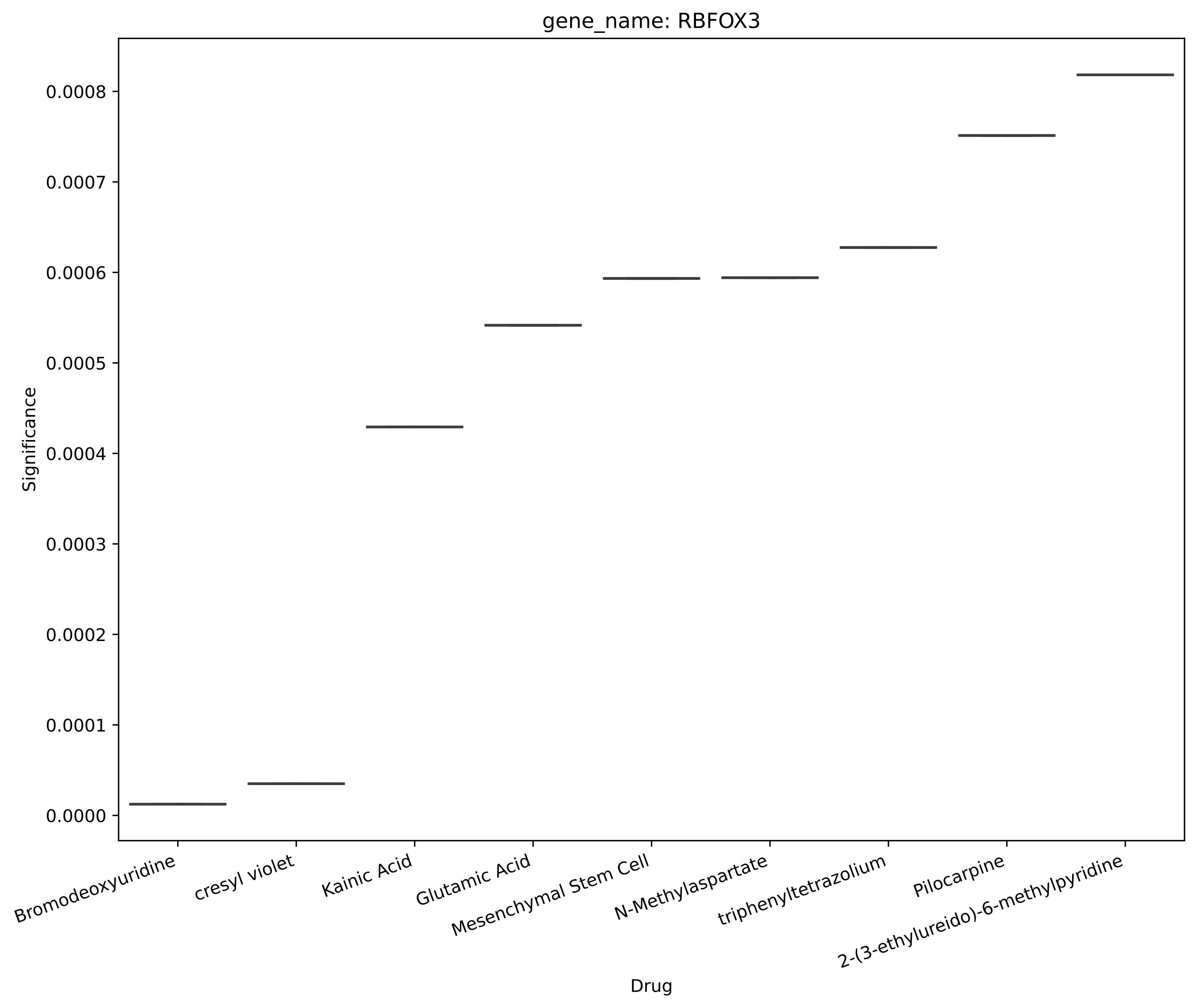
gene_name: RBFOX3
Significance
0.0000
0.0001
0.0002
0.0003
0.0004
0.0005
0.0006
0.0007
0.0008
Bromodeoxyuridine
cresyl violet
Kainic Acid
Glutamic Acid
Mesenchymal Stem Cell
N-Methylaspartate
triphenyltetrazolium
Pilocarpine
2-(3-ethylureido)-6-methylpyridine
Drug

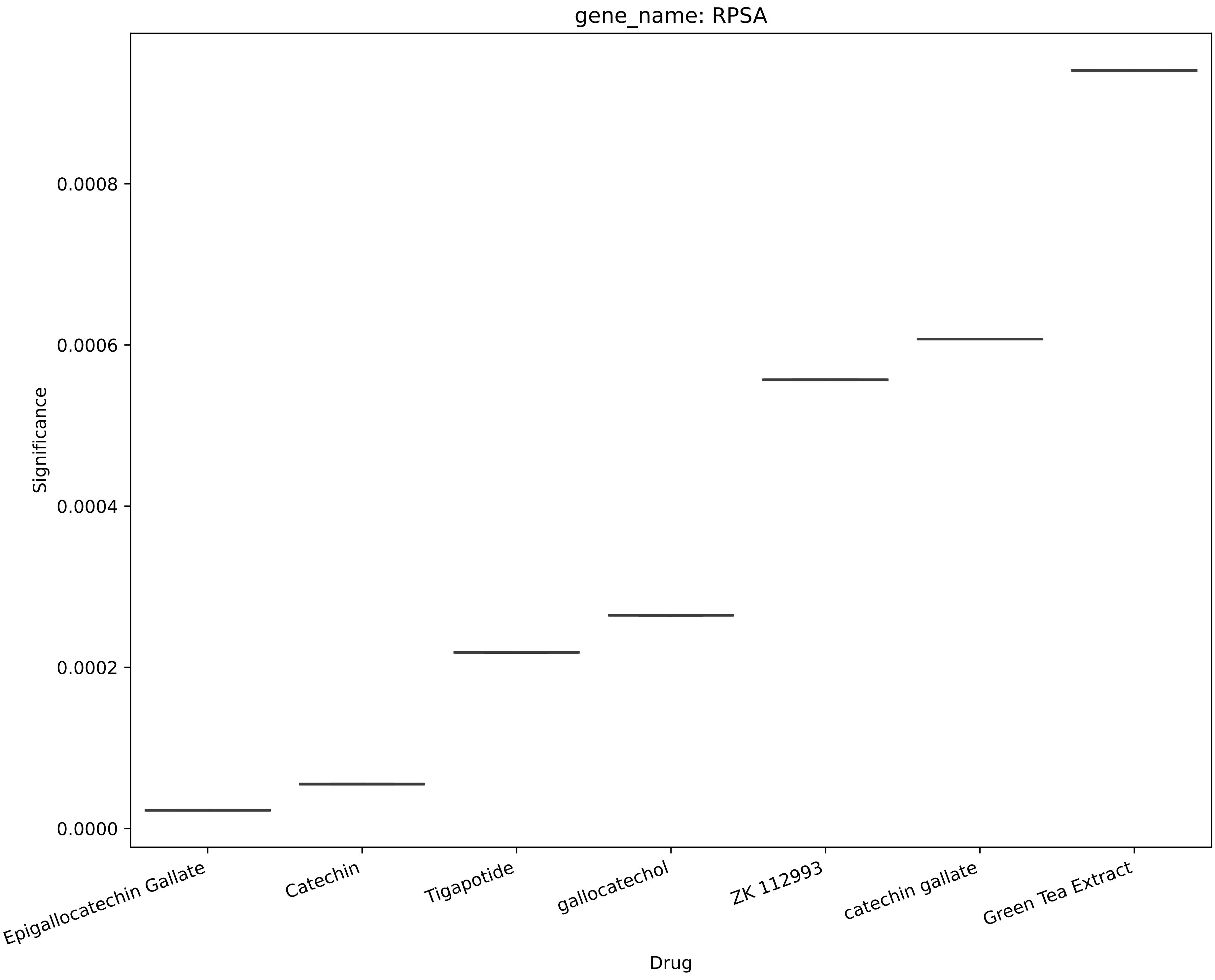
gene_name: RPSA
Significance
0.0000
0.0002
0.0004
0.0006
0.0008
Epigallocatechin Gallate
Catechin
Tigapotide
gallocatechol
ZK 112993
catechin gallate
Green Tea Extract
Drug

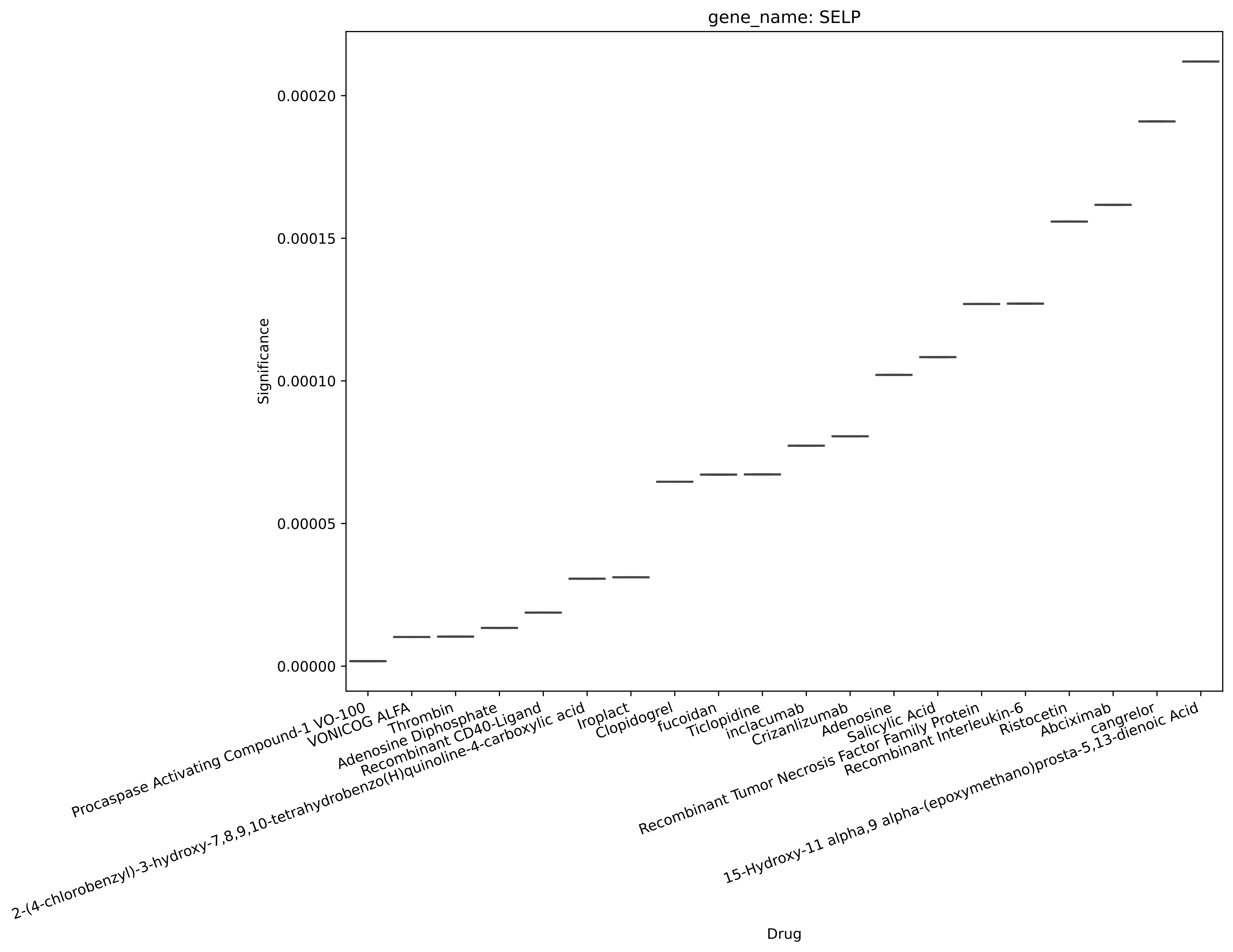
gene_name: SELP
Significance
0.00000
0.00005
0.00010
0.00015
0.00020
Procaspase Activating Compound-1 VO-100
VONICOG ALFA
Thrombin
Adenosine Diphosphate
Recombinant CD40-Ligand
2-(4-chlorobenzyl)-3-hydroxy-7,8,9,10-tetrahydrobenzo(H)quinoline-4-carboxylic acid
Iroplact
Clopidogrel
fucoidan
Ticlopidine
inclacumab
Crizanlizumab
Adenosine
Salicylic Acid
Recombinant Tumor Necrosis Factor Family Protein
Recombinant Interleukin-6
Ristocetin
Abciximab
cangrelor
15-Hydroxy-11 alpha,9 alpha-(epoxymethano)prosta-5,13-dienoic Acid
Drug

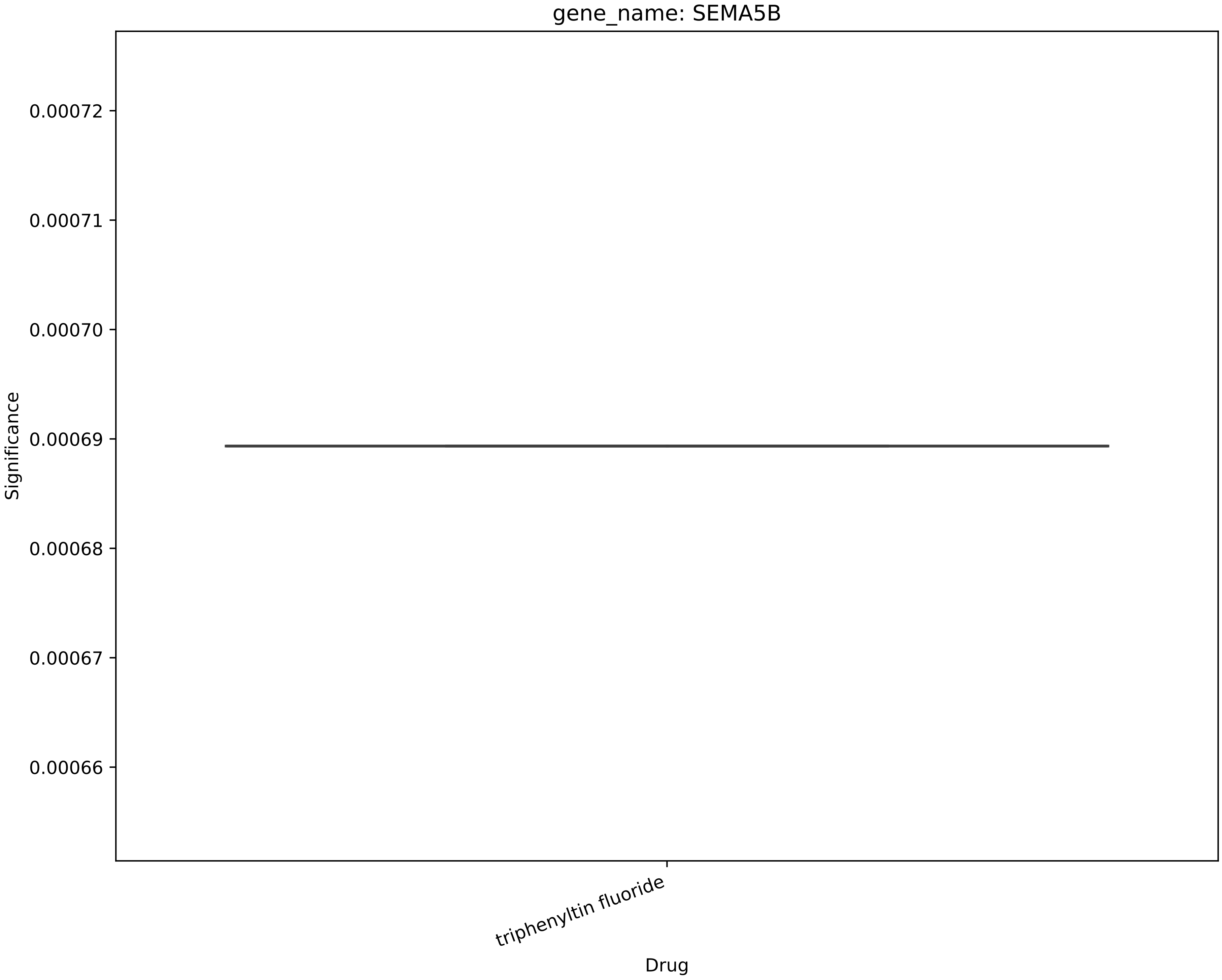

gene_name: SEMA5B
0.00072
0.00071
0.00070
0.00069
0.00068
0.00067
0.00066
Significance
triphenyltin fluoride
Drug

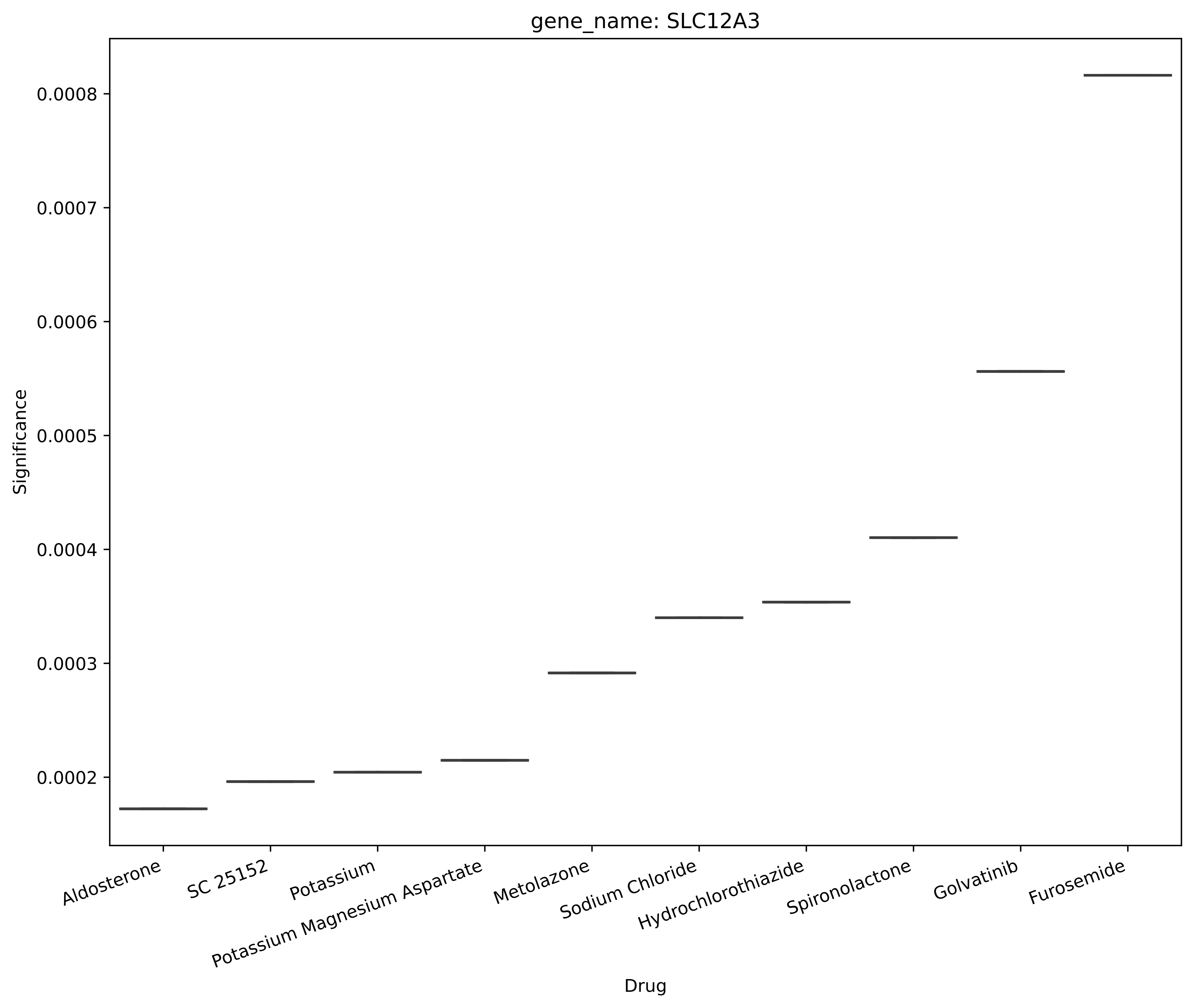
gene_name: SLC12A3
Significance
0.0008
0.0007
0.0006
0.0005
0.0004
0.0003
0.0002
Aldosterone
SC 25152
Potassium
Potassium Magnesium Aspartate
Metolazone
Sodium Chloride
Hydrochlorothiazide
Spironolactone
Golvatinib
Furosemide
Drug

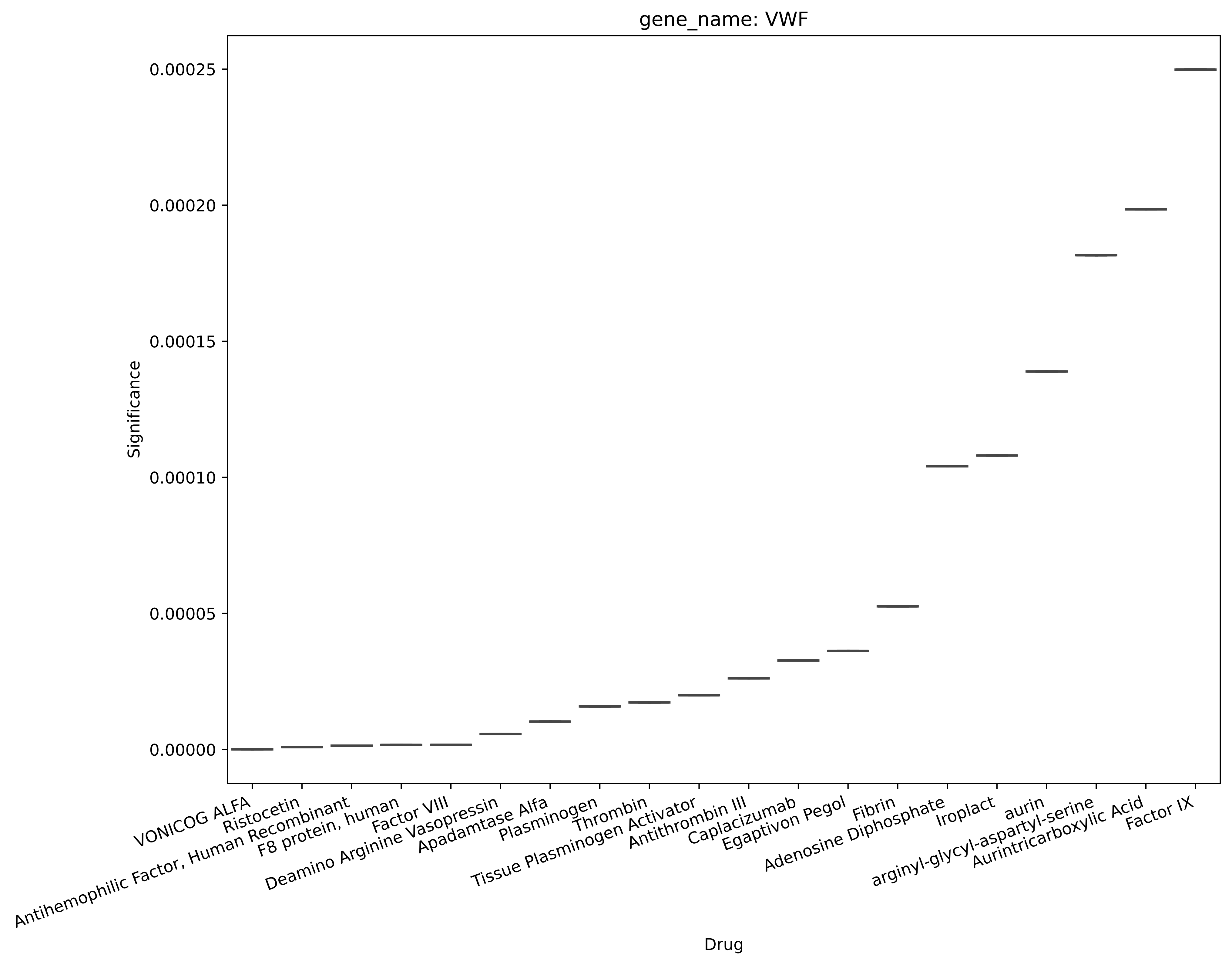
gene_name: VWF
Significance
0.00025
0.00020
0.00015
0.00010
0.00005
0.00000
VONICOG ALFA
Ristocetin
Antihemophilic Factor, Human Recombinant
F8 protein, human
Factor VIII
Deamino Arginine Vasopressin
Apadamtase Alfa
Plasminogen
Thrombin
Tissue Plasminogen Activator
Antithrombin III
Caplacizumab
Egaptivon Pegol
Fibrin
Adenosine Diphosphate
Iroplact
aurin
arginyl-glycyl-aspartyl-serine
Aurintricarboxylic Acid
Factor IX
Drug
