## Supplementary figures and images for "Recommending Drug Combinations using Reinforcement Learning to target Genes/proteins that cause Stroke: A comprehensive Systematic Review and Network Meta-analysis"

### v36_sm1.pdf

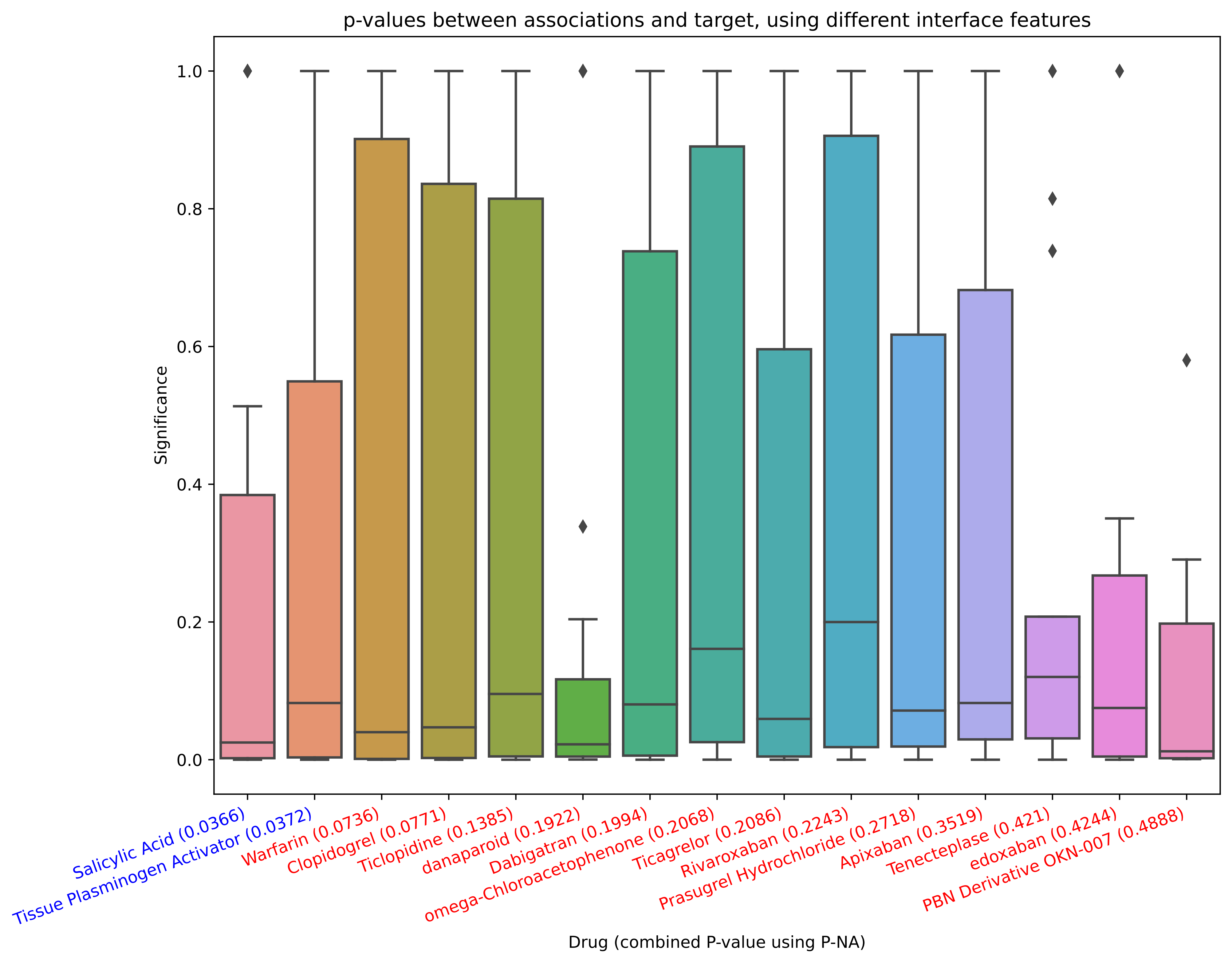

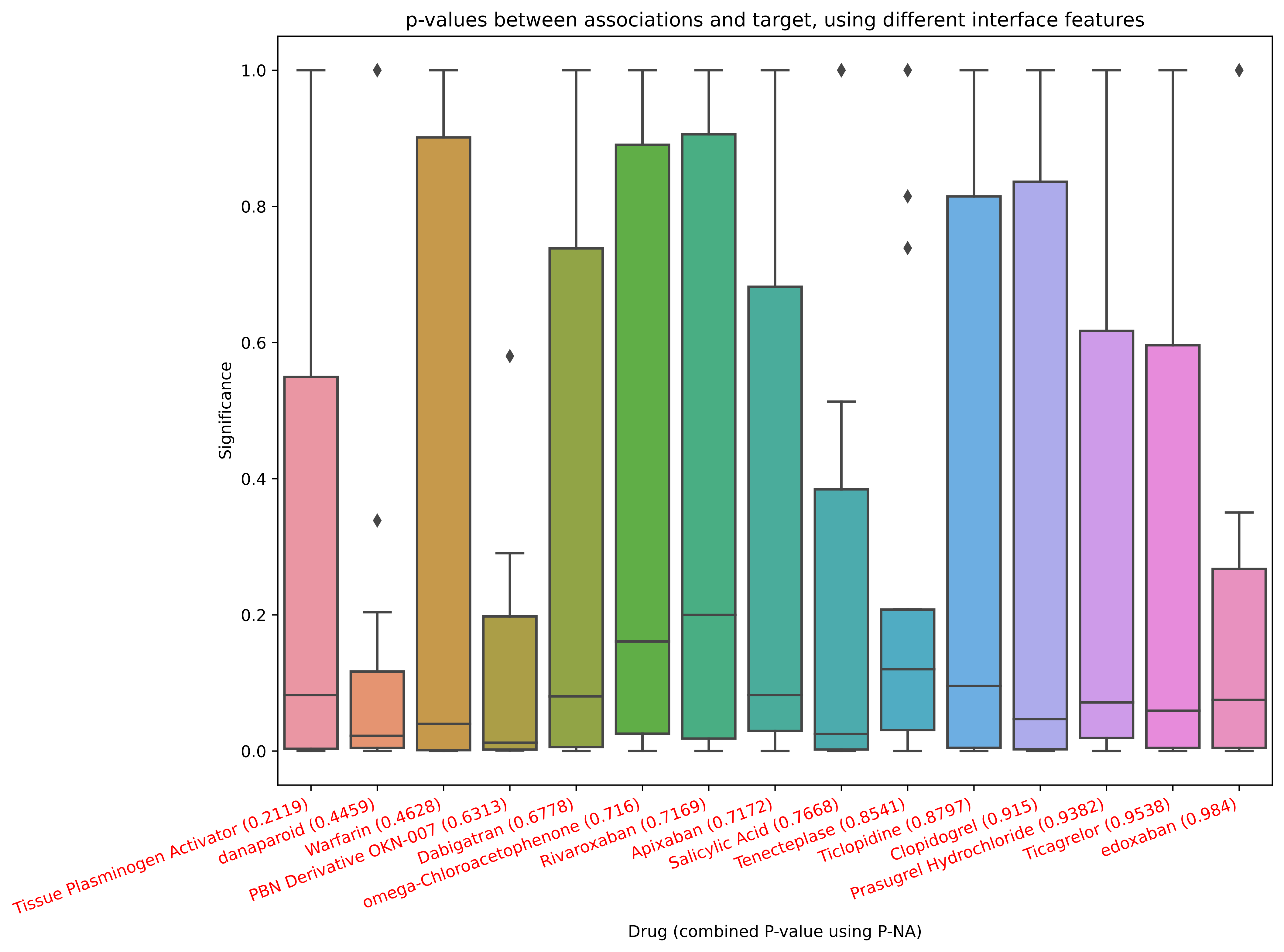

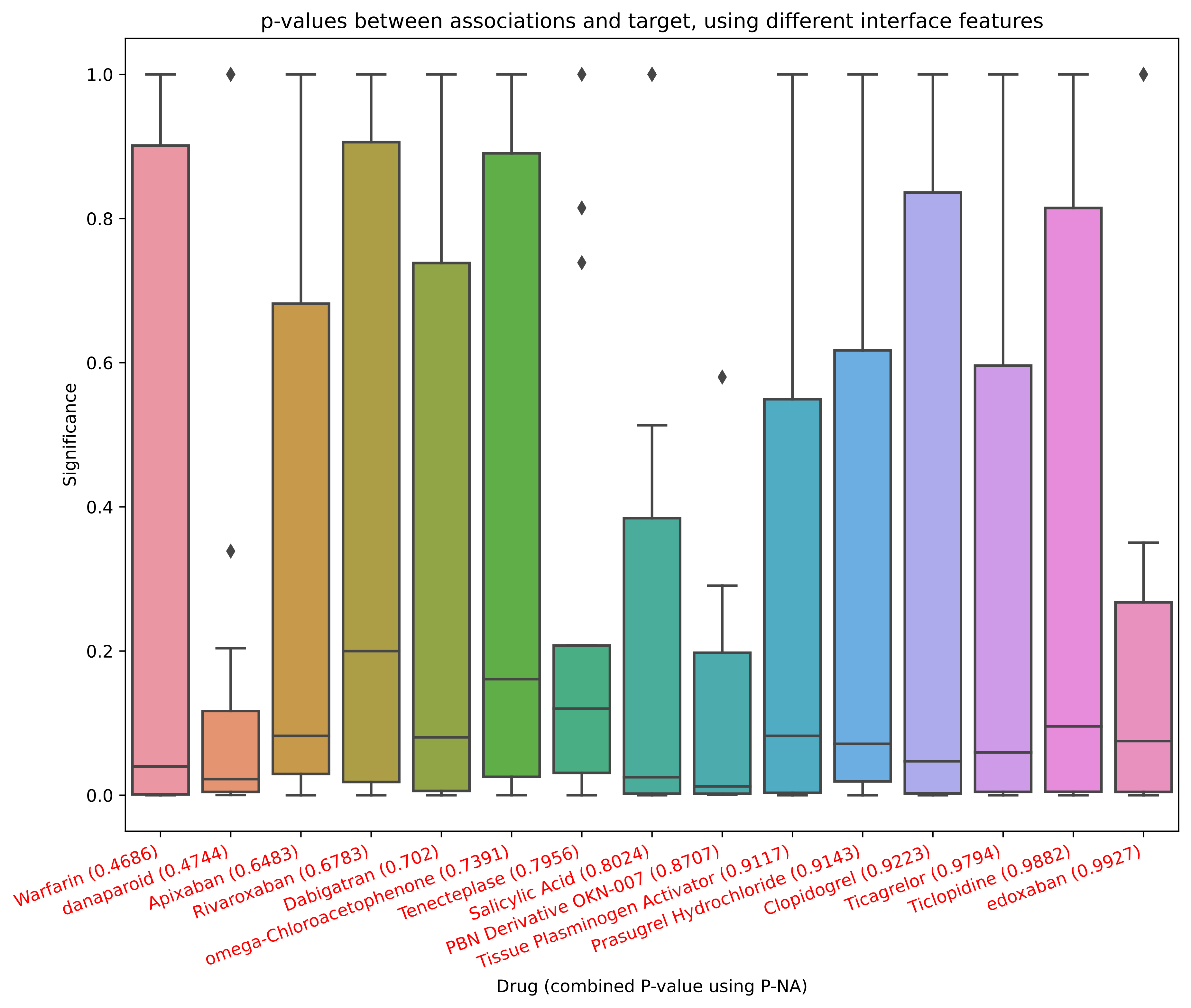
